# Transcriptional and genetic sex differences for schizophrenia across the dorsolateral prefrontal cortex, hippocampus, and caudate nucleus

**DOI:** 10.1101/2022.09.30.22280452

**Authors:** Kynon JM Benjamin, Ria Arora, Joshua M Stolz, Laura D’Ignazio, Leonardo Collado-Torres, Thomas M Hyde, Joel E Kleinman, Daniel R Weinberger, Apuã CM Paquola, Jennifer A Erwin

**Author notes:** Co-first authors (Kynon JM Benjamin, Ria Arora).

## Abstract

Schizophrenia is a complex neuropsychiatric disorder with sexually dimorphic features, including differential symptomatology, drug responsiveness, and male incidence rate. To date, only the prefrontal cortex has been examined in large-scale transcriptome analyses for sex differences in schizophrenia. Here, we examined the BrainSeq Consortium RNA-sequencing and genotypes for the caudate nucleus (n=399), dorsolateral prefrontal cortex (DLPFC; n=377), and hippocampus (n=394) to characterize sex differences in schizophrenia. We identified genomic features (genes, transcripts, exons, and exon-exon junctions) associated with sex, sex-specific expression in schizophrenia, and sex-interacting expression quantitative trait loci (si-eQTL) associated with schizophrenia risk. We found 878 unique genes with sex differences across brain regions, including *ANK3*, which shows male-biased expression in the caudate nucleus. X-chromosome dosage was significantly decreased in the hippocampus of female and male individuals with schizophrenia. Our sex interaction model revealed 15 novel junctions dysregulated for schizophrenia in a sex-specific manner. Sex-specific schizophrenia analysis identified dozens of expressed, sex-specific features with enrichment in the transcriptional response of cellular stress. Finally, our si-eQTL analysis revealed 974 unique genes, 14 of which are associated with schizophrenia risk. Overall, our results increased the number of annotated sex-biased features, identified sex-specific schizophrenia genes, and provided the first annotation of si-eQTL in the human DLPFC and hippocampus. Altogether, these results point to the importance of sex-informed analysis of sexually dimorphic traits and inform personalized therapeutic strategies in schizophrenia.

## Introduction

For more than a century, sex differences have been observed in schizophrenia – a complex, chronic neuropsychiatric disorder affecting approximately 1% of the adult population worldwide. These sex differences include differences in cognitive severity and age of onset: for example, women appearing to be less vulnerable to altered verbal processing deficits^1^, and men having an earlier age of disease onset^2, 3^. Additionally, prenatal stress may significantly increase the risk of schizophrenia in men compared to women^4, 5^. To date, only two large-scale RNA-sequencing studies examining sex differences in schizophrenia^6, 7^, focusing exclusively on one brain region – the prefrontal cortex. Furthermore, the GTEx analyses of sex differences across 45 tissues found fewer than 100 differentially expressed genes in 13 of 14 brain regions. This small number of identified, differentially expressed genes might be attributed to limited sample size (114 to 209 individuals)^8^.

Recent large-scale studies have used statistical association between genotype and expression (expression quantitative trait loci [eQTL]) with schizophrenia genome-wide association studies (GWAS)^9, 10^ to identify genomic features (gene, transcript, exon, and exon-exon junctions) underlying schizophrenia risk^11–15^. However, these studies have not explored potential sex-interacting eQTL (si-eQTL). Furthermore, the GTEx study across 44 tissues found only four si-eQTL genes with a nominally significance of FDR < 0.25 in only two of the 13 brain regions examined^8^. Other recent si-eQTL studies involving whole blood^16, 17^ and lymphoblastoid cell lines^18^ have found fewer than 25 si-eQTL. As eQTLs have tissue specificity^19^, the field of neuropsychiatric genetics needs a sizable and comprehensive analysis of si-eQTL in the human brain.

Here, we leverage the BrainSeq Consortium RNA-sequencing and genotypes datasets to identify genes associated with sex, sex-specific expression in schizophrenia, and sex-interacting eQTL using a total of 1170 samples across 504 individuals (**Table 1**) for the caudate nucleus (n=399), dorsolateral prefrontal cortex (DLPFC; n=377), and hippocampus (n=394). Our work increases the number of annotated sex-biased features, examines sex-chromosome dosage, identifies sex-specific schizophrenia features, provides the first annotation of si-eQTL in the human DLPFC and hippocampus, and increases si-eQTL annotations for the caudate nucleus. Altogether, these results provide novel insights into sex differences, highlighting the importance of sex-informed analysis of sexually dimorphic traits, and informing personalized therapeutic strategies in schizophrenia.

**Table 1.** A sample breakdown of eQTL analysis for individuals (age > 13) postmortem caudate nucleus, DLPFC, and hippocampus from the BrainSeq Consortium, separated by sex. Abbreviations: Female (F), Male (M), Neurotypical Control (CTL), Schizophrenia (SZ), African American (AA), European American (EA).

| Brain Region | Sex | Sample Size | Diagnosis | Ancestry | Age (mean ± sd) | RIN (mean ± sd) |
| --- | --- | --- | --- | --- | --- | --- |
| Caudate Nucleus | F | 126 | 76CTL/50SZ | 79AA/47EA | 50.2 ± 16.9 | 7.8 ± 0.9 |
| | M | 273 | 169CTL/104SZ | 127AA/146EA | 48.6 ± 15.7 | 7.9 ± 0.8 |
| DLPFC | F | 121 | 73CTL/48SZ | 75AA/46EA | 48.4 ± 17.1 | 7.4 ± 1.0 |
| | M | 256 | 156CTL/100SZ | 129AA/127EA | 44.6 ± 16.1 | 7.8 ± 0.9 |
| Hippocampus | F | 126 | 79CTL/47SZ | 82AA/44EA | 47.9 ± 16.7 | 7.64 ± 1.1 |
| | M | 268 | 182CTL/86SZ | 131AA/137EA | 44.4 ± 16.2 | 7.7 ± 1.0 |

## Materials and Methods

The research described herein complies with all relevant ethical regulations. All specimens used in this study were obtained with informed consent from the next kin under protocols No. 12-24 from the Department of Health and Mental Hygiene for the Office of the Chief Medical Examiner for the State of Maryland, and No. 20111080 for the Western Institutional Review Board for the Offices of the Chief Medical Examiner for Kalamazoo Michigan, University of North Dakota in Grand Forks North Dakota, and Santa Clara County California. Details of case selection, curation, diagnosis, and anatomical localization and dissection can be found in previous publications from our research group^12, 15^.

### BrainSeq Consortium RNA-sequencing data processing

We surveyed data from the BrainSeq Consortium^12, 15^ for caudate nucleus, DLPFC, and hippocampus, specifically: phenotype information; FASTQ files; single nucleotide polymorphism (SNP) array genotypes; region-specific covariates; and gene, exon, and exon-exon junction raw counts. For transcript estimated counts, we processed FASTQ files with Salmon (v1.1.0)^20^ using the reference transcriptome (GENCODE v25, GRCh38.p7) for annotation based alignment in parallel with GNU parallel^21^.

### BrainSeq Consortium imputation and genotype processing

We imputed genotypes as previously described^15^. Briefly, we first converted genotype positions from hg19 to hg38 with liftOver^22^. The Trans-Omics for Precision Medicine (TOPMed) imputation server^23–25^ was used for imputation of genotypes filtered for high quality (removing low-quality and rare variants) using the genotype data phased with the Haplotype Reference Consortium (HRC) reference panels (https://mathgen.stats.ox.ac.uk/impute/1000GP_Phase3.html). Genotypes were phased per chromosome using Eagle (version 2.4;^26^). We performed quality control with the McCarthy Tools (https://www.well.ox.ac.uk/~wrayner/tools/HRC-1000G-check-bim-v4.3.0.zip): specifically, we removed variants and samples with minor allele frequency (MAF) less than 0.01, missing call frequencies greater than 0.1, and Hardy-Weinberg equilibrium below p-value of 1e-10 using PLINK2 (v2.00a3LM)^27^. This resulted in 11474007 common variants.

For population stratification of samples, we performed multidimensional scaling (MDS) with PLINK version 1.9^28, 29^ on linkage disequilibrium (LD)-independent variants. The first component separated samples by ancestry as reported by the medical examiner’s office.

### Sample selection

We selected samples from the caudate nucleus, DLPFC, and hippocampus based on four inclusion criteria: 1) RiboZero RNA-sequencing library preparation, 2) age > 13 years, 3) ethnicity either African American or Caucasian American, and 4) TOPMed imputed genotypes available. This resulted in a total of 1170 samples from 504 unique individuals across the three brain regions for expression quantitative trait loci (eQTL) analysis. For expression-based analysis, we excluded individuals with age < 17 years resulting in a total of 1127 samples from 480 unique individuals across the three brain regions.

### Subject details

Of all 1170 samples used in the eQTL portion of this study, 399 were from the caudate nucleus, 377 from the DLPFC, and 394 from the hippocampus. Out of the 1170 samples, 126, 121, and 126 were female and 273, 256, 268 were male from the caudate nucleus, DLPFC, and hippocampus, respectively (**Table 1**). For the 1127 samples used in the expression analyses of this study, 393, 359, and 375 samples were located in the caudate nucleus, DLPFC, and hippocampus, respectively (**Table 2**). More information can be found in **Tables 1 and 2**.

**Table 2.**
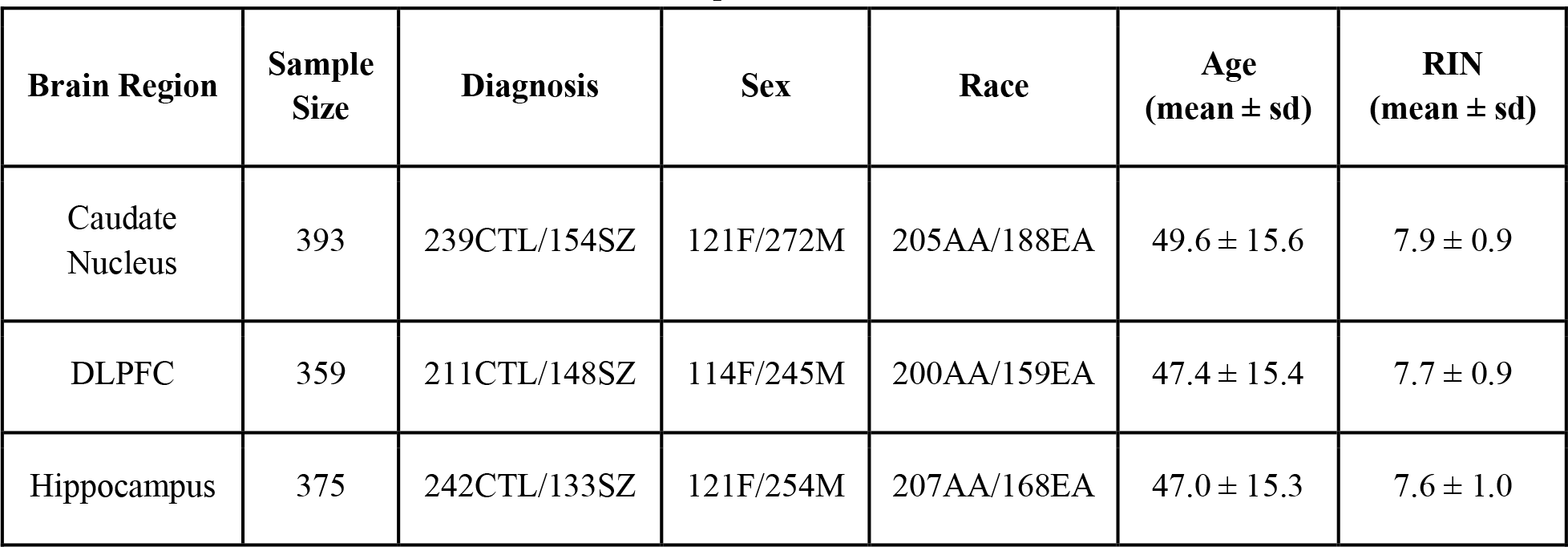
A sample breakdown of expression analysis for adult (age > 17) postmortem caudate nucleus, DLPFC, and hippocampus from the BrainSeq Consortium. Abbreviations: Neurotypical Control (CTL), Schizophrenia (SZ), Female (F), Male (M), African American (AA), European American (EA).

| Brain Region | Sample Size | Diagnosis | Sex | Race | Age (mean ± sd) | RIN (mean ± sd) |
| --- | --- | --- | --- | --- | --- | --- |
| Caudate Nucleus | 393 | 239CTL/154SZ | 121F/272M | 205AA/188EA | 49.6 ± 15.6 | 7.9 ± 0.9 |
| DLPFC | 359 | 211CTL/148SZ | 114F/245M | 200AA/159EA | 47.4 ± 15.4 | 7.7 ± 0.9 |
| Hippocampus | 375 | 242CTL/133SZ | 121F/254M | 207AA/168EA | 47.0 ± 15.3 | 7.6 ± 1.0 |

### Match gender phenotype to sex chromosomes

To match gender phenotype with sex chromosomes, we applied the sex imputation function (--check-sex) from PLINK (v1.9)^28, 29^. This compares sex assignments in the input dataset with those imputed from X-chromosome inbreeding coefficients. We used a Jupyter Notebook (v6.0.2) with the R kernel to compare reported gender with genotype imputed sex (F estimates). Here, we found all gender phenotypes matched sex chromosomes with F estimates for females below 0.22 and males above 0.9 (Fig. S1).

### Quality control and covariate exploration for sex

Observed expression measurements can be affected by biological and technical factors. To evaluate potential sources of confounding for expression or sex, we correlated biological (i.e., antipsychotics status at time of death, age at death, and global ancestry) and technical variables (i.e., RIN, mitochondria mapping rate, overall mapping rate, total gene assignment, and ERCC mapping rate) with gene expression as a function of sex (**Fig. S2**). For model covariates, we used variables that had a significant correlation (Bonferroni corrected p-value < 0.05) with gene expression for either sex in at least one brain region. To account for possible hidden effects on gene expression not captured by the above covariates, we also applied surrogate variable analysis^30, 31^. When we regressed out biological, technical, and hidden effects, we found this reduced all spurious correlations (**Fig. S3**).

### Expression normalization

For expression normalization, we constructed edgeR^32, 33^ objects for each brain region with raw counts and sample phenotype information. Next, we filtered out low expression counts using filterByExpr from edgeR, which keeps features above a minimum of 10 count-per-million (CPM) in 70% of the smallest group sample size (i.e., female individuals). Following this, we normalized library size using trimmed mean of M-values (TMM) before applying voom normalization^34, 35^ on four different linear models that examine: 1) sex (**Equation 1**), 2) interaction of brain region and sex (**Equation 2**), 3) interaction of sex and diagnosis (**Equation 3**), and 4) diagnosis subset by sex (**Equation 4**). Example covariates are diagnosis, age, brain region, ancestry (SNP PCs 1-3), RNA quality (RIN, mitochondria mapping rate, gene assignment rate, genome mapping rate, and rRNA mapping rate). For sex, interaction of sex and diagnosis, and diagnosis by sex analyses, we also corrected for any hidden variance via surrogate variable analysis.

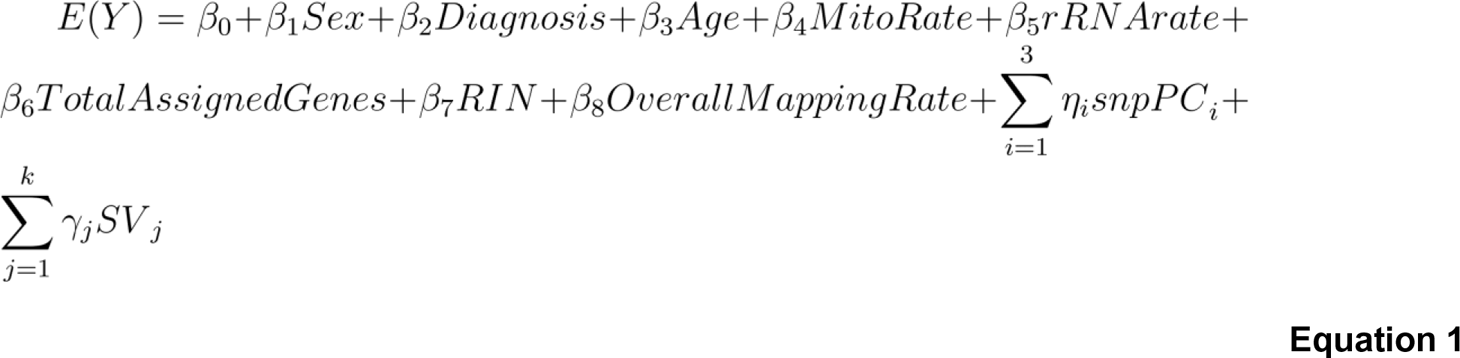

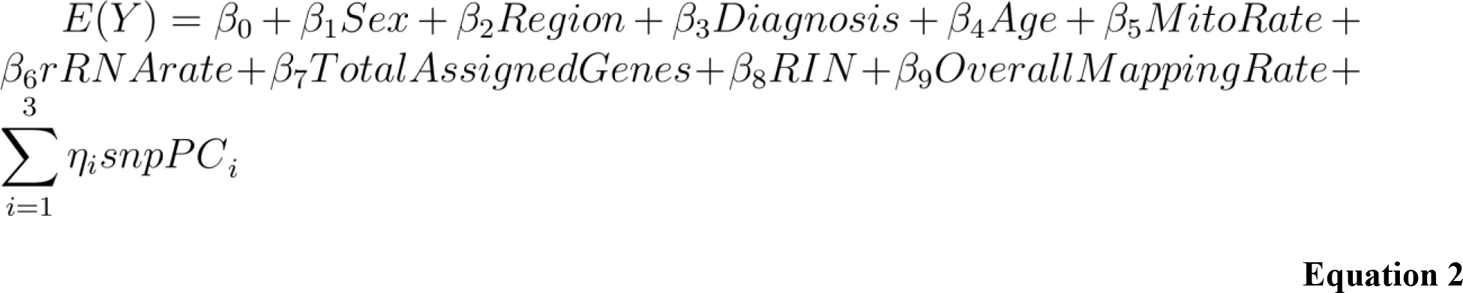

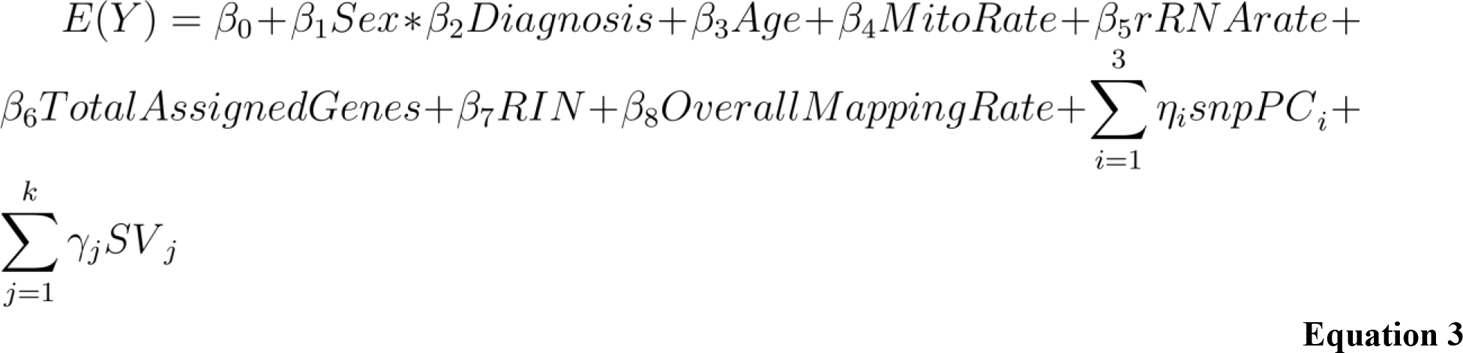

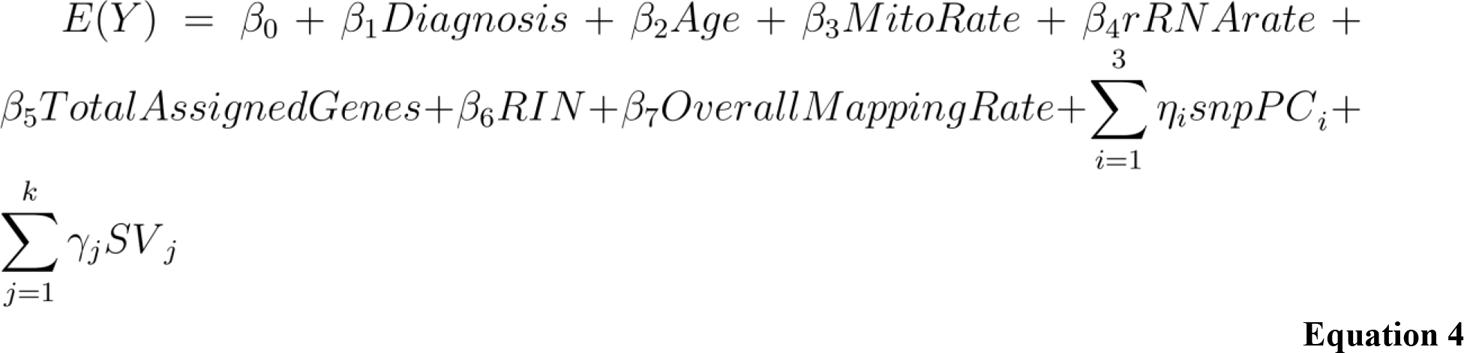

As there was significant overlap of individuals among the three brain regions examined, we used dream^36^ (differential expression for repeated measures) from variancePartition to correct for random effect of duplicate individuals across brain regions to assess potential significant interactions between brain region and sex (**Equation 2**). As such, we applied voom via dream with voomWithDreamWeights.

### Expression residualization

For residualized expression, we used voom-normalized expression and null models to regress out covariates as previously described^15^. After regressing out covariates, we applied a z-score transformation. Null models were created without variable(s) of interest to examine: 1) sex (**Equation 4**), 2) interaction of brain region and sex (**Equation 5**), 3) interaction of sex and diagnosis (**Equation 6**), and 4) diagnosis (**Equation 6**).

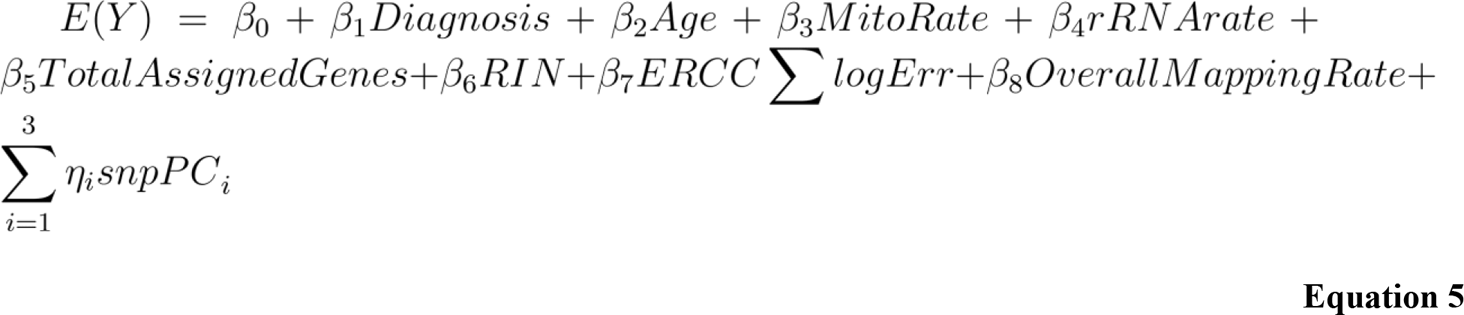

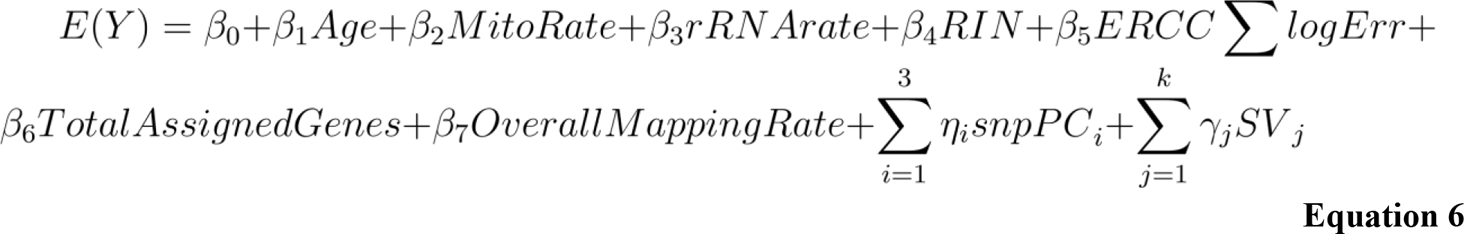

### Differential expression analysis

Following voom normalization, we fit four linear models (**Equations 1-4**) to examine: 1) sex, 2) interaction of sex and brain region, 3) interaction of sex and diagnosis, and 4) diagnosis subset by sex. With our fitted model, we identified differentially expressed features using the eBayes^37^ function from limma. Linear model fitting and differential expression calculation was completed in one step for interaction of sex and brain region with dream.

### Random forest dynamic recursive feature elimination

For autosomal sex prediction, we used dRFEtools^38^ to apply dynamic recursive feature elimination with random forest classification^39^. We set elimination rate to 10% and set 0.35 as the fraction of samples used for lowess smoothing. To reduce overfitting, we generated 10 sex-stratified folds for cross-validation with the *StratifedKFold* function from scikit-learn^39^. Model performance was measured using normalized mutual information, accuracy, and area under the receiver operating characteristic (ROC) curve with Out-of-Bag samples.

### Weighted correlation network analysis (WGCNA) analysis

We performed a signed network WGCNA^40^ analysis using residualized expression (**Equation 4**) to generate the co-expression network using: 1) all genes and 2) autosomal only genes in a single block by brain region. First, we filtered genes and outlier individuals with the WGCNA function *goodSamplesGenes*. We also filtered individuals with Z-normalized expression greater than 2.5. After evaluating power and network connectivity for each brain region we selected eight for soft power for both network constructions. Networks were constructed using *bicor* correlation and set a *deepSplit* of two for the caudate nucleus and hippocampus and three for the DLPFC. Additionally, *mergeCutHeight* was set to 0.15 and *minModuleSize* set to 50 for all brain regions and gene networks. The full (allosomal + autosomal) co-expression network was made using Pearson correlation values with 381, 349, 364 samples and 23488, 23039, and 22990 genes for the caudate nucleus, DLPFC, and hippocampus, respectively. The autosomal only co-expression network was made using Pearson correlation values with 381, 349, and 364 samples and 22649, 22219, and 22154 genes for the caudate nucleus, DLPFC, and hippocampus, respectively. Significant associations with sex were determined using a linear model and Pearson correlation between binary sex and module eigengenes.

### Sex-specific differential expression analysis for schizophrenia

To determine more stringent sex-specific differential expression features among the three brain regions using diagnosis subset by sex, we applied additional selection criteria following differential expression analysis. First, we removed any overlapping differentially expressed features. Following removal, we tested features for significant differences in residualized expression (**Equation 6**) for the opposite sex using Mann-Whitney U and removed significant features (p-value < 0.05).

### Subsampling analysis for male only schizophrenia differential expression

For subsampling of the BrainSeq Consortium brain region analysis, we randomly sampled male individuals using the female sample sizes (121, 114, and 121 for the caudate nucleus, DLPFC, and hippocampus) and performed differential expression analysis (**Equation 6**) for schizophrenia. We performed this 1000 times.

### Functional gene term enrichment analysis

For functional enrichment analysis, we used g:Profiler via R (version 4.0.2) with the gprofiler2 package^41^. We used a variety of curated gene sets, including: the database from Gene Ontology (GO); human disease phenotypes from Human Phenotype Ontology (HP); regulatory motifs for miRNA targets with miRTarBase and transcription factors with TRANSFAC; pathways from Kyoto Encyclopedia of Genes and Genomes (KEGG), Reactome, and WikiPathways (WP); and protein databases from Human Protein Atlas (HPA) and the comprehensive resource of mammalian (CORUM) protein complexes.

The GO database included molecular functions (MF), cellular components (CC), and biological processes (BP). We corrected for multiple testing using g:Profiler algorithm g:SCS with alpha set to 5%. We generated Manhattan-like plots using the gprofiler2 package with the *gostplot* function. To measure GO term elements semantic similarity across brain regions, we used R package GOSemSim^42^ with the Wang method^43^ and Best-Match Average strategy.

For gene-term enrichment analysis for WGCNA modules, we used GOATOOLS Python package^44^ with the GO database and hypergeometric tests for enrichment and depletion as previously described^15^.

Specifically, we converted gencode IDs to Entrez IDs using pybiomart (https://github.com/jrderuiter/pybiomart). With Entrez IDs, we applied enrichment analysis per module. Multiple testing correction was done using Benjamini-Hochberg FDR method.

### X-chromosome inactivation (XCI) enrichment analysis

For XCI enrichment analysis, we downloaded the XCI status annotation from^45^. We accessed enrichment of sex bias for XCI status using Fisher’s exact test with the known XCI categories, including 631 genes defined as escape (n=99), variable escape (n=101), or inactive (n=431). We corrected for multiple testing with Bonferroni.

### Dosage compensation

Relative X expression (RXE) was determined as previously described^46^ with slight modifications; specifically, we used transcripts per million (TPM). We generated TPM using the mean of the read insert size for effective length (**Equation 7**). Mean insert size was determined with the Picard tool CollectInsertSizeMetrics (http://broadinstitute.github.io/picard/). We dropped any genes with effective length less than or equal to one. Following TPM calculation, we performed a log2 transformation (**Equation 8**). Next, we filtered low-expressing genes present in at least 20% of samples. To compute RXE, we calculated the differences in the mean chromosome-wide log2 TPM expression with X-chromosome log2 TPM expression (**Equation 9**).

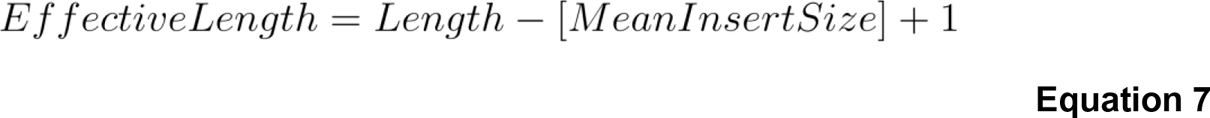

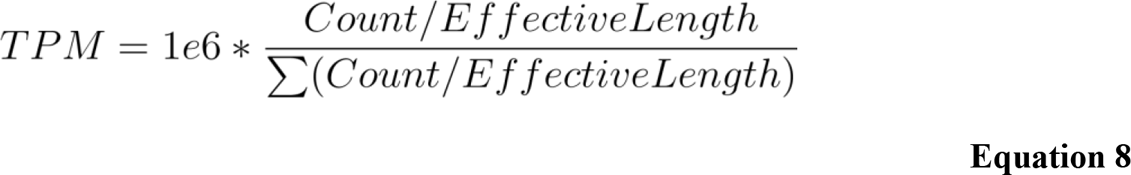

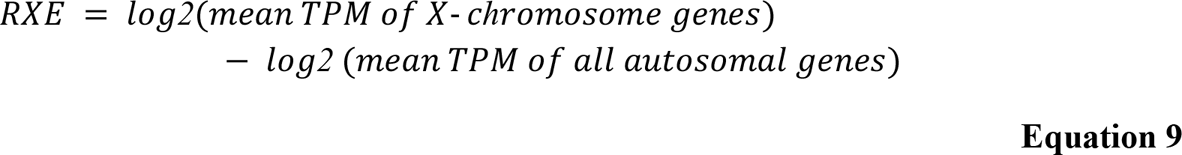

### Sex interacting eQTL analysis in *cis* and region specificity

To identify sex-interacting cis-eQTL (si-eQTL) across the caudate nucleus, DLPFC, and hippocampus, we first separated out female and male individuals and, using PLINK2, excluded variants with MAF < 0.05 and variants with less than one allele by sex. To generate a common list, we overlapped these filtered variants, resulting in a total of 6,816,103 SNPs. Following SNP filtering, we performed eQTL analysis using FastQTL^47^ as previously described^15, 19^ with modification for a sex-interaction model. We filtered low expression using the GTEx python script, eqtl_prepare_expression.py, with modification for processing transcripts, exons, and junctions. This retained features with expression estimates > 0.1 TPM in at least 20% of samples and aligned read count of six or more per brain region. Following low expression filtering, we performed TMM normalization on filtered counts using the GTEx python script, rnaseqnorm.py (https://github.com/broadinstitute/gtex-pipeline/tree/master/qtl/src/rnaseqnorm.py). Following normalization, we implemented FastQTL using an interaction linear regression model. To do this, we performed three major steps: 1) we adjusted expression for covariates (i.e., diagnosis, population stratification [SNP PCs 1-3], and expression PCs specific to brain region and feature); 2) selected *cis*-SNP using a mapping window of 0.5 Mb within the transcriptional start site (TSS) of each feature; and 3) filtered SNPs based on an interaction MAF ≥ 0.05 and the minor allele present in at least 10 samples.

To assess sharing across brain regions and to increase our power to detect sex-interacting eQTL effects, we used multivariate adaptive shrinkage in R (mashr; v0.2.57)^48^ as previously described^15^. mashr uses an empirical Bayes approach to learn patterns of similarity among conditions (e.g., brain regions) then leverage these prior patterns to improve accuracy of effect size estimates. We obtained effect sizes and standard errors for these effect sizes from the FastQTL interaction model results. To account for correlations among measurements across brain regions (i.e., overlapping sample donors), we used the *estimate_null_correlation_simple* function to specify a correlation structure prior to fitting the mash model. The mash model included both the canonical covariance matrices and data-driven covariance matrices learned from our data. We defined the data-driven covariance matrices as the top three PCs from the principal components analysis (PCA) performed on the significant signals (i.e., most significant nominal p-values by brain region). To learn the mixture weights and scaling for the si-eQTL effects, we initial fit the mash model with a random set (i.e., unbiased representation of the results) of the FastQTL interaction model results (i.e., 5% for gene-SNP pairs and 1% for transcript-, exon-, and junction-SNP pairs). We next fitted these mixture weights and scaling to all of the si-eQTL results in chunks. We extracted posterior summaries and measures of significance (i.e., local false sign rate [lfsr]). We considered si-eQTL significant if the lfsr < 0.05.

### Schizophrenia risk GWAS association

We downloaded the latest schizophrenia GWAS summary statistics with index and high-quality imputation SNPs as determined by Psychiatric Genomics Consortium (PGC3)^49^. Following download, we selected and converted PGC3 GWAS SNPs associated with BrainSeq Consortium SNPs as previously described^15^; specifically, we converted GWAS SNPs from hg19 to hg38 using pyliftover, merged them with BrainSeq Consortium SNPs on hg38 coordinates, and matched alleles.

### Fine mapping and colocalization

We performed fine mapping and colocalization with gene level si-eQTL for the caudate nucleus, DLPFC, and hippocampus as previously described^15, 50^ with slight modification for priors. Briefly, we estimated priors from the FastQTL nominal results with torus^51^. Following estimation of priors, we implemented dap-g^52, 53^ to generate posterior inclusion probabilities (pip) that provide an estimate of the probability of a variant being causal for downstream colocalization with fastENLOC^54, 55^. We applied fastENLOC with schizophrenia GWAS (PGC3)^49^.

We generated colocalization plots using eQTpLot^56^ and eQTL results from sex-only analysis. Specifically, we applied gene-level *cis*-eQTL analysis to female and male individuals separately with FastQTL as described above without modification for sex interaction^15^. We used a genebody window of 0.5Mb, MAF ≥ 0.01, and confounders generated from the *Sex-interacting eQTL analysis in* cis *and region specificity*. We determined significance for the most highly associated variant per gene using empirical p-values based on beta-distribution fitted with an adaptive permutation (1000 to 10000). These p-values were corrected for multiple testing across genes using Storey’s q-value. For each brain region, we applied eQTpLot using sex-specific nominal and permutation results (**Data S1**) for each significant colocalized genes identified (regional colocalization probability [RCP] > 0.5).

### General replication analysis

We downloaded differential expression results for sex differences from the supplemental materials for Trabzuni et al., Mayne et al., and Gershoni and Pietrokovski^57–59^. For sex differences in schizophrenia replication, we downloaded Qin et al. results^6^. For sex-interacting eQTL, we downloaded results from Trabzuni et al., Yao et al., Kukurba et al., and Shen et al.^16–18, 57^.

For CommonMind Consortium replication of differential expression analysis, we downloaded differential expression results for sex differences from Hoffman et al.^7^, as well as normalized expression from Synapse (syn18103849). For dosage compensation replication, we calculated TPM using a mean insert size of 200. We computed relative X expression as described above (**Dosage compensation**).

We downloaded gene TPM from the GTEx v8 portal (https://www.gtexportal.org/home/datasets), as well as sample phenotype information. Relative X expression was computed as described above (**Dosage compensation**).

### Graphics

We generated Venn diagrams with matplotlib_venn (v0.11.5) Python (version 3.8.6) package. We generated UpSet plots in R using ComplexHeatmap^60^ (v2.6.2). Unless otherwise stated, we generated boxplots and scatterplots in R using ggpubr (v0.4.0). We generated enrichment dotplots, enrichment heatmaps, and gene term enrichment plots using ggplot2^61^. To generate circos plots, we used circlize^62^ (v0.4.11) and ComplexHeatmap utilities in R. We used plotnine, a Python implementation of ggplot2, to generate enrichment heatmaps comparing public datasets with BrainSeq Consortium analysis, RXE scatter plots, and sex-interacting eQTL boxplots. To generate rank-rank hypergeometric overlap (RRHO), we used the RRHO^63^ (v1.28.0) and lattice packages in R.

### Code availability

All code and Jupyter Notebooks are available through GitHub at https://github.com/LieberInstitute/sex_differences_sz with more detail.

## Results

### Sex-specific expression across the caudate nucleus, DLPFC, and hippocampus

We first explored sex differences in the brain of the 480 unique individuals (caudate nucleus [n=393], DLPFC [n=359], and hippocampus [n=375]) by performing differential expression of sex after adjusting for diagnosis, age, ancestry (SNP PCs 1-3), RNA quality, and hidden variances (**Equation 1** and **Table 2**) using the BrainSeq Consortium dataset^12, 15^. We observed 878 unique, differentially expressed genes (DEGs; false discovery rate [FDR] < 0.05; **Fig. 1A**) between the sexes across the caudate nucleus (n=380 DEGs), DLPFC (n=573), and hippocampus (n=105). Of these 878 unique DEGs, the sex chromosomes showed the most significant sex-biased expression (**Data S2**). Interestingly, most sex-associated DEGs for the caudate nucleus and DLPFC were autosomal (**Table S1**). When we expanded our analysis to transcripts, exons, and exon-exon junctions, we observed a similar pattern of majority autosomal genes; however, the most statistically significant differentially expressed features were located on sex chromosomes (**Table S1** and **Data S2**).

**Fig. 1.**
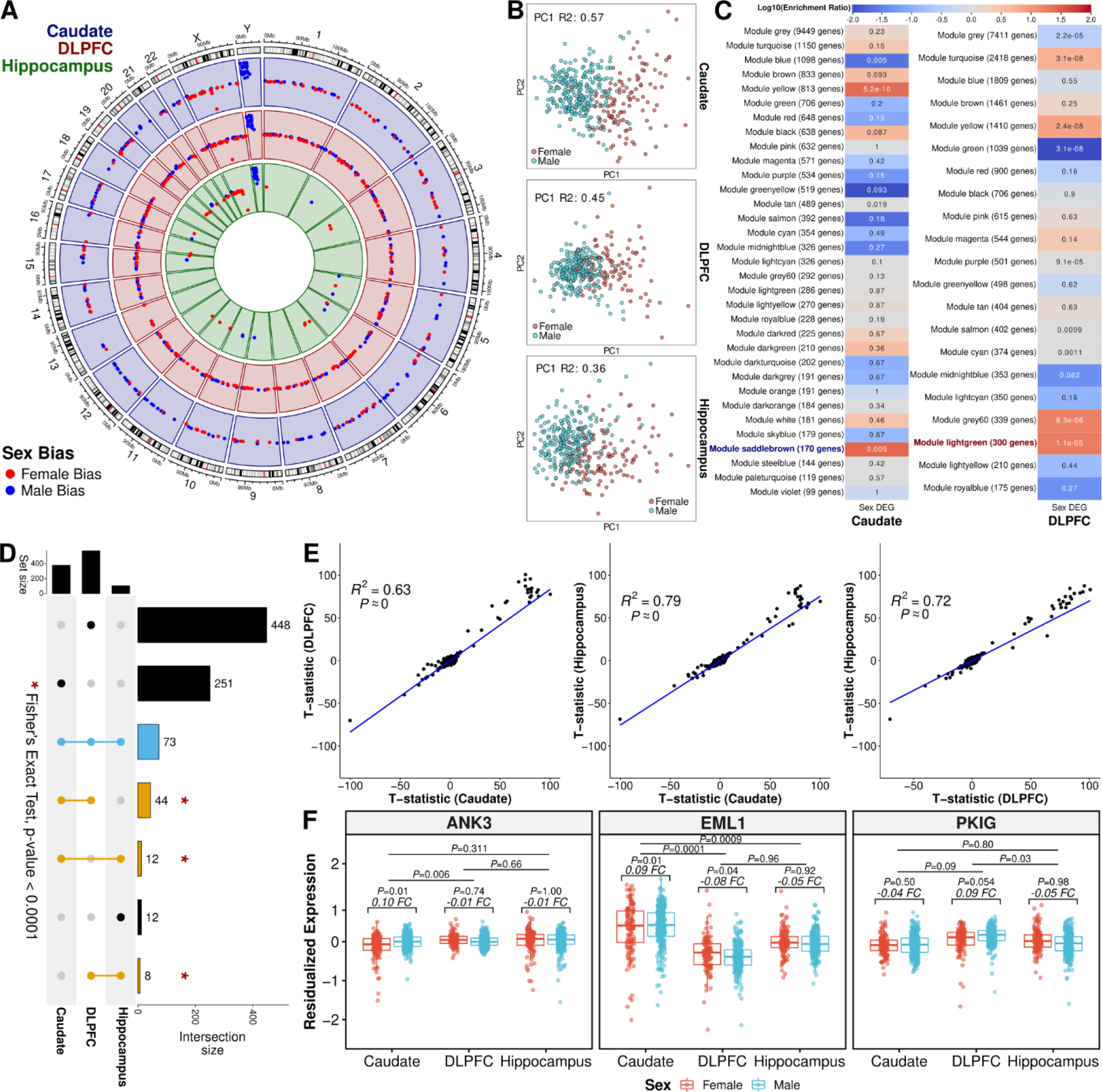
Sex-biased expression across the caudate nucleus, DLPFC, and hippocampus. **A.** Circos plot showing significant DEGs for the caudate nucleus (blue; n=393; 121 female and 272 male), DLPFC (red; n=359; 114 female and 245 male), and hippocampus (green; n=375; 121 female and 254 male) across all chromosomes. Female bias (up-regulated in female individuals) in red, and male bias (up-regulated in male individuals) in blue. **B.** Scatterplots of the estimated proportion of expression variance explained by sex within the 100 most significant autosomal DEGs (i.e., adjusted p-value) for the caudate nucleus (DEGs, n=60), DLPFC (DEGs, n=25), and hippocampus (DEGs, n=42). **C.** Enrichment heatmap of sex DEGs for WGCNA autosomal-only modules for caudate nucleus and DLPFC. Tiles annotated with FDR, two-sided Fisher’s exact test p-value corrected with Benjamini-Hochberg procedure and color associated with depletion (blue) and enrichment (red) for sex-biased DEGs. Modules significantly correlated (two-sided, Pearson, p-value < 0.05) with female individuals highlighted in blue or male individuals in red. **D.** UpSet plot showing overlap of differentially expressed genes across the caudate nucleus, DLPFC, and hippocampus. **\*** Indicating p-value < 0.0001 for two-sided, Fisher’s exact test. **E.** Scatter plot of t-statistics for all genes tested showing concordant positive directionality with significant two-sided, Spearman correlation (R^2^) of all genes. **F.** Boxplots of *ANK3*, *EML1*, and *PKIG* residualized expression showing an interaction between sex and brain region. FC= fold change log2 (male / female). Female individuals in red and male individuals in blue. Adjusted P-value (*P*) annotation using dream^36^ (default of Satterthwaite approximation) generated statistics annotation.

To evaluate the functions associated with sexually dimorphic gene expression, we performed functional gene term enrichment analysis on DEGs by bias status (all, female-bias [up-regulated in females], or male-bias [up-regulated in males]) separately per brain region. We observed significant enrichment (hypergeometric, adj. p-value < 0.05) of several ontology terms for each brain region (**Data S3**). The caudate nucleus DEGs showed enrichment of terms related to: histone demethylation; X-linked inheritance; the structural integrity of the extracellular matrix; and DNA structure including desmosome and nucleosome (**Fig. S4A**). When we separated the DEGs by direction of effect, female-bias genes were significantly enriched in DNA structure and histone deacetylation pathways. In contrast, male-biased genes were enriched in neuronal function and cyclic nucleotide metabolic processes (**Fig. S4A**). In the DLPFC, DEGs showed enriched terms related to transcription factor motifs (**Fig. S4B**). Female-bias genes in the DLPFC were enriched for cellular structure, whereas male-biased genes were enriched in metabolic related pathways (**Fig. S4B**). The hippocampal DEGs were enriched in translational activity and histone demethylase activity similar to the caudate (**Fig. S4C**). Interestingly, only the hippocampal DEGs showed enrichment for synaptic localization (**Data S3**), implicating inhibitory and excitatory synapses as well as transcriptional activity differences between sex in the hippocampus. Female-biased genes were enriched in translational activity and aggressive behavior terms (**Data S3**).

To further understand the sex differences in the brain, we used WGCNA (weighted gene correlation network analysis) to identify co-expression modules associated with sex across the caudate nucleus, DLPFC, and hippocampus. Here, we found four modules significantly associated with sex (linear regression, p-value < 0.05) for the caudate nucleus (one module; R^2^ = 0.02) and the DLPFC (three modules; R^2^ > 0.01). For the caudate module, we found significant enrichment (hypergeometric test, FDR < 0.05) for terms related to the response to hypoxia, embryonic development, and angiogenesis (**Data S4**). For the three DLPFC modules, we found significant enrichment (hypergeometric test, FDR < 0.05) for terms related to mitochondrial function and chromatin remodeling (**Data S4**). Interestingly, hippocampus co-expression modules showed no association with sex (**Data S4**).

### Autosomes influence sex differences in the brain

With many autosomal sex-biased DEGs, we asked to what degree sex-biased autosomal genes contribute to sex differences in the brain. We separated allosomal and autosomal DEGs and performed principal component analysis to assess explained variance of these DEGs across the brain. While the first principal component of all allosome DEGs explained greater than 89% variance across brain regions, the autosomal DEGs also showed significant association with sex (**Fig. S5**). Interestingly, within the 100 most significant autosomal DEGs for the caudate nucleus and DLPFC, explained variances increased drastically to 57% and 45% compared to all autosomal DEGs, respectively (**Fig. 1B** and **Fig. S5**). As it appeared that a small subset of these autosomal DEGs explained a large proportion of expression variances between the sexes, we formally tested this using dynamic recursive feature elimination (dRFE)^64^. To this end, we applied random forest classification using 10-fold, sex-stratified, cross-validation with dRFE and found a median of 48 genes (48, 48, and 48.5 for caudate nucleus, DLPFC, and hippocampus respectively) with greater than 85% accuracy for sex classification (**Table S2** and **Fig. S6**). Interestingly, only 15 of 102 (14.7%) unique autosomal predictive genes were shared across brain regions with a majority (75 of 102 [73.5%]) showing brain-region specificity (**Fig. S7**). These results indicate that a subset of autosomal genes significantly contributes to sexually dimorphic gene expression in the brain.

To further understand the autosomal contribution to sex differences in the brain, we also identified co-expression modules associated with sex using only autosomal co-expression modules. Here, we found one module significantly associated with sex (linear regression, p-value < 0.05) for the caudate nucleus (R^2^ = 0.02) and DLPFC (R^2^ = 0.01), both of which showed significant enrichment (Fisher’s exact test, FDR < 0.05) for sex-biased DEGs (**Fig. 1C**). When we examined the functional relevance of these modules, we found significant enrichment (hypergeometric test, FDR < 0.05) for terms related to the modules, including allosomes (i.e., angiogenesis, blood-brain barrier, and placenta development for the caudate nucleus and proteasome function and activity, GTPase activity, and protein binding for DLPFC; **Data S4**). As with the full gene network, we did not find any modules showing an association with sex in the hippocampus (**Data S4**). These results suggest that autosomal modules associated with sex are involved in diverse biological processes in the brain.

### Brain region interaction with sex

To understand the regional specificity of sex DEGs, we compared DEGs from each brain region. We observed a significant enrichment of shared DEGs across the three brain regions (Fisher’s exact test, p-value < 0.05; **Fig. 1D**) with the majority (51 DEGs, 70%) on sex chromosomes. For replication analysis, we compared the DEGs with previous sex differences analysis in the brain^7, 57–59^ and found greater than 66% of DEGs were significantly differentially expressed in all brain regions except for the GTEx cerebellum and anterior cingulate cortex (**Fig. S8**) with a concordant direction of effect between BrainSeq Consortium and GTEx brain regions (Fisher’s exact test, p-value < 0.01). For a more in-depth comparison, we examined the sex differences found using the CommonMind Consortium (CMC) DLPFC^7^. We also discovered a large number of DEGs on sex chromosomes (39 of 51 [76.5%] and 41 of 54 [75.9%] for the NIMH HBCC and MSSM-Penn-Pitt cohorts, respectively) similar to our BrainSeq Consortium analysis. Additionally, we observed significant pairwise enrichment of these CMC DEGs with our BrainSeq Consortium DEGs across brain regions (Fisher’s exact test, p-value < 0.01; **Fig. S9**). Altogether, this suggests that X- and Y-linked genes drive brain-wide sex expression differences and autosomal genes drive brain region-specific differences. Additionally, autosomal DEGs were less likely to replicate in different datasets.

Interestingly, we found that all genes regardless of significant association with sex showed a significant positive correlation for the direction of effect between pairwise comparisons of the three brain regions (Pearson, R^2^ > 0.63, p-value < 0.01; **Fig. 1E**). At significant levels (DEGs, adjusted p-value < 0.05), these pairwise correlations dramatically increased (Pearson, R^2^ > 0.96, p-value < 0.01; **Fig. S10A**) with only one gene (*EML1*) showing region-specificity – male-bias in the caudate nucleus and female-bias in the DLPFC (**Fig. 1F**). Expanded analysis of transcripts, exons, and exon-exon junctions displayed a similar pattern with all shared, differentially expressed features (DE, FDR < 0.05) having significant concordant direction and significant positive correlation among brain regions (Pearson, R^2^ > 0.88, p-value < 0.01; **Fig. S10B-D**). Moreover, at significant levels (FDR < 0.05) all directions agreed between the CMC DLPFC NIMH HBCC cohort and the BrainSeq Consortium brain regions with a significant positive correlation (Pearson; R^2^ > 0.95 for all pairwise comparisons; p-value < 0.01; **Fig. S11A**), and all but one DEG agreed with the CMC DLPFC MSSM-Penn-Pitt cohort with a significant positive correlation (Pearson; R^2^ > 0.88 for all pairwise comparisons; p-value < 0.01; **Fig. S11B**). In summary, the direction of change for sexually dimorphic genes are generally shared across multiple brain regions and independent datasets.

We next evaluated the degree of sex-bias among brain regions formally with an interaction model for sex and brain region. Here, we found extensive interactions, particularly in the caudate nucleus versus DLPFC (**Table S3** and **Data S5;** FDR < 0.05, caudate nucleus vs DLPFC, n=668; caudate nucleus vs hippocampus, n=23; and DLPFC vs hippocampus, n=26), including *ANK3*, *EML1*, and *PKIG* (**Fig. 1F**). Furthermore, we found similar levels of differentially expressed features on the transcript-, exon-, and junction-level analysis (**Table S3**).

Of note, *EML1* showed an interaction of sex with the caudate nucleus-hippocampus and caudate nucleus-DLPFC comparisons, confirming the previously detected discordance between the caudate nucleus and DLPFC. In the hippocampus, *EML1* showed a non-significant trend in the same direction as DLPFC. Disruption of *EML1* is associated with subcortical heterotopia in the mouse brain^65, 66^. Interestingly, *ANK3*, a marker for early life stress and a risk gene for neurodevelopmental and neuropsychiatric disorders – including schizophrenia^67^ – showed a significant interaction between sex in the caudate nucleus with male-biased in the caudate nucleus on the gene, exon, and junction levels. Altogether, this analysis further highlights the transcriptional sex differences in the caudate compared with the DLPFC or hippocampus.

### X-chromosome inactivation and dosage compensation in the brain

As 70% of the brain-wide, sexually dimorphic genes are located on sex chromosomes, we next evaluated the dosage of X-linked genes compared to autosomes. In order to equalize the dosage of X-linked genes between XX females and XY males, female mammals epigenetically silence one X chromosome in a process called X-chromosome inactivation. X-chromosome inactivation is a chromosome-wide process where the majority of X-linked genes are nearly completely silenced, and a minority of X-linked genes either escape X inaction or show variable X inactivation. When we examined the DEGs by brain region for X-linked gene dosage, we found the majority of DEGs were enriched for X-chromosome inactivation (XCI) escape genes (**Fig. S12**, Fisher’s exact test, Bonferroni < 0.01; **Fig. 2A**), reflecting dosage compensation for the majority of X-linked genes subject to XCI as seen in previous studies^45, 68^.

**Fig. 2.**
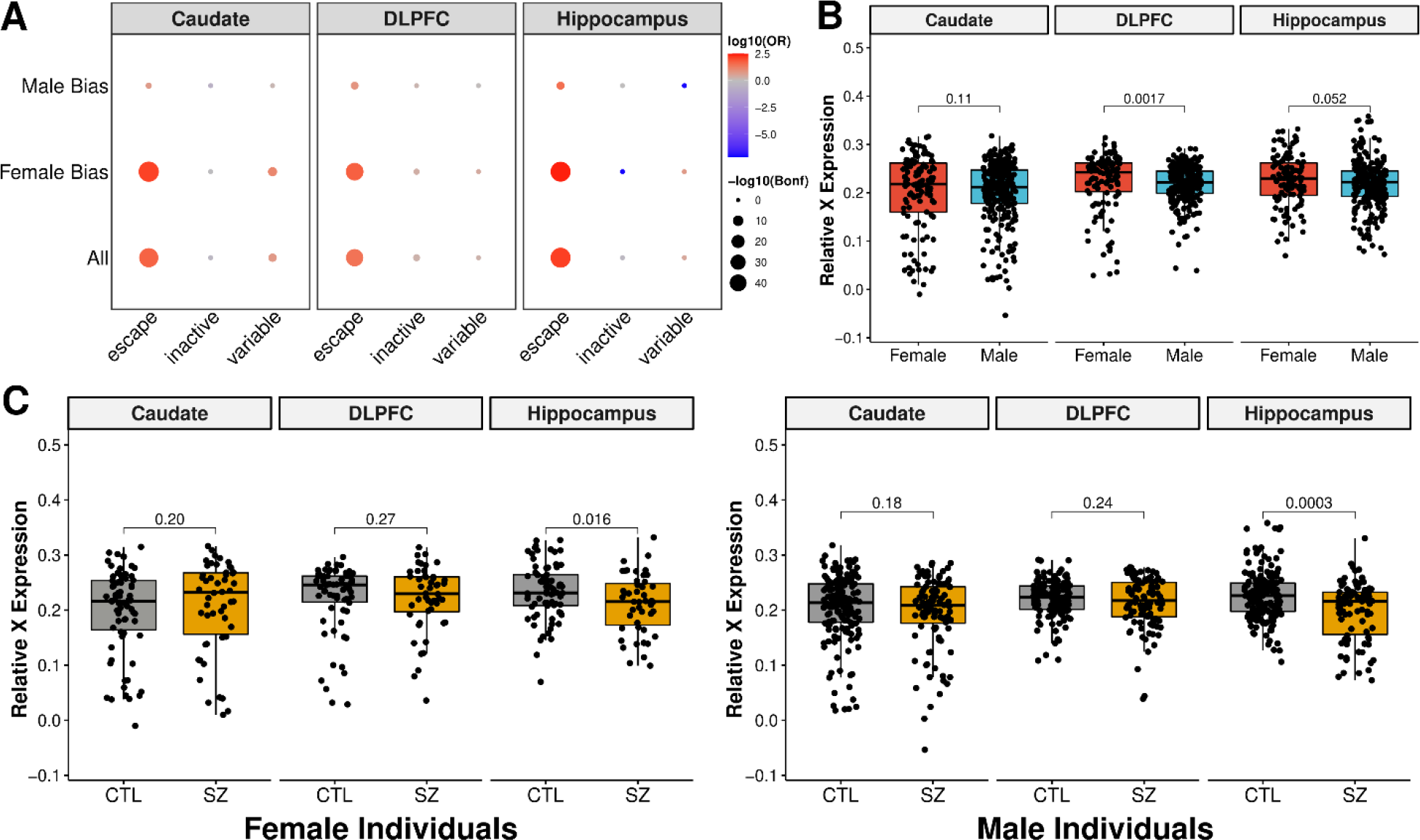
X-linked gene expression and dosage compensation observed across the caudate nucleus, DLPFC, and hippocampus. **A.** Enrichment of significant sex-biased genes relative to genes known to escape XCI across the caudate nucleus (n=393; 121 female and 272 male), DLPFC (n=359; 114 female and 245 male), and hippocampus (n=375; 121 female and 254 male). Dotplot of enrichment (two-sided, Fisher’s exact test) of DEGs for X-chromosome inactivation (XCI) genes by brain region separated by male bias (up-regulated in male individuals), female bias (up-regulated in female individuals), and all DEGs. Size of dots denotes -log10 of Bonferroni corrected p-values. Color relates to log10 odds ratio (OR) with depletion in blue and enrichment in red. **B.** Boxplot showing RXE comparison between female (red) and male (blue) individuals with two-sided, Mann-Whitney U p-values annotated. **C.** Boxplot showing significant differences between neurotypical control (CTL; gray) and schizophrenic (SZ; gold) individuals in the caudate nucleus, DLPFC, and hippocampus for female (left) and male (right) individuals with two-sided, Mann-Whitney U p-values annotated.

For all three brain regions, we found XCI escape genes were significantly enriched within the female-biased DEGs, whereas male-biased DEGs, surprisingly, were significantly enriched for genes escaping XCI in DLPFC and hippocampus, but not the caudate nucleus (Fisher’s exact test, Bonferroni < 0.01; **Fig. 2A**). These male-biased escaping XCI genes in the DLPFC and hippocampus were associated with genes (*PLCXD1*, *CD99*, *ASMTL* and *GTPBP6*) located on the PAR (pseudoautosomal regions) of both X and Y chromosomes (**Data S6**). Additionally, we found the most male-biased XCI-annotated genes in the DLPFC (n = 18, 3, and 1 DEGs for DLPFC, caudate nucleus, and hippocampus, respectively), which were mostly annotated as inactive XCI genes (**Data S6**). In contrast, we only found enrichment of variable XCI genes in the caudate nucleus (Fisher’s exact test, Bonferroni < 0.01). Altogether, XCI escaping genes demonstrated higher expression in female individuals across brain regions, suggesting sex differences shared across brain regions are associated with well documented XCI escaping genes for females.

Next, we evaluated differences in chromosome-wide dosage by comparing the relative X-chromosome expression (RXE) to autosomes (**Fig. 2B** and **Fig. S13**). Interestingly, we observed a significant decrease of RXE in male individuals only in the DLPFC (Mann-Whitney U, p-value = 0.0017), demonstrating region-specific dosage compensation. We also observed a similar trend of decreased RXE in the CMC DLPFC MSSM-Penn-Pitt cohort (Mann-Whitney U, p-value = 0.07; **Fig. S14A**), but not the GTEx frontal cortex (**Fig. S15**). Even so, the large RXE variation across the 13 GTEx brain regions demonstrated region-specific dosage compensation (**Fig. S15**).

As we found differences in the DLPFC between sexes, we next asked if this might be due to individuals with schizophrenia. Interestingly, we found decreased RXE in the hippocampus of ill individuals for both sexes (Mann-Whitney U, p-value < 0.01; **Fig. 2C**) but not in the caudate nucleus, DLPFC, or CMC DLPFC (**Fig. S14B**). However, there was no significant interaction between sex and diagnosis status for any brain region for RXE. These results demonstrate slight differences between X-chromosome dosage in the hippocampus of individuals with schizophrenia.

### Interaction of schizophrenia and sex in the brain

After investigating sex differences in the brain without consideration of diagnosis in 480 unique individuals (caudate nucleus [n=393], DLPFC [n=359], and hippocampus [n=375]), we next identified statistically significant differentially expressed features (FDR < 0.05) with respect to sex differences and diagnosis through an interaction model. No genes, transcripts, or exons were significant by this interaction model, similar to a previous study^7^. On the junction level, 15 novel junctions demonstrated a significant (FDR < 0.05) interaction between sex and diagnosis across the caudate nucleus (two junctions, **Fig. S16**), DLPFC (seven junctions, **Fig. S17**), and hippocampus (six junctions, **Fig. S18**). In the hippocampus, one of the six unannotated junctions (chr18:31592940-31592993[+]) was located within *TTR* (Transthyretin), a thyroid hormone-binding protein. Thyroid hormone deficiency in early gestation is linked to increased odds of schizophrenia^69^.

Larger sample sizes are needed to have effective statistical power to identify interactions compared to main effects. As such, we also examined differential expression for schizophrenic female and male individuals separately across the caudate nucleus, DLPFC, and hippocampus using rank-rank hypergeometric overlap (RRHO) analysis to increase our power of detecting transcriptional changes. Here, we found schizophrenia-related transcriptional signatures varied dramatically depending on sex and brain region (**Data S7** and **Table S4**). Specifically, female schizophrenia transcriptional signatures showed the strongest pattern of sharing between the caudate nucleus and the DLPFC (**Fig. 3A**), while males showed the strongest pattern of sharing between DLPFC and hippocampus (**Fig. 3B**), similar to ours and others’ previous schizophrenia analysis adjusted for sex^12, 15^. Altogether, sex-adjusted schizophrenia analysis largely reflects male transcriptional changes likely due to larger male sample sizes.

**Fig. 3.**
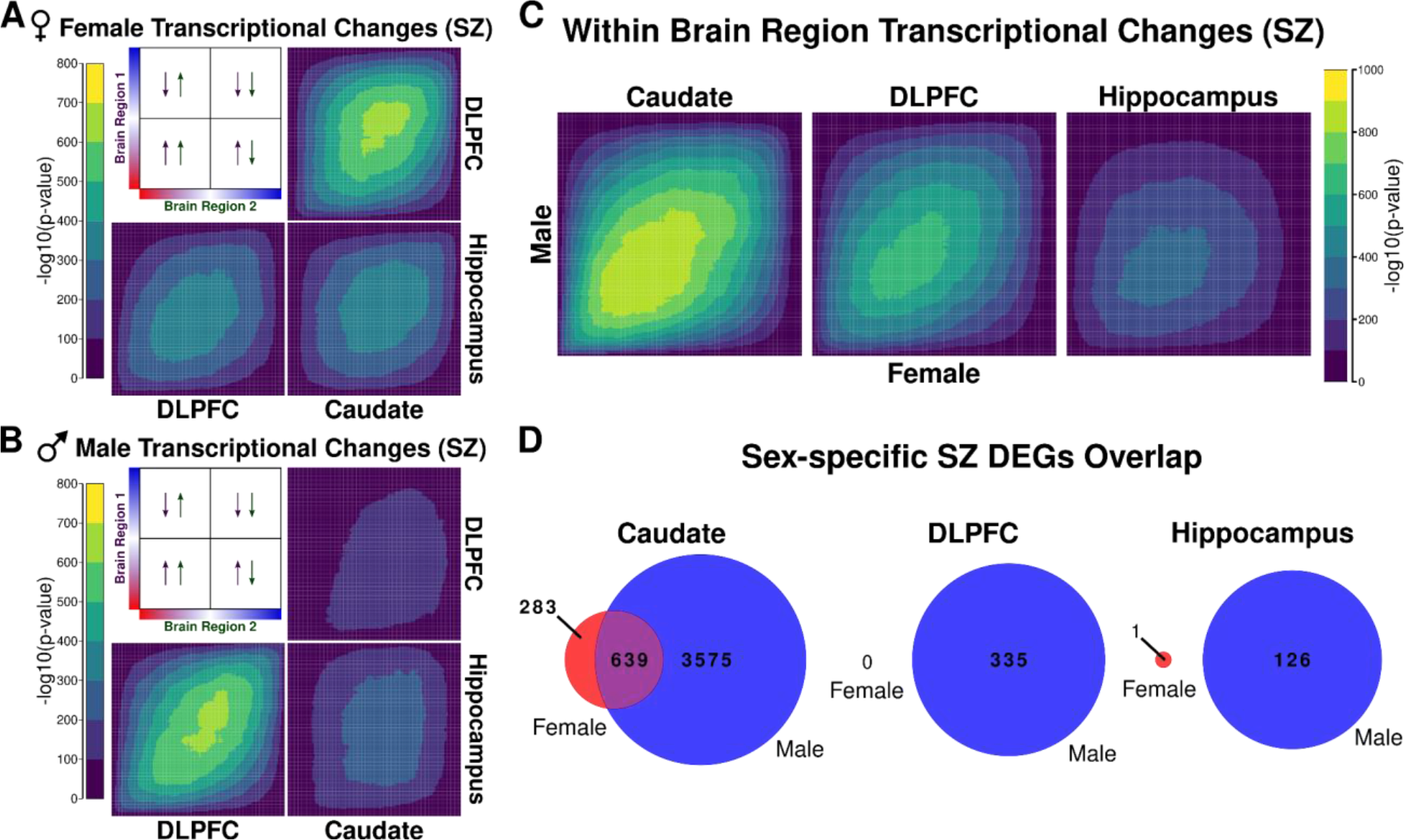
Transcriptional changes for schizophrenia shared between sexes within brain regions. RRHO (rank-rank hypergeometric overlap) maps comparing schizophrenia transcriptional changes for all genes between brain region pairs in **A.** females and **B.** males. The panel presents the overlap relationship between two brain regions. The color bars represent the degree of significance [-log10(p-value)] of overlap between two brain regions. Arrows show the direction of effect for schizophrenia (up- or down-regulated in schizophrenia) by brain region. **C.** RRHO map comparing female and male schizophrenia transcriptional changes within brain regions for all genes. The color bar represents the degree of significance [-log10(p-value)] of the overlap between the sexes. **D.** Venn diagram showing overlap within brain regions for sex-stratified schizophrenia DEGs (female in red and male in blue; FDR < 0.05). Female-specific schizophrenia DEGs in red, male-specific schizophrenia DEGs in blue, and schizophrenia DEGs shared between female- and male-specific schizophrenia analyses in purple. SZ: schizophrenia.

Next, we examined female and male transcriptional changes for schizophrenia within individual brain regions. Here, we found the strongest shared signature within the caudate nucleus with very little observable overlap for the DLPFC and hippocampus (**Fig. 3C**). This was further supported when we examined DEGs (FDR < 0.05) and found large overlap for the caudate nucleus but little to no overlap between females and males for the DLPFC and hippocampus (**Fig. 3D**). Similar trends occurred for transcripts, exons, and junctions (**Data S7** and **Fig. S19**).

### Sex-specific schizophrenia expression in the brain

To further examine female-specific schizophrenia DEGs, we removed any female-specific DEGs that were significantly different (Mann-Whitney U, p-value < 0.05) with residualized expression for male-only individuals. The single hippocampal DEG and 159 DEGs from the caudate nucleus were filtered out because they showed significant residualized expression for male schizophrenic individuals (Mann-Whitney U, p-value < 0.05) resulting in a total of 124 female-specific schizophrenia DEGs in the caudate nucleus (**Table S4**). Gene term enrichment analysis revealed enrichment in DNA repair, DNA methylation, and transcription-related complexes (**Fig. S20**).

We applied our more stringent filter excluding genes with significant residualized expression for female schizophrenic individuals (Mann-Whitney U, p-value < 0.05) and subsequently found 1858, 122, and 104 male-specific schizophrenia DEGs for the caudate nucleus, DLPFC, and hippocampus, respectively. We found a small number of shared genes across the three brain regions, including *RP11-489O18.1* and *CARD11* (**Table S5**). Using gene term enrichment analysis, we observed wide-spread enrichment of multiple pathways across the brain, including neuron and dendrite development in the caudate nucleus; leukocyte differentiation and activation in the DLPFC; and immune response in the hippocampus (**Fig. S21** and **Data S8**).

We hypothesized that the smaller sample size for female individuals might explain why we identified few if any female-specific schizophrenia DEGs within the BrainSeq Consortium dataset. To test this hypothesis, we performed 1000 random samplings of the male individuals at female sample sizes (n = 121, n = 114, and n = 121, for the caudate nucleus, DLPFC, and hippocampus, respectively) and calculated DEGs for each brain region and permutation. On average, we identified a drastically smaller number of schizophrenia DEGs (median male-only schizophrenia DEGs of 200.5, 1, and 0 for the caudate nucleus, DLPFC, and hippocampus, respectively; **Fig. S22**) in our subsampled male samples that showed no significant difference (permutation p-value = 0.44, 0.64, and 0.70 for the caudate nucleus, DLPFC, and hippocampus, respectively) between the number of schizophrenia DEGs identified from the female-only analysis. Altogether, the smaller female sample size, at least partially, explains the lack of identification of female-specific schizophrenia DEGs within the BrainSeq Consortium datasets.

We next compared our results with the recent meta-analysis for sex-specific schizophrenia DEGs in the prefrontal cortex^6^. Of the 46 male-specific DEGs identified by Qin et al., we found a total of three overlapping genes: two overlapping genes (*USE1* and *BBX*) with the caudate nucleus stringent male-specific DEGs and one gene overlapping (*USE1*) with hippocampus stringent male-specific DEGs, which also shared direction of effect. When we compared the full set of male schizophrenia DEGs across brain regions, we found an additional three overlapping genes (*ABCG2, GABARAPL1,* and *PARD3*) shared with the caudate nucleus and one of those three (*ABCG2*) also shared with the DLPFC and hippocampus as male-specific schizophrenia DEG. Of these three only *GABARAPL1* had a discordant direction of effect.

### Sex-dependent eQTL in the brain

We asked whether genetic regulation of expressed features would manifest differently in females compared to males for the caudate nucleus, DLPFC, and hippocampus. We tested for statistical interaction between genotype and sex in the brain by applying multivariate adaptive shrinkage (mash) modeling in the 504 individuals (age > 13) for the caudate nucleus (n=399), DLPFC (n=377), and hippocampus (n=394). We identified hundreds of sex-interacting variants (si-eQTL) across brain regions for gene, transcript, exon, and junction level analysis (**Table 3** and **Data S9**). For example, we found 950, 867, and 830 gene-level si-eQTL (local false sign rate [lfsr] < 0.05) for the caudate nucleus, DLPFC, and hippocampus, respectively, accounting for 974 unique genes across the three brain regions with si-eQTL (eGenes) (**Table 3** and **Data S9**).

**Table 3.** Summary of sex-interacting eQTL (lfsr < 0.05) across brain regions for genes, transcripts, exons, and exon-exon junctions associated with all si-eQTL. eQTL: number of variant-feature pairs for each feature type: genes, transcripts, exons, and junctions. eFeature: number of unique features that have eQTL associations. eGene: number of eQTL associations with unique genes. lfsr: local false sign rate^72^.

| Brain Regions |  | Caudate Nucleus | DLPFC | Hippocampus |
| --- | --- | --- | --- | --- |
| Gene | eQTL | 5785 | 5071 | 4821 |
|  | eFeature | 950 | 867 | 830 |
|  | eGenes | 950 | 867 | 830 |
| Transcript | eQTL | 8525 | 5276 | 5290 |
|  | eFeature | 1381 | 982 | 985 |
|  | eGenes | 1313 | 940 | 943 |
| Exon | eQTL | 11364 | 11331 | 9867 |
|  | eFeature | 2098 | 2085 | 1847 |
|  | eGenes | 1154 | 1137 | 1031 |
| Junction | eQTL | 2479 | 2437 | 2336 |
|  | eFeature | 502 | 489 | 458 |
|  | eGenes | 358 | 342 | 326 |

With only 30 (3.1%) of these eGenes (si-eQTL associated with unique genes) located on the X chromosome, the majority of eGenes appeared to be on autosomes, similar to sex-specific expression analysis.

To understand the regional specificity of these si-eQTL, we examined the proportion of si-eQTL detected across brain regions. Here, we found the majority (814 [84%]) of eGenes were shared across brain regions (**Fig. 4A**), which was also on the transcript, exon, and junction level (**Fig. S23**). Remarkably, all of the shared si-eQTL showed concordant directionality with the DLPFC and hippocampus showing nearly identical si-eQTL effect sizes (**Fig. 4B**). The few brain-region specific si-eQTL showed small but significant sexual dimorphic genetic regulation of expression (**Fig. S24**). Unsurprisingly, the exon and junction level sharing showed a smaller proportion of shared si-eQTL for the DLPFC and hippocampus at an effect size within a factor of 0.99 (**Fig. S25**), suggesting alternative isoform usage drives differences in si-eQTL effect size between the two brain regions.

**Fig. 4.**
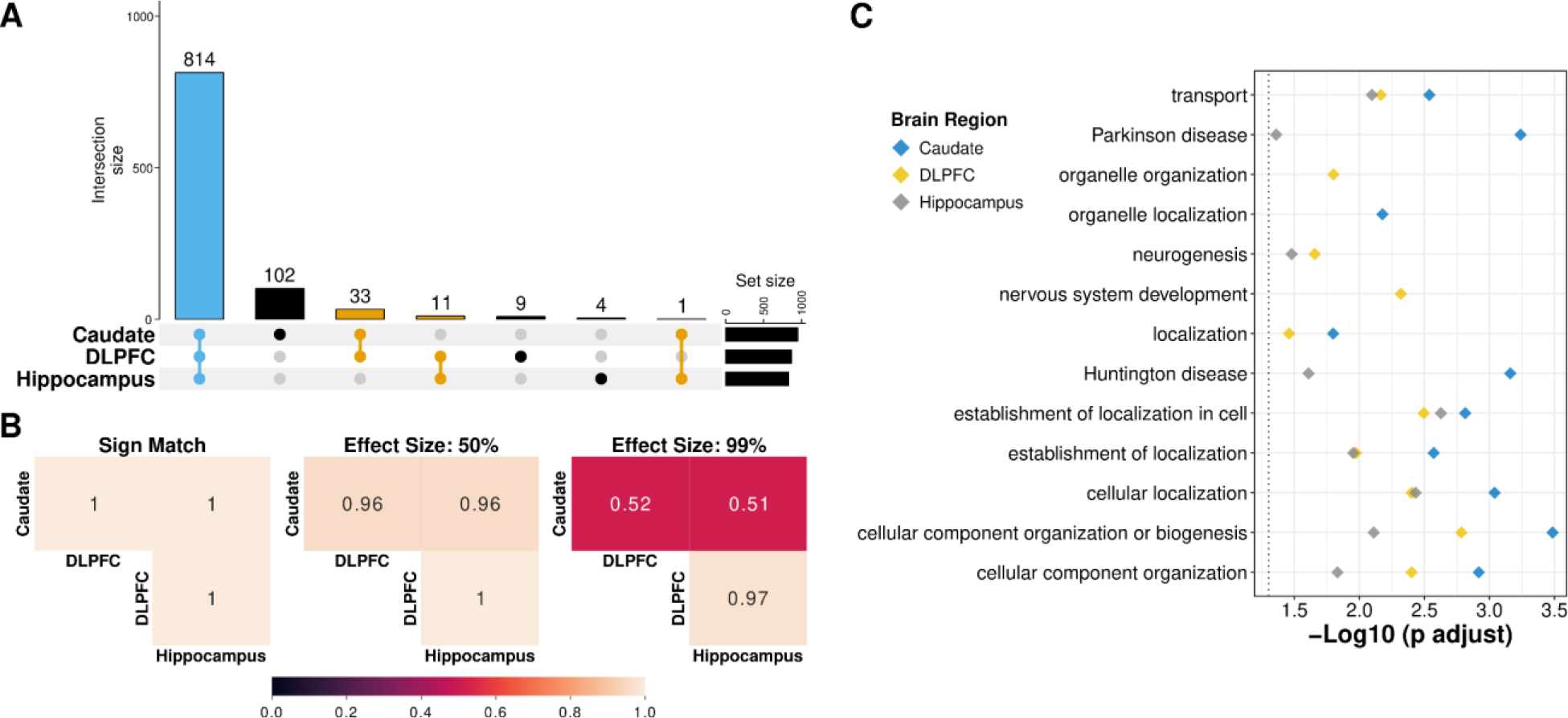
Sex-interacting eQTL are shared across brain regions. **A.** UpSet plot displaying overlap across brain regions for si-eQTL (lfsr < 0.05). Blue is shared across caudate nucleus (n=399; 126 female and 273 male), DLPFC (n=377; 121 female and 256 male), and hippocampus (n=394; 126 female and 268 male); orange, shared between two brain regions; and black, unique to a specific brain region. **B.** Heatmap of the proportion of gene level si-eQTL sharing with sign match (left), within a factor of 0.5 effect size (middle), and within a factor 0.99 effect size (right). **C.** Functional enrichment plot of the ten most significant KEGG and GO Biological Processes of eGenes for the caudate nucleus (blue), DLPFC (yellow), and hippocampus (gray). Female (red); male (blue). Schizophrenia risk increases from left (0) to right (2). lfsr: local false sign rate^72^.

To evaluate the functional relevance underlying si-eQTL in the caudate nucleus, DLPFC, and hippocampus, we performed functional gene term enrichment analysis on the eGenes for each brain region. We observed significant enrichment (hypergeometric, adj. p-value < 0.05) across brain regions (**Data S10**), including enrichment for neurogenesis as well as Parkinson and Huntington diseases (**Fig. 4C**). Notably, we found that these enriched GO terms showed high semantic similarity^43^ (Best Match Average >87%) across brain regions (**Fig. S26**). Additionally, we found these si-eQTL associated eGenes were enriched (Fisher’s exact test, FDR corrected p-value < 0.05) for neuropsychiatric disorders including schizophrenia^12, 13, 15, 70^, autism spectrum disorder^70^, and bipolar disorder^70^ (**Fig. S27**).

When we compared our si-eQTL with previous work in whole blood and in lymphoblastoid cell lines, we found no overlap with 19 si-eQTL identified in whole blood^16, 17^ and one gene (*ATG4C*) of the 21 si-eQTL identified in lymphoblastoid cell lines^18^ also present in the caudate nucleus si-eQTL. We next compared our results with the four si-eQTL (q-value < 0.25) identified in GTEx brain regions (amygdala and nucleus accumbens basal ganglia)^8^ and found no overlaps. When we expanded to the 369 si-eQTL (q-value < 0.25) from all 43 GTEx tissues^8^, we found 14 overlapping genes (*FAM129B, CALM2, TMEM136, ANKRD65, ATP8A1, UFSP2, COL11A2, VWDE, LITAF, MAG, ENOSF1, ENSG00000267056, ENSG00000268362,* and *ENSG00000272977*) between the caudate nucleus and multiple GTEx tissues (adipose subcutaneous, adipose visceral omentum, artery coronary, breast mammary tissue, muscle skeletal, and spleen; **Data S11**). The relatively low replication rate with GTEx brain regions can, in part, be attributed to low sample sizes in the GTEx dataset^71^.

We next set out to determine if any of these si-eQTL had causal associations with schizophrenia risk (PGC3 GWAS p-value < 5e-8)^49^. To formally identify variants associated with schizophrenia risk, we performed colocalization analysis on fine-mapped gene level si-eQTL across brain regions (**Data S12**). We identified 14 unique genes across the three brain regions with significant colocalization (regional colocalization probability [RCP] > 0.5; **Data S13**): two genes (*RP11-399D6.2* and *ELAC2*; **Fig. S28** and **Data S14**) in the caudate nucleus; 13 genes in the DLPFC (*RP11-430H10.1*, *ZSCAN29*, *ENSG00000249839*, *FURIN*, *ENSG00000278434*, *ELAC2*, *MMD*, *ACE*, *LINC00320*, *FOXN2*, *FTCDNL1*, *TTYH3*, and *IQANK1*; **Fig. S29** and **Data S14**); and two genes in the hippocampus (*FURIN* and *MMD*). Interestingly, the highest signal si-eQTL associated with schizophrenia risk was associated with the DLPFC, and three of the four genes identified in the caudate nucleus (*ELAC2*) and hippocampus (*FURIN* and *MMD*; **Fig. S30** and **Data S14**) also shared with the DLPFC.

## Discussion

Sex has been associated with differential gene expression in the brain and sex-specific effects in neuropsychiatric disorders like schizophrenia. Here, we aimed to take a holistic exploratory analysis approach to sex differences for schizophrenia in the caudate nucleus, DLPFC, and hippocampus, and identified numerous genetic features (genes, transcripts, exons, and exon-exon junctions) that 1) are associated with sex, 2) demonstrate sex-specific expression in schizophrenia, and 3) whose expression interacts with genotypes and sex as si-eQTL. Furthermore, we identified 14 genes showing sex variable association with schizophrenia risk^49^ (PGC3 GWAS p-value < 5e-8) with colocalization. Additionally, we demonstrate slight differences between X-chromosome dosage in the hippocampus of schizophrenic individuals as well as variation of dosage compensation in the brain. To the best of our knowledge, this is the largest multi-brain region analysis for sex differences in schizophrenia.

In this study, we found 878 unique genes with sex-associated differential expression in the caudate nucleus, DLFPC, and hippocampus. Unsurprisingly, allosomes, while making up a small portion, heavily influenced sex differences. Our results support previous findings of sex differences, including differences that: are brain-region specific^59^; are primarily located on autosomes^7, 57, 58^; and have exhibited shared direction of effect driven by allosomal DEGs^59^. Larger number of samples allowed us to identify many more genes than previous analyses conducted across multiple regions, contributing to the up-to-date knowledge on sex-specific genomic features and sex differences in the brain.

Of particular interest to us was the identification of a sex dependent variation in expression of *ANK3,* a marker of early life stress associated with schizophrenia risk. This gene showed male-bias sex differences in the caudate nucleus on gene, exon, and junction levels. Moreover, a formal interaction model also found sex and brain region differences for *ANK3* with male-bias expression present in the caudate nucleus. Previous work for sex differences in mice found no observable differences using RT-PCR^67^; however, this study was considered only the prefrontal cortex while we found *ANK3* expression to be higher in the caudate nucleus compared to the DLPFC or hippocampus.

To determine if sex differences observed across brain regions were related to X-chromosome inactivation, we further examined DEGs located on the X chromosome. Here, our analysis aligned with previous work showing an enrichment of genes known to escape XCI^45, 68^. Additionally, we found that across the brain (i.e., GTEx and BrainSeq Consortium) X-chromosome dosage showed brain-region specific dosage compensation levels.

The second aim of the study was to identify any sex-specific schizophrenia genomic features. While our interaction model found only 15 novel junctions across brain regions, comparing female to male schizophrenia expression allowing us to identify magnitudes more sex-specific schizophrenia genomic features than previous studies – even after applying an additional, more stringent filter to our sex-specific schizophrenia DEG results. As the first study to compare multiple brain regions, we found that sex-specific schizophrenia differentially expressed features were highly brain-region specific. This was not unexpected as DEGs for schizophrenia, irrespective of sex, is also highly brain-region specific^11, 12, 15^. However, the smaller number of identified DE features for transcripts, exons, and junctions was surprising. This might be due to our study being underpowered, as we found twice as many schizophrenia differentially expressed features in male individuals as compared to females. Altogether, these findings suggest that an increase of samples from female individuals would further advance our understanding of potential sex differences in multiple brain regions.

In addition to our expression analysis, we also annotated hundreds of sex-interacting eQTL associated with 974 unique genes (eGenes) for the caudate nucleus, DLPFC, and hippocampus using mash modeling^72^ that increases the power of detection while also improving effect size estimates. With this study, we provided the first annotation of si-eQTL in the DLPFC and hippocampus. In addition, we found that these sex-interacting eGenes were enriched for neurological disorders, such as Parkinson’s and Huntington’s diseases, which show significant enrichment for the caudate nucleus and hippocampus. Given the association of cognitive dysfunction with the caudate nucleus^73^, as well as previous reports showing sex differences in Parkinson’s and Huntington’s diseases for cognitive dysfunction^74–76^, this is not unexpected. Even so, our results demonstrate the power of tissue-specific sex-interacting eQTL and its potential for identifying genes with sexually dimorphic expression for neurological disorders as novel therapeutic targets.

In addition to annotating hundreds of si-eQTL, we also provide the first annotation of sex-interacting genes with causal variants associated with schizophrenia risk. This highlights the importance of examining genes associated with schizophrenia risk for potential differences in expression for population covariates (e.g., sex, age, and ancestry).

In summary, we provided a comprehensive genetic and transcriptional analysis of sex differences in schizophrenia. We increased the number of annotated features exhibiting sex bias in the brain adding to our current understanding of sex differences in the brain, identified sex-specific schizophrenia genes with indications for novel therapeutic targets, and provided the first annotation of sex-interacting eQTL for the DLPFC and hippocampus. These results have the potential to direct novel therapeutics and new strategies that can address sex-biased responses in the treatment of schizophrenia.

## Data Availability

Publicly available BrainSeq Consortium total RNA DLPFC and hippocampus RangedSummarizedExperiment R Objects with processed counts are available at http://eqtl.brainseq.org/phase2/. Publicly available BrainSeq Consortium total RNA caudate nucleus RangedSummarizedExperiment R Objects with processed counts are available at http://erwinpaquolalab.libd.org/caudate_eqtl/. Analysis-ready genotype data will be shared with researchers that obtain dbGaP access. FASTQ files are available for total RNA DLPFC and hippocampus via Globus collections jhpce#bsp2-dlpfc and jhpce#bsp2-hippo. For the caudate nucleus, FASTQ files are available on NCBI SRA under project ID PRJNA874683. PGC3 GWAS summary statistics are available at https://figshare.com/articles/dataset/scz2022/19426775.

https://doi.org/10.5281/zenodo.7125280

## Acknowledgements

The authors would like to extend their appreciation to the Offices of the Chief Medical Examiner of Washington DC, Northern Virginia, and Maryland for the provision of brain tissue used in this work. The authors also extend their appreciation to Dr. Llewellyn B. Bigelow and members of the LIBD Neuropathology Section for their work in assembling and curating the clinical and demographic information and organizing the Human Brain Tissue Repository of the Lieber Institute. Finally, the authors gratefully acknowledge the families that have donated this tissue to advance our understanding of psychiatric disorders. This work is supported by the Lieber Institute for Brain Development, the National Institutes of Health (NIH) T32 fellowship (T32MH015330) and K99 award (K99MD016964) to KJMB, NIH R01 (R01MH123183) to LC-T, and a NARSAD Young Investigator Grant from the Brain & Behavior Research Foundation to JAE. RA would like to thank the Hopkins Office for Undergraduate Research (HOUR), Johns Hopkins University for their support through the Summer PURA program as well as the Albstein Research Scholarship for their support.

## Author Contributions

Conceptualization, KJMB, RA, and JAE; Methodology, KJMB, RA, JMS, LD, ACMP, and JAE; Software, KJMB, RA, and ACMP; Formal Analysis, KJMB and RA; Investigation, JHS and TMH; Data Curation, KJMB, RA, and JEK; Writing – Original Draft, KJMB, RA, and JAE; Writing – Review & Editing, KJMB, RA, LC-T, ACMP, DRW, and JAE; Visualization, KJMB, RA, and LD; Supervision, ACMP and JAE; Project Administration, JAE; Funding Acquisition, KJMB, RA, LC-T, DRW, and JAE.

## Data availability

Publicly available BrainSeq Consortium total RNA DLPFC and hippocampus RangedSummarizedExperiment R Objects with processed counts are available at http://eqtl.brainseq.org/phase2/. Publicly available BrainSeq Consortium total RNA caudate nucleus RangedSummarizedExperiment R Objects with processed counts are available at http://erwinpaquolalab.libd.org/caudate_eqtl/. Analysis-ready genotype data will be shared with researchers that obtain dbGaP access. FASTQ files are available for total RNA DLPFC and hippocampus via <u>Globus</u> collections <u>jhpce#bsp2-dlpfc</u> and <u>jhpce#bsp2-hippo</u>. For the caudate nucleus, FASTQ files are available on NCBI SRA under project ID PRJNA874683. PGC3 GWAS summary statistics are available at https://figshare.com/articles/dataset/scz2022/19426775.

## Supplementary information

### Figures

**Fig. S1.**
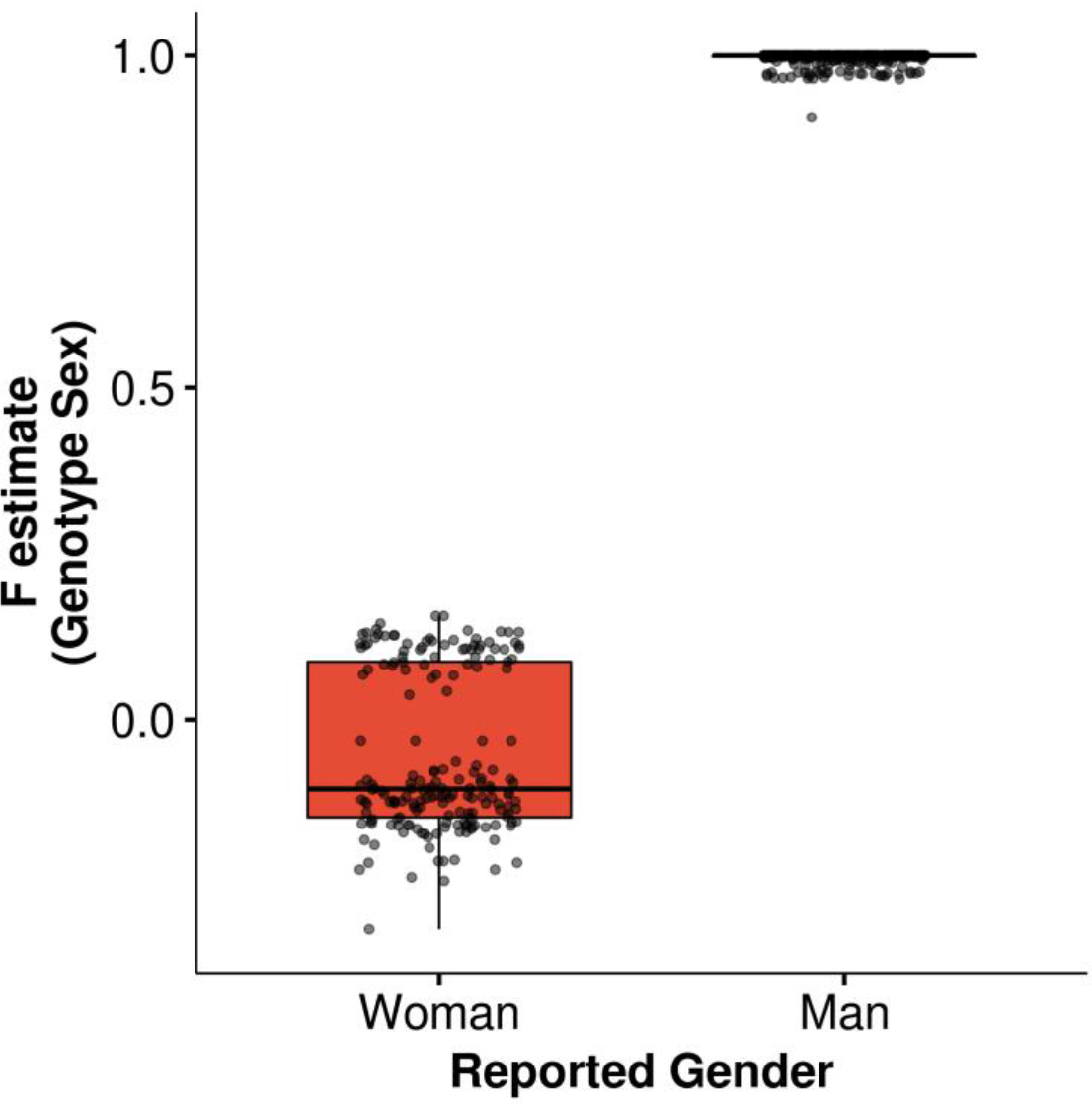
Completed overlap of reported gender with sex genotype. Boxplot showing F estimates of genotype sex for female and male individuals correlate with reported gender (i.e., woman or man).

**Fig. S2.**
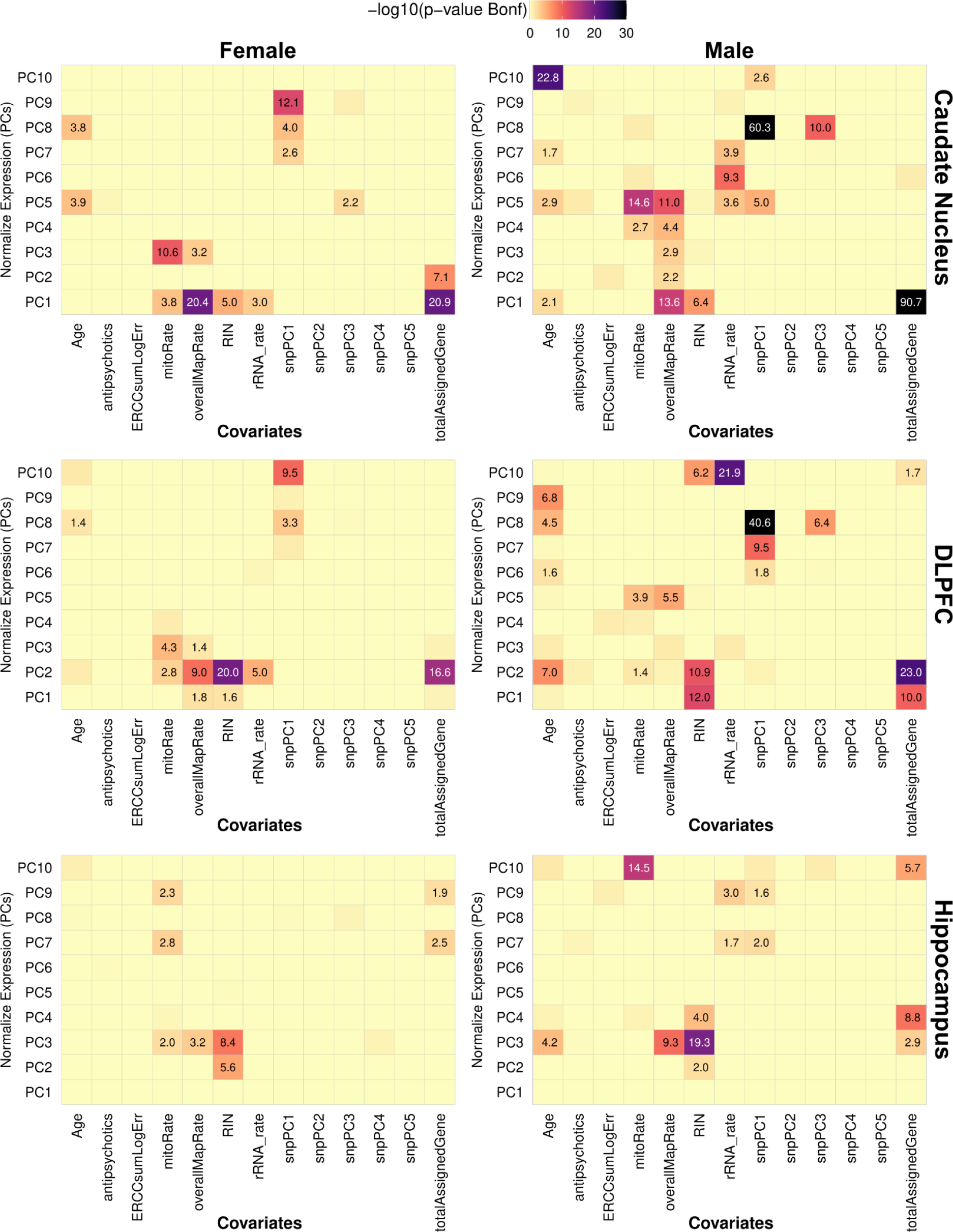
Correlation of covariates with gene expression by sex. Heatmaps showing correlation between PCA of normalized gene expression and covariates separated by sex. A value of 1.3 or greater is significant and equivalent to Bonferroni corrected p-value < 0.05. Significant correlations are denoted within each tile.

**Fig. S3.**
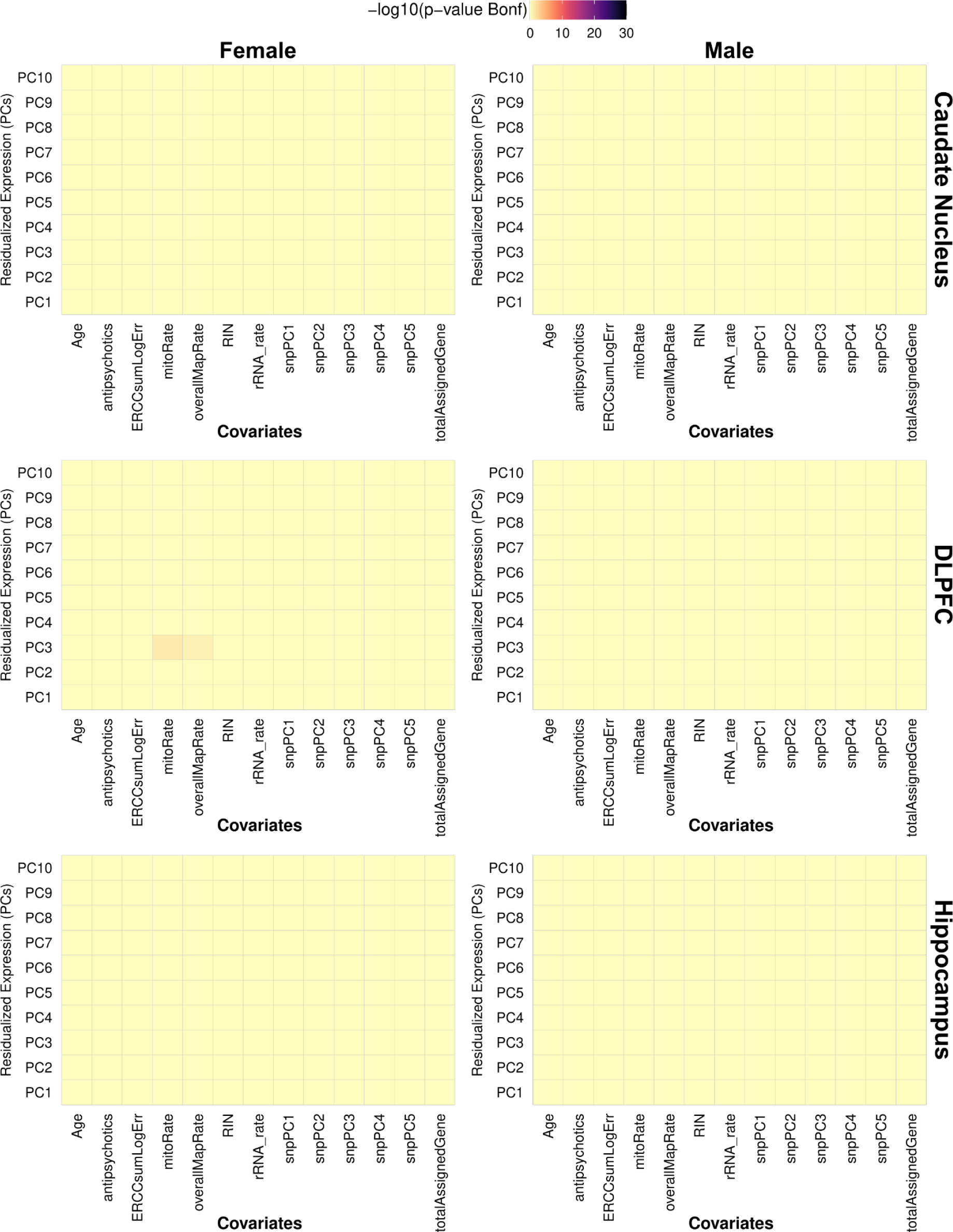
Correction for confounders eliminates significant correlation with gene expression. Heatmaps showing no correlation between PCA of normalized gene expression and covariates separated by sex. A value of 1.3 or greater is significant and equivalent to Bonferroni corrected p-value < 0.05. Significant correlations are denoted within each tile.

**Fig. S4.**
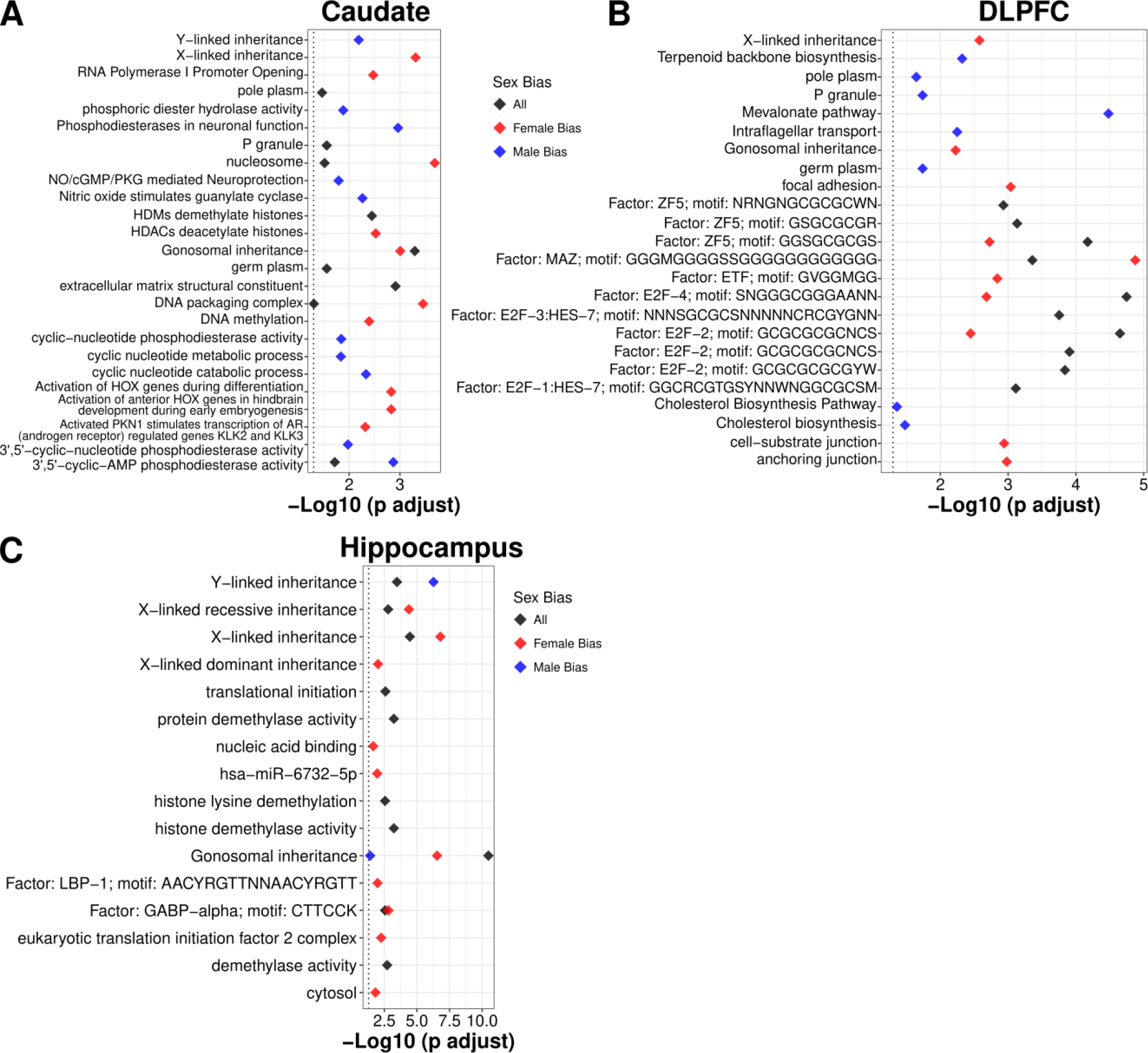
Gene term enrichment for gene expression analysis of sex in the **A.** caudate nucleus, **B.** DLPFC, and **C.** hippocampus. Red denotes female bias; blue denotes male bias.

**Fig. S5.**
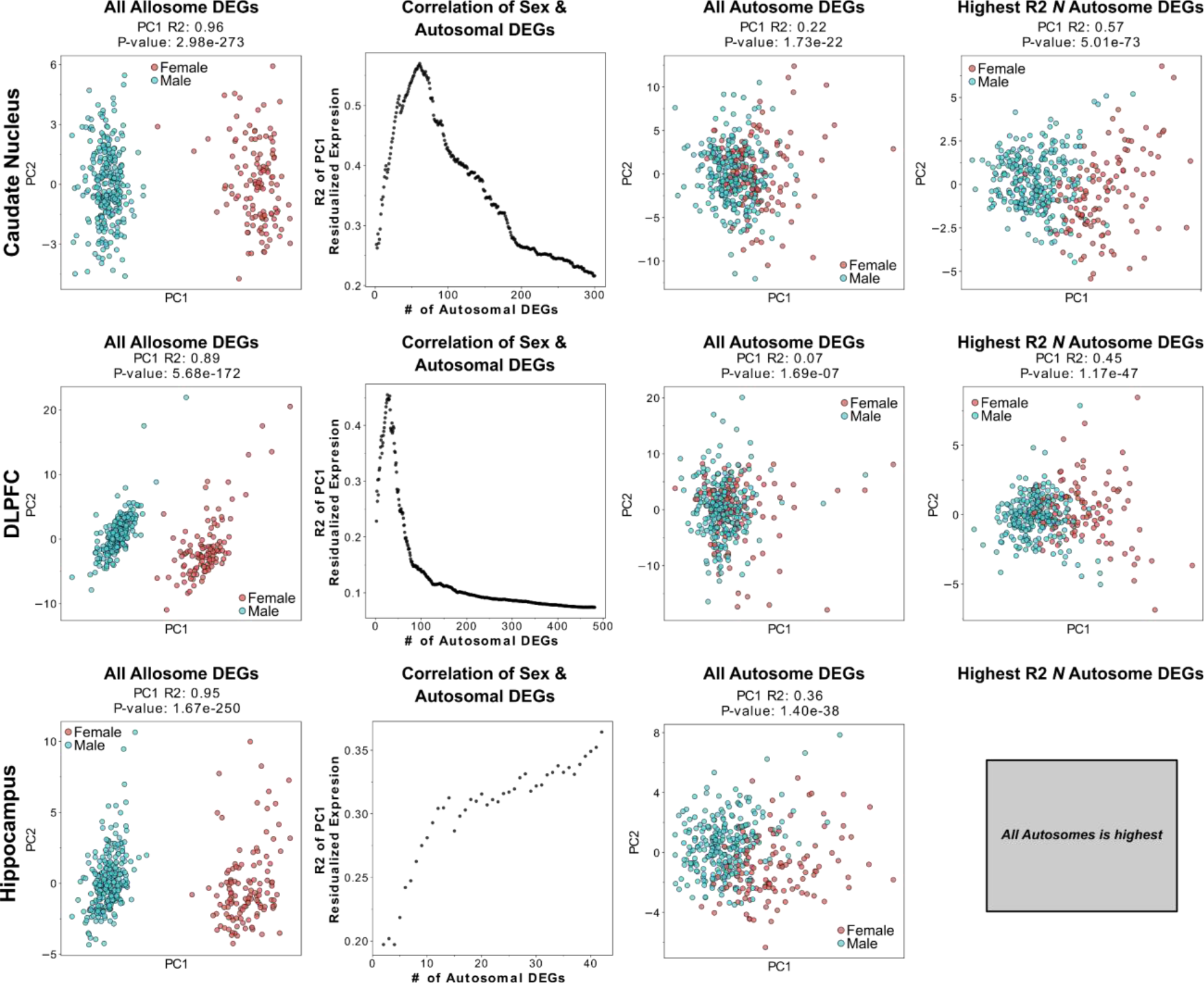
Autosomal DEGs show significant correlation with sex as well as show predictive power for sex in the brain. Scatter plots of principal components (PC) 1 and 2 from dimensionally reduced expression of all allosomal and autosomal DEGs for the caudate nucleus, DLPFC, and hippocampus.

**Fig. S6.**
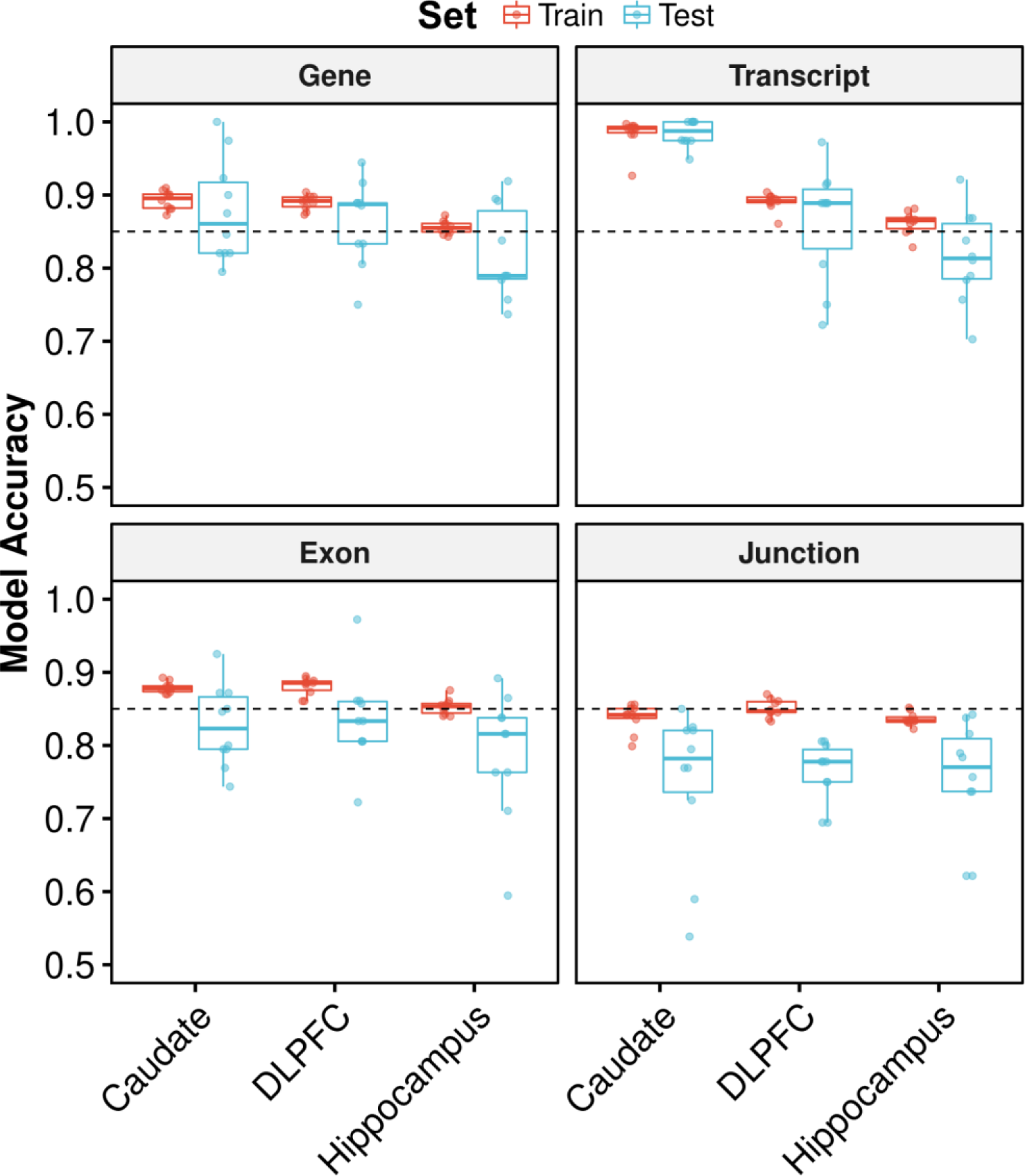
Metric summary across brain region and features show high classification accuracy for sex using autosomes. Boxplot of train (red) and test (blue) accuracy for sex classification in the caudate nucleus, DLPFC, and hippocampus for genes, transcripts, exons, and junctions. Each point represents results from one fold within the 10-fold cross-validation. Dashed line denotes 85% accuracy.

**Fig. S7.**
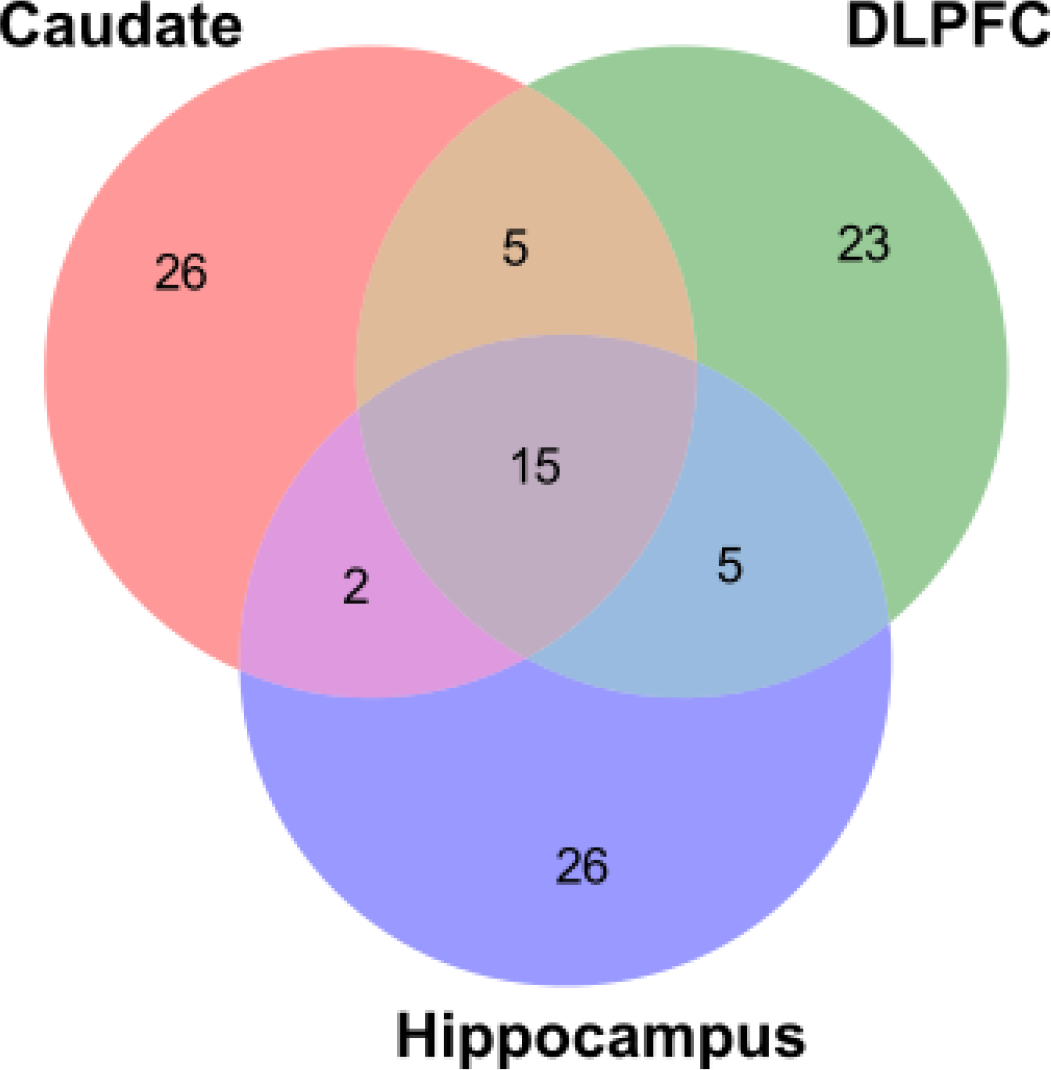
Venn diagram showing the smallest subset of predictive autosomal genes are brain region specific.

**Fig. S8.**
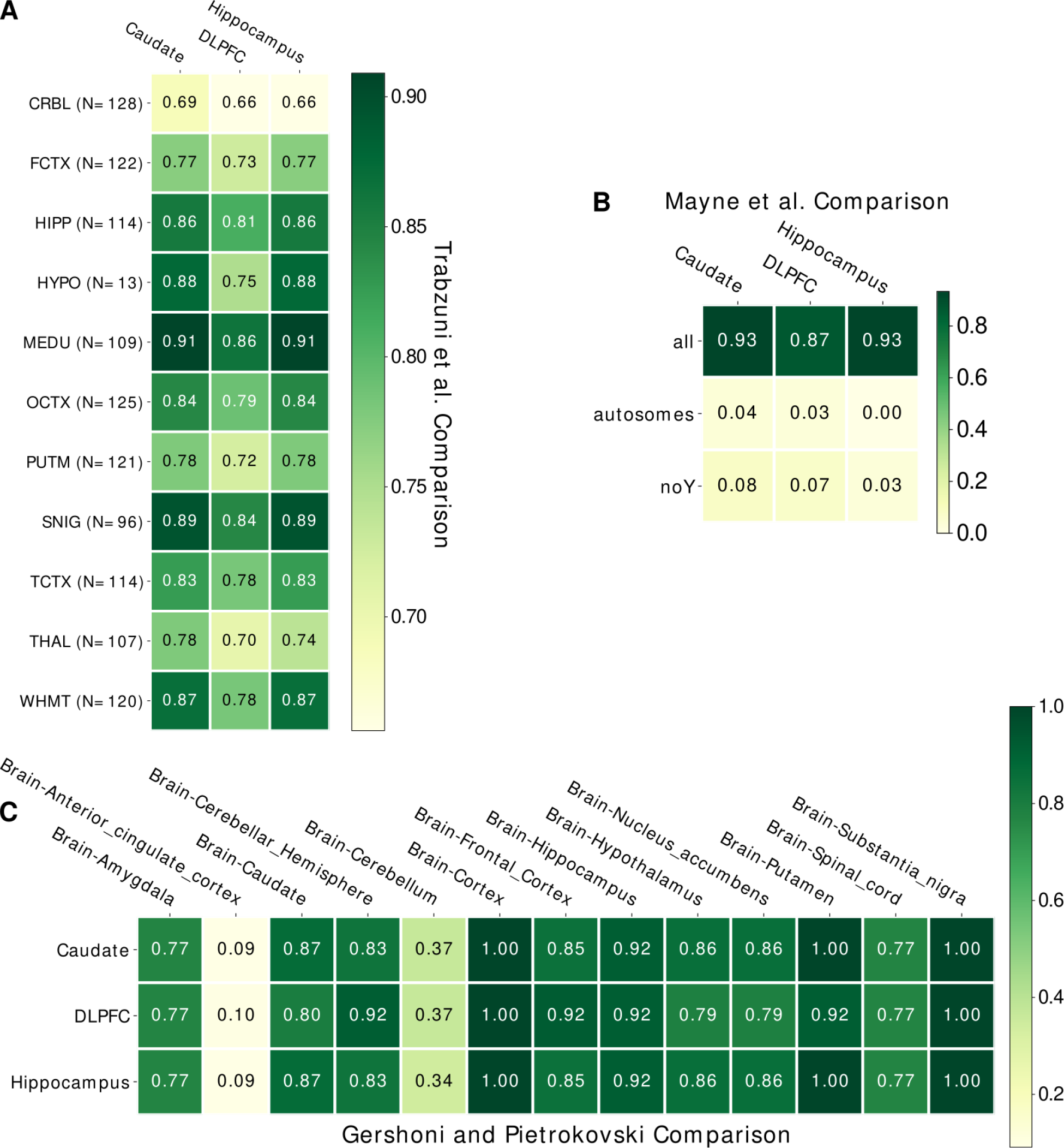
High overlap of differentially expressed genes across multiple datasets and brain regions. Heatmaps comparing BrainSeq Consortium brain regions that show the ratio of differentially expressed genes overlap with **A.** Trabzuni et al. brain regions, **B.** Mayne et al. meta analysis with significant expression in at least one brain region using all chromosomes, only autosomes, or no Y chromosomes, and **C.** Gershoni and Pietrokovski brain regions. Abbreviations: CRBL: cerebellum, FCTX: frontal cortex, HIPP: hippocampus, HYPO: hypothalamus, MEDU: medulla, OCTX: occipital cortex, PUTM: putamen, SNIG: substantia nigra, TCTX: temporal cortex, THAL: thalamus, and WHMT: white matter.

**Fig. S9.**
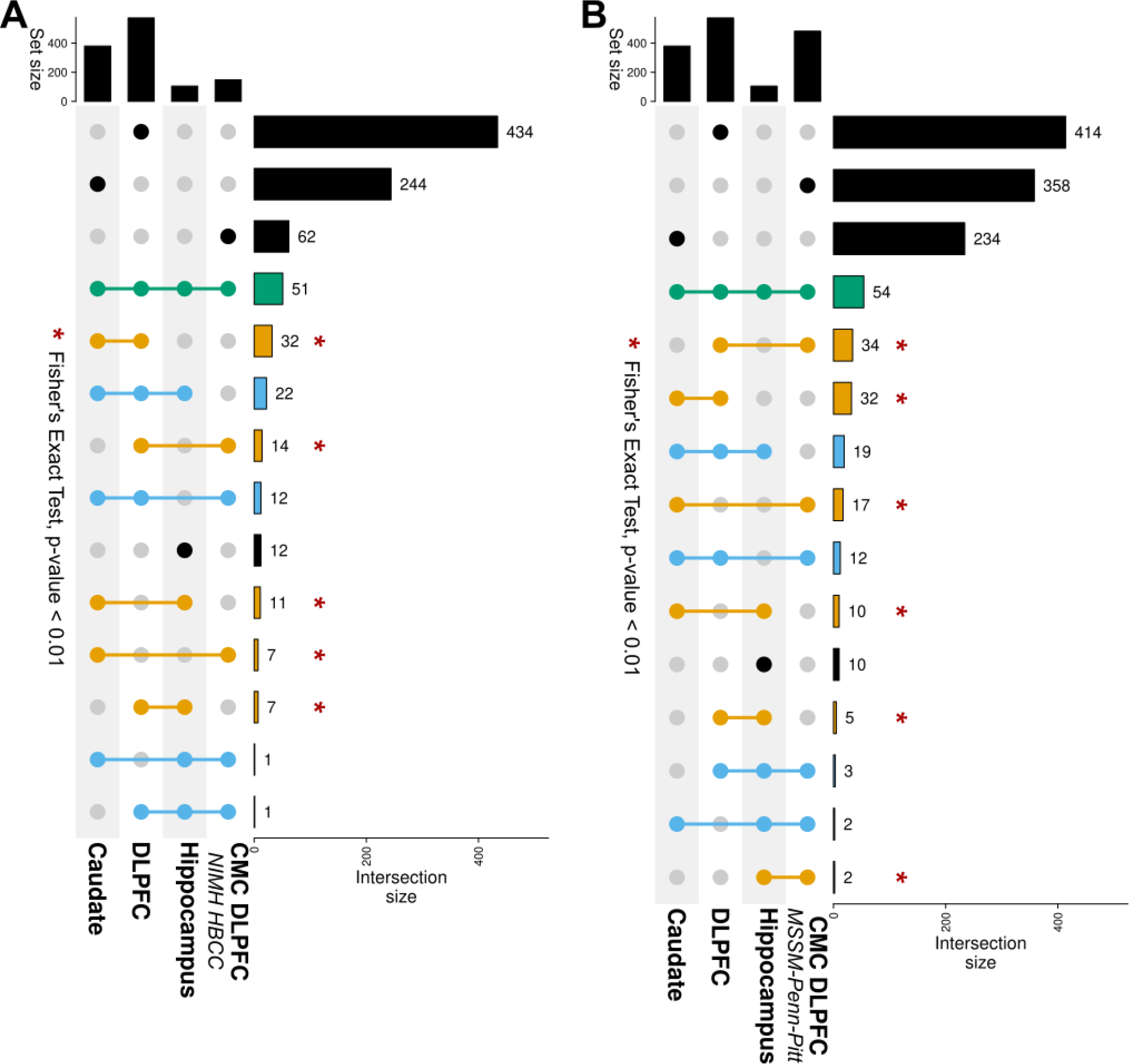
Significant sharing of sex-biased DEGs across brain regions replicate in the CommonMind (CMC) DLPFC. UpSet plot showing number of differentially expressed genes shared across brain regions with the CMC DLPFC cohort **A.** NIMH HBCC and **B.** MSSM-Penn-Pitt. **\*** Indicating p-value < 0.01 for two-tailed, Fisher’s exact test.

**Fig. S10.**
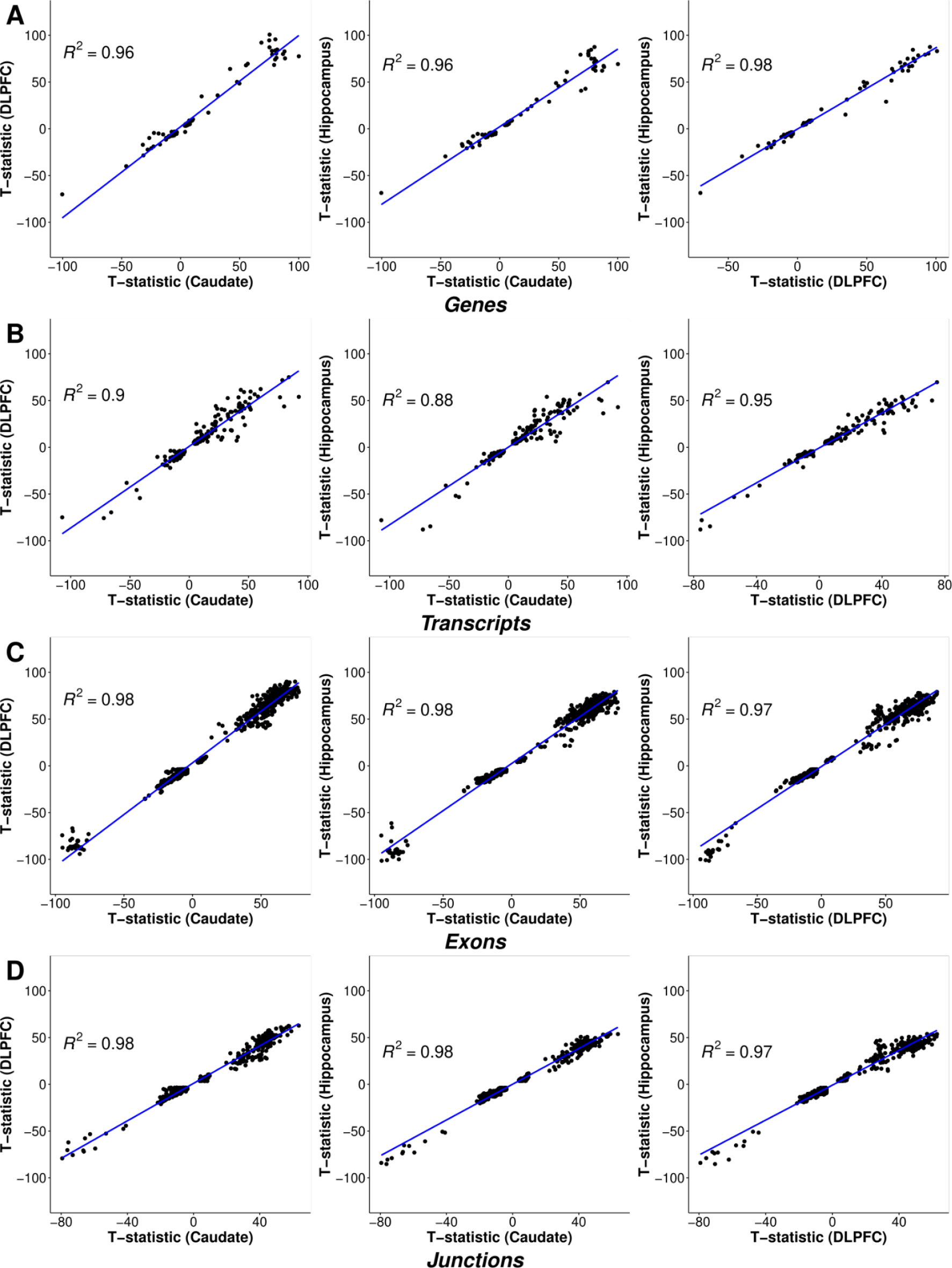
All significantly differentially expressed features (FDR < 0.05) have concordant directionality for sex differences across the three brain regions. Scatter plot of Pearson correlation comparing t-statistic from sex differentially expressed features (FDR < 0.05) between brain region pairs for **A**. genes, **B**. transcripts, **C.** exons, and **D**. junctions.

**Fig. S11.**
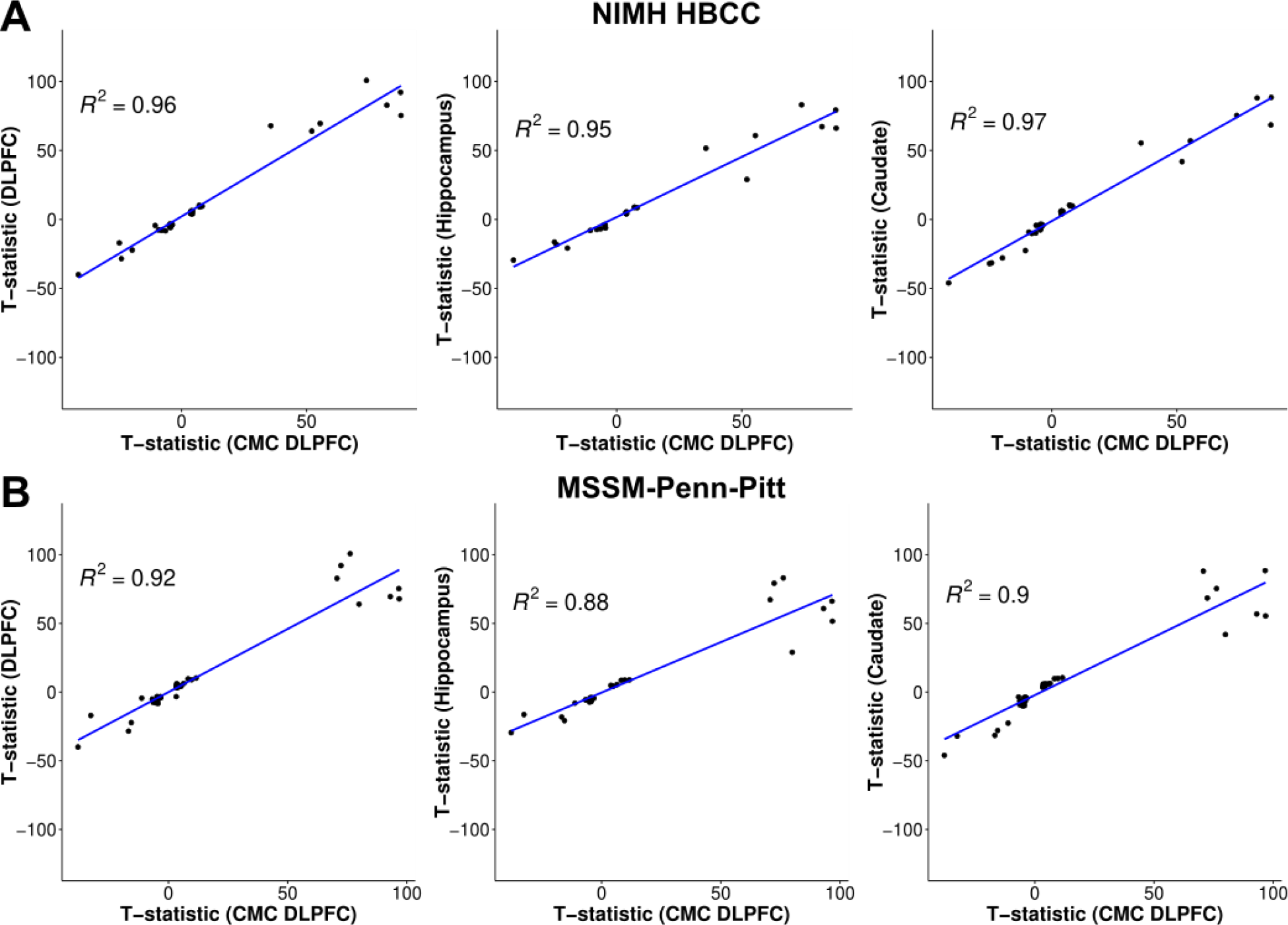
Replication of concordant directionality of sex DEGs with CommonMind (CMC) DLPFC. Scatter plot comparing t-statistics between BrainSeq Consortium brain regions (caudate nucleus, DLPFC, and hippocampus) and CMC DLPFC cohort **A.** NIMH HBCC and **B.** MSSM-Penn-Pitt. Pearson correlation (R^2^) in blue.

**Fig. S12.**
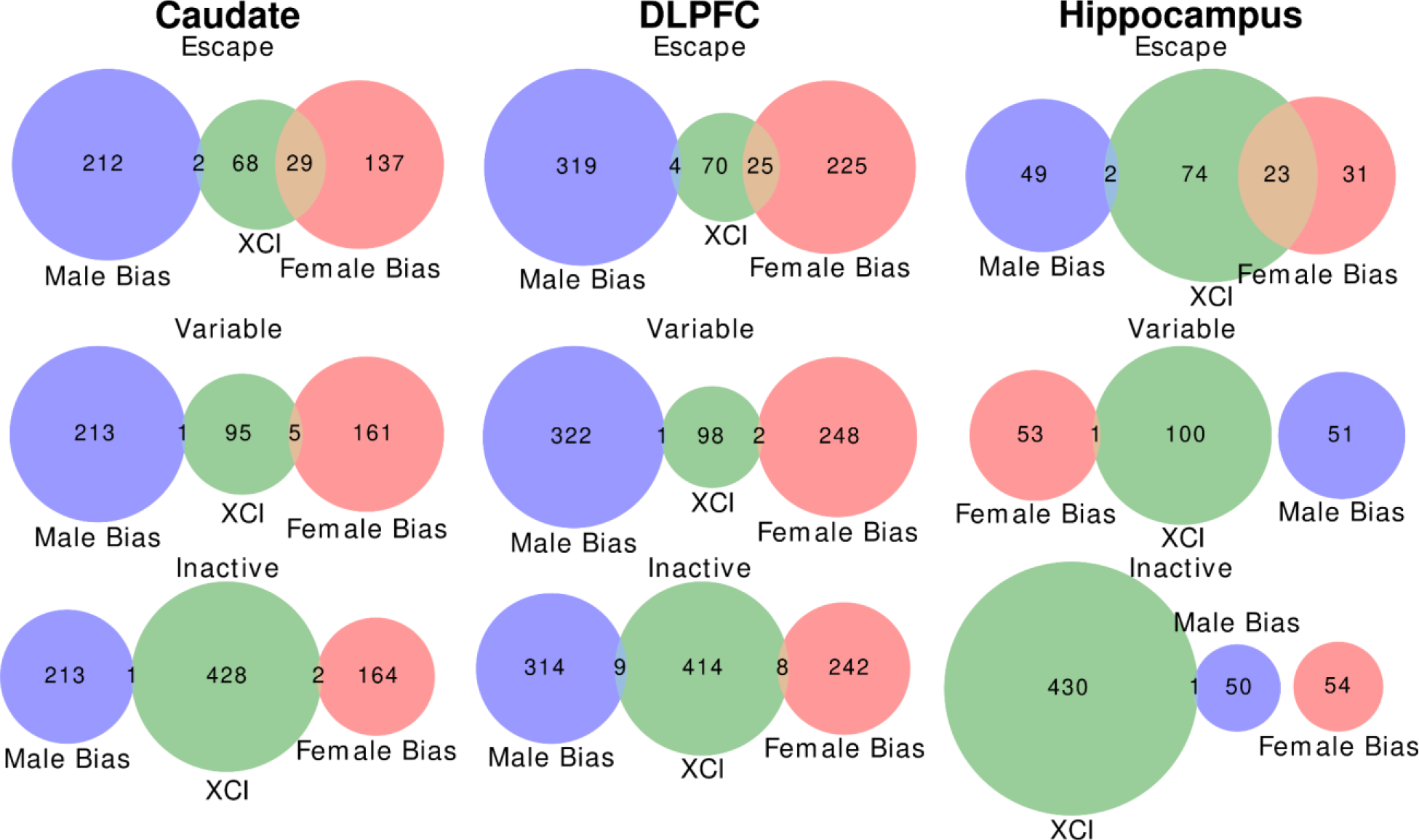
Venn diagram demonstrating large overlap of escaping X-chromosome inactivation (XCI) genes with DEGs up-regulated in female individuals (female bias; red) compared with variable and inactive XCI genes for the caudate nucleus, DLPFC, and hippocampus. DEGs up-regulated in male individuals (male bias; blue). XCI genes in green.

**Fig. S13.**
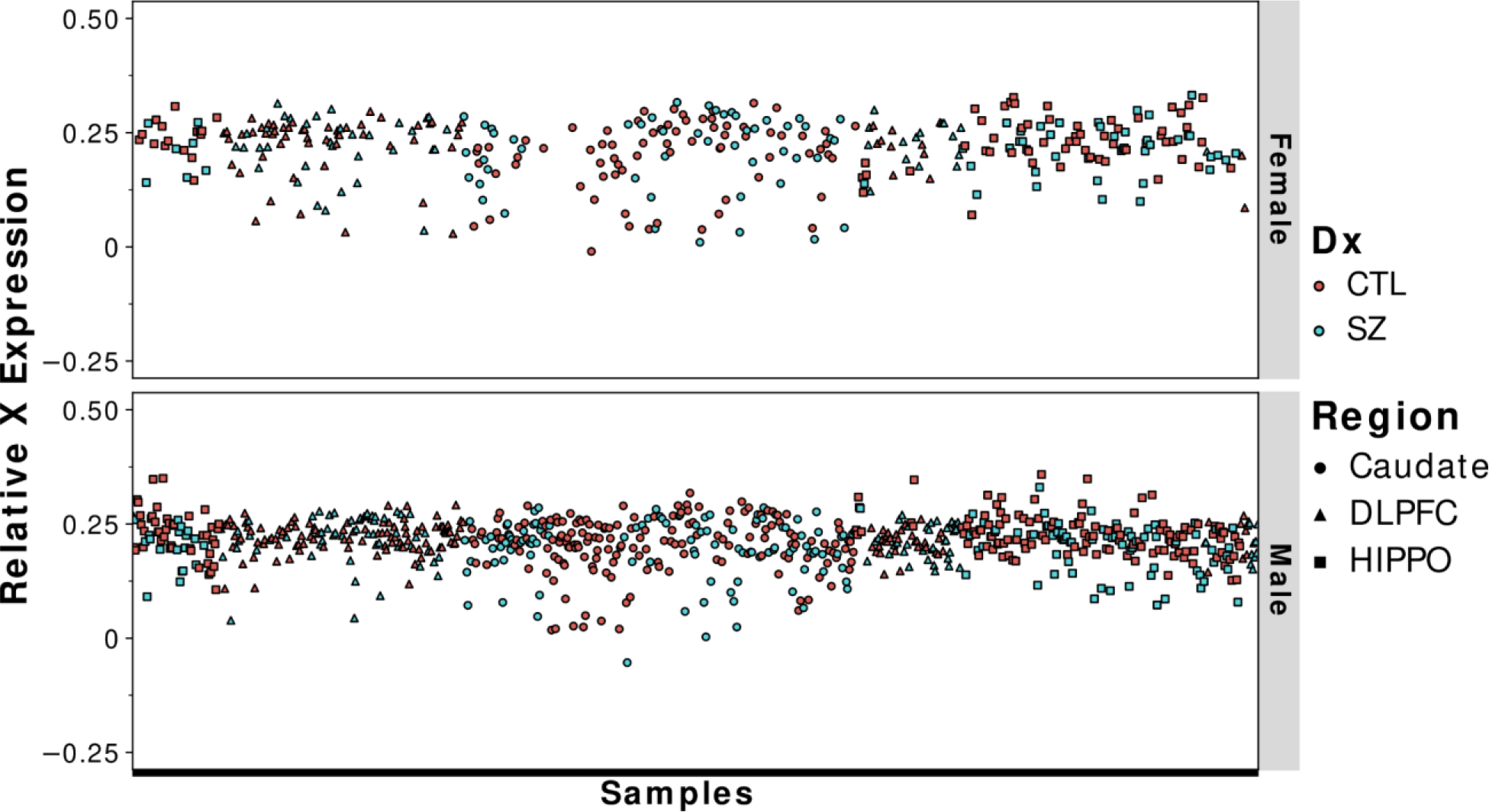
Dosage is properly compensated in the caudate nucleus, DLPFC, and hippocampus. Scatterplot of relative X expression (RXE) across brain regions separated by sex. Control (CTL; red), Schizophrenia (SZ; blue), caudate nucleus (circle), DLPFC (triangle), and hippocampus (square).

**Fig. S14.**
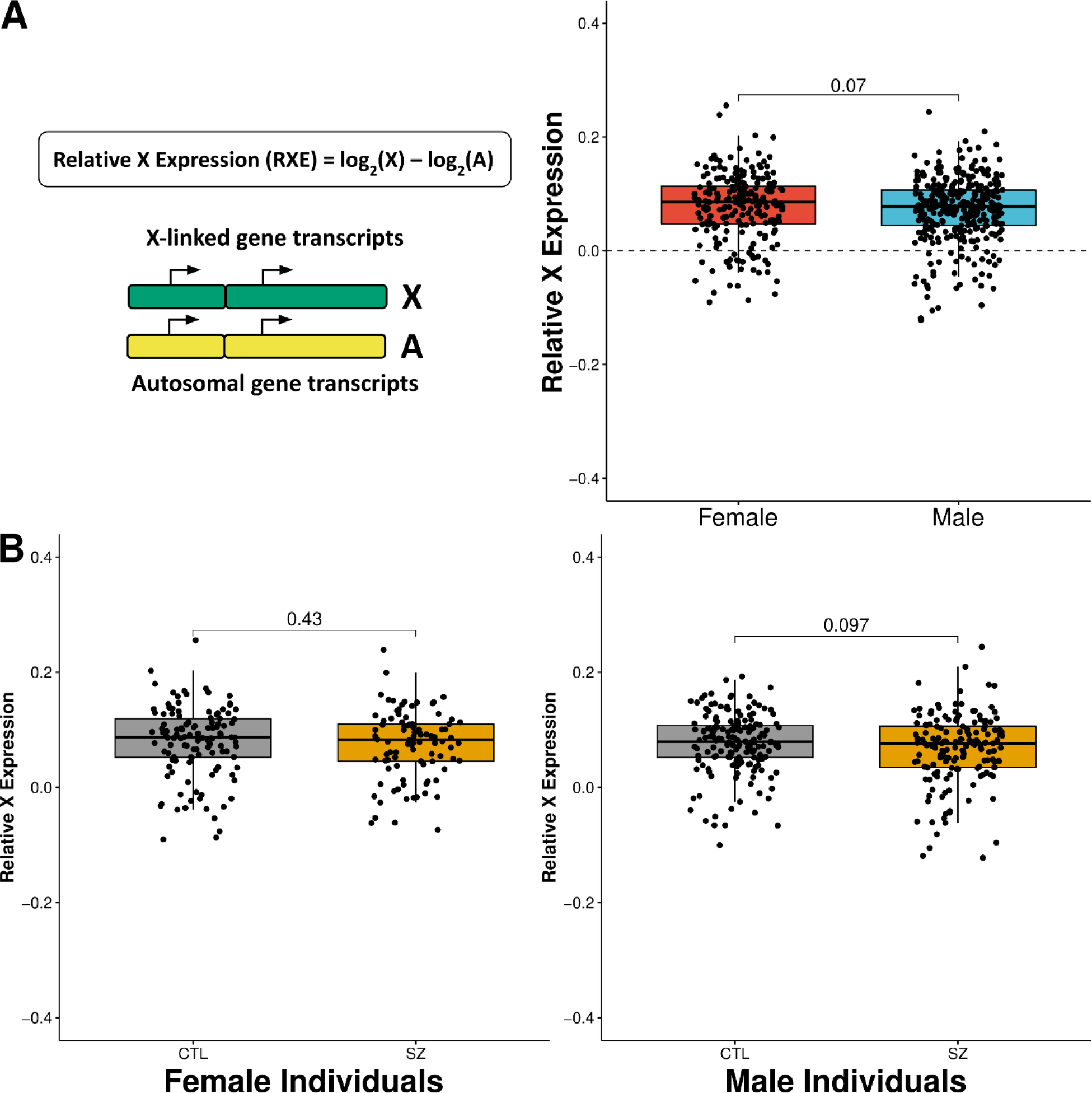
Replication of relative X expression (RXE) sex differences within the DLPFC of CommonMind Consortium (CMC). A. Schematic of relative X expression (left) and boxplots showing RXE comparison between female (red) and male (blue) individuals for the CMC DLPFC (right). **B.** Boxplots showing RXE comparison between neurotypical controls (gray) and schizophrenia (gold) individuals for female (left) and male (right) individuals in the CMC DLPFC. Mann-Whitney U two-tailed test.

**Fig. S15.**
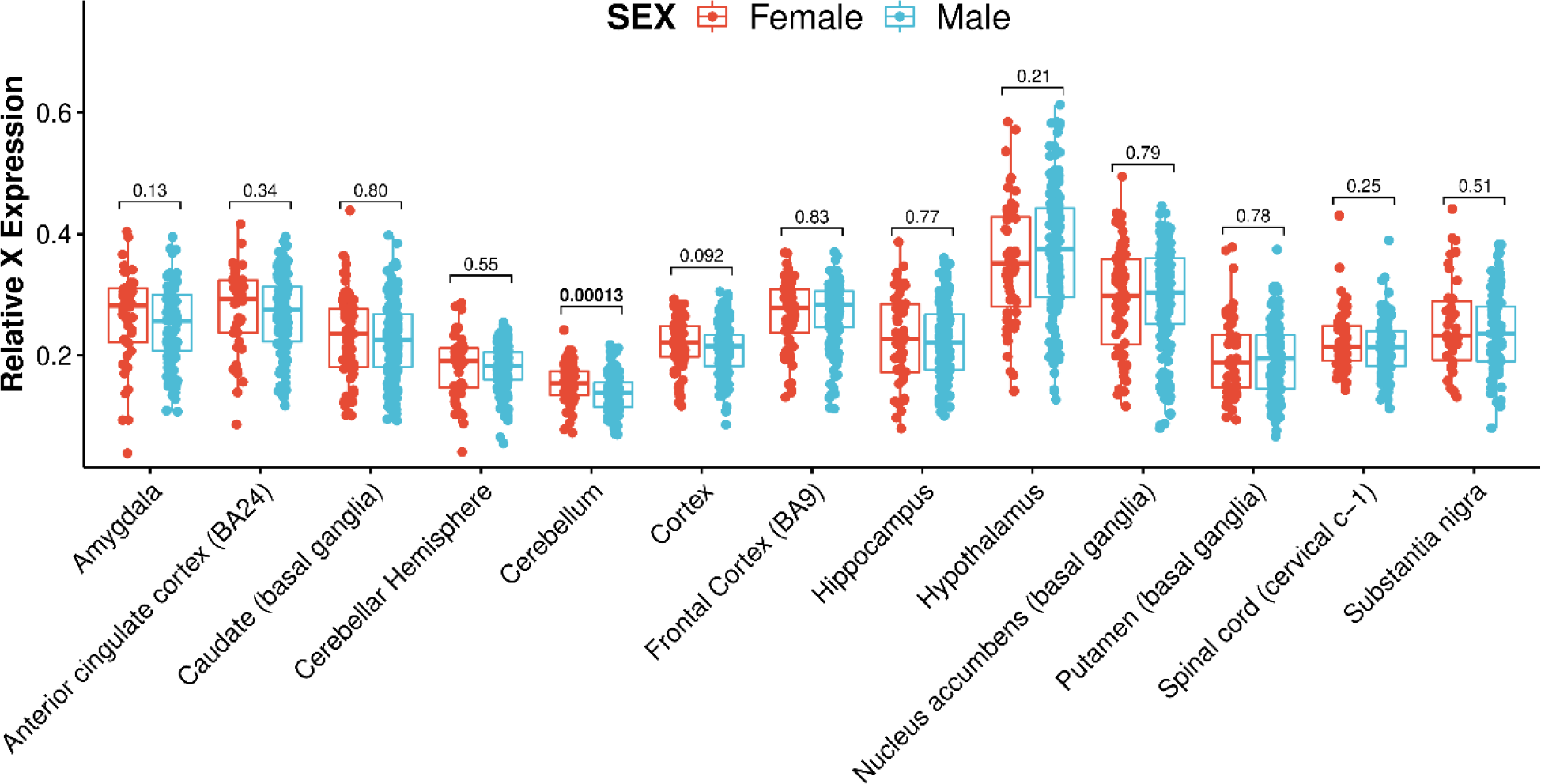
Replication of relative X expression (RXE) sex differences within the 13 brain regions of GTEx shows RXE variation within GTEx brain regions. Boxplots showing RXE comparison between female (red) and male (blue) individuals for the 13 GTEx brain regions. Mann-Whitney U two-tailed test. Significant difference bolded.

**Fig. S16.**
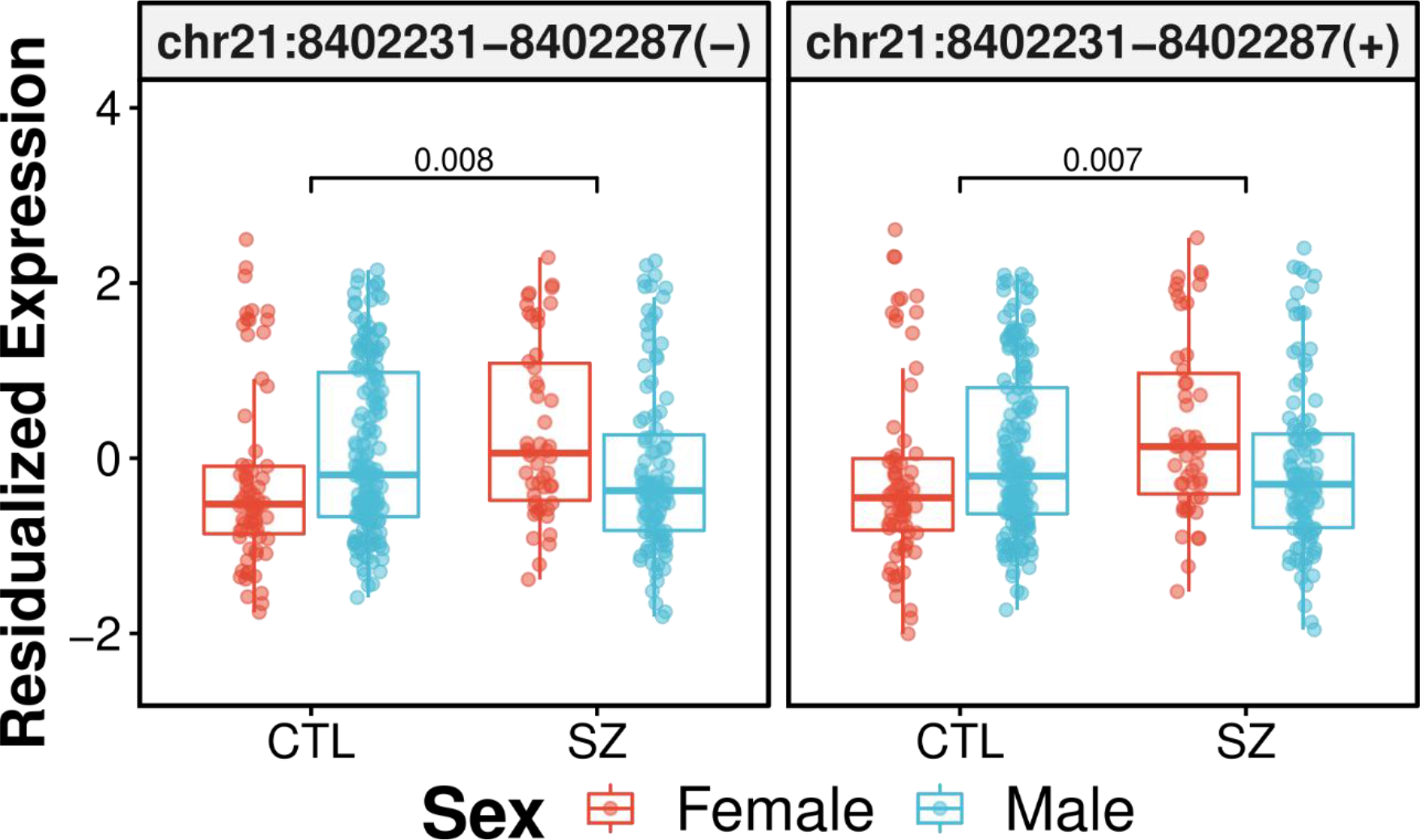
Differentially expressed junctions demonstrating sex and diagnosis interaction in the caudate nucleus. Neurotypical controls (CTL), schizophrenia (SZ), female (red), and male (blue). Adjusted p-value annotated on plot.

**Fig. S17.**
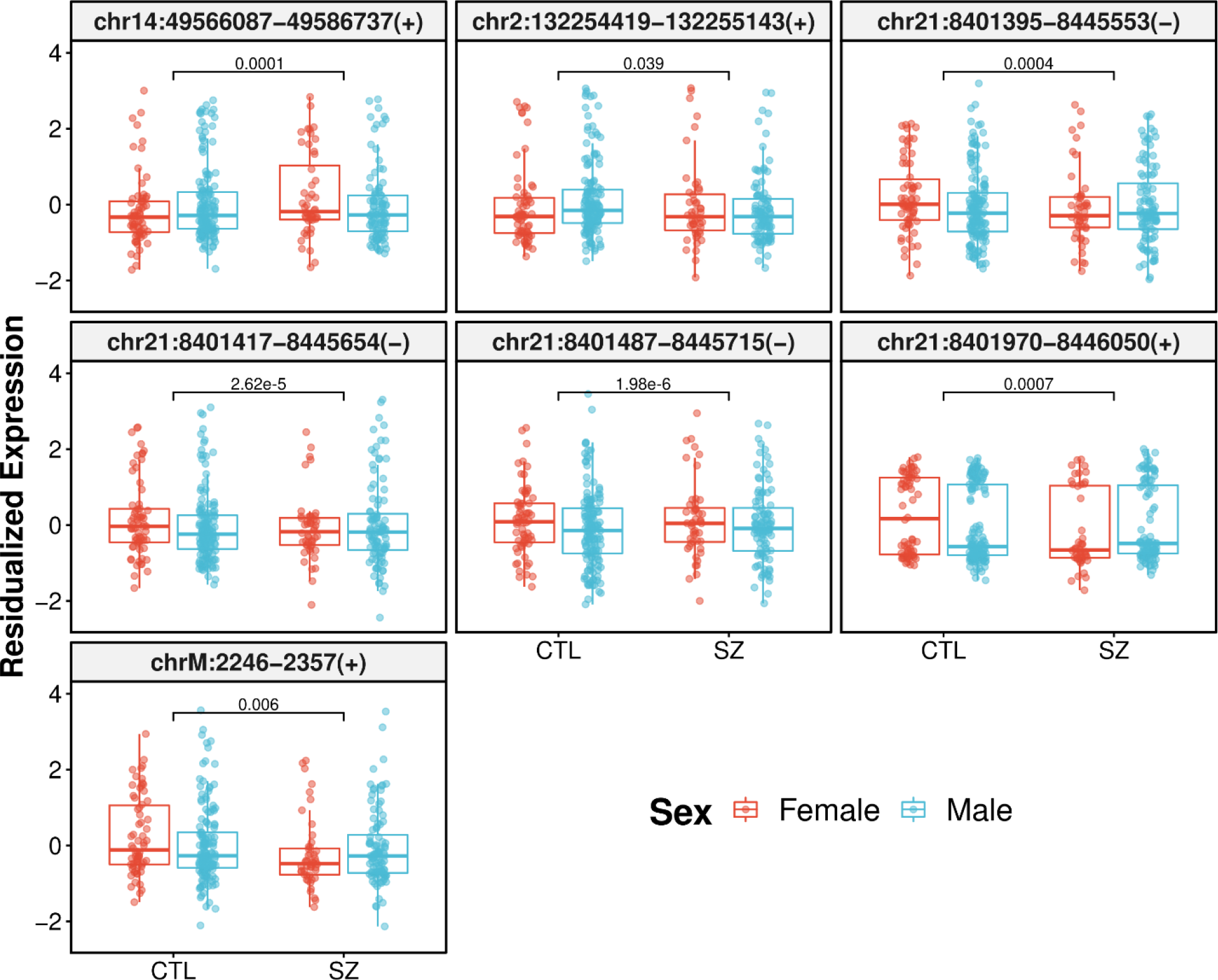
Differentially expressed junctions demonstrating sex and diagnosis interaction in DLPFC. Neurotypical controls (CTL), schizophrenia (SZ), female (red), and male (blue). Adjusted p-value annotated on plot.

**Fig. S18.**
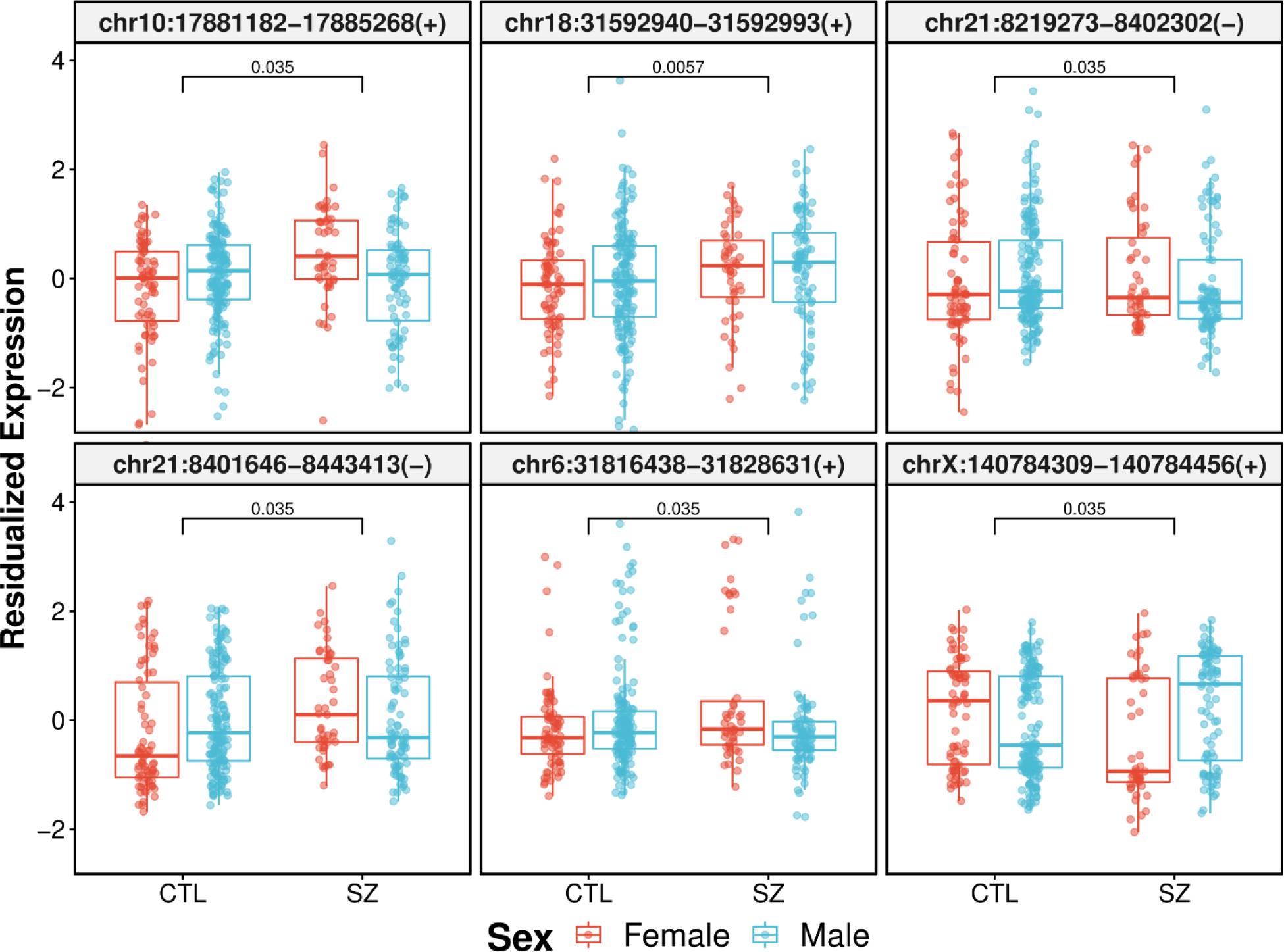
Differentially expressed junctions demonstrating sex and diagnosis interaction in the hippocampus. Neurotypical controls (CTL), schizophrenia (SZ), female (red), and male (blue). Adjusted p-value annotated on plot.

**Fig. S19.**
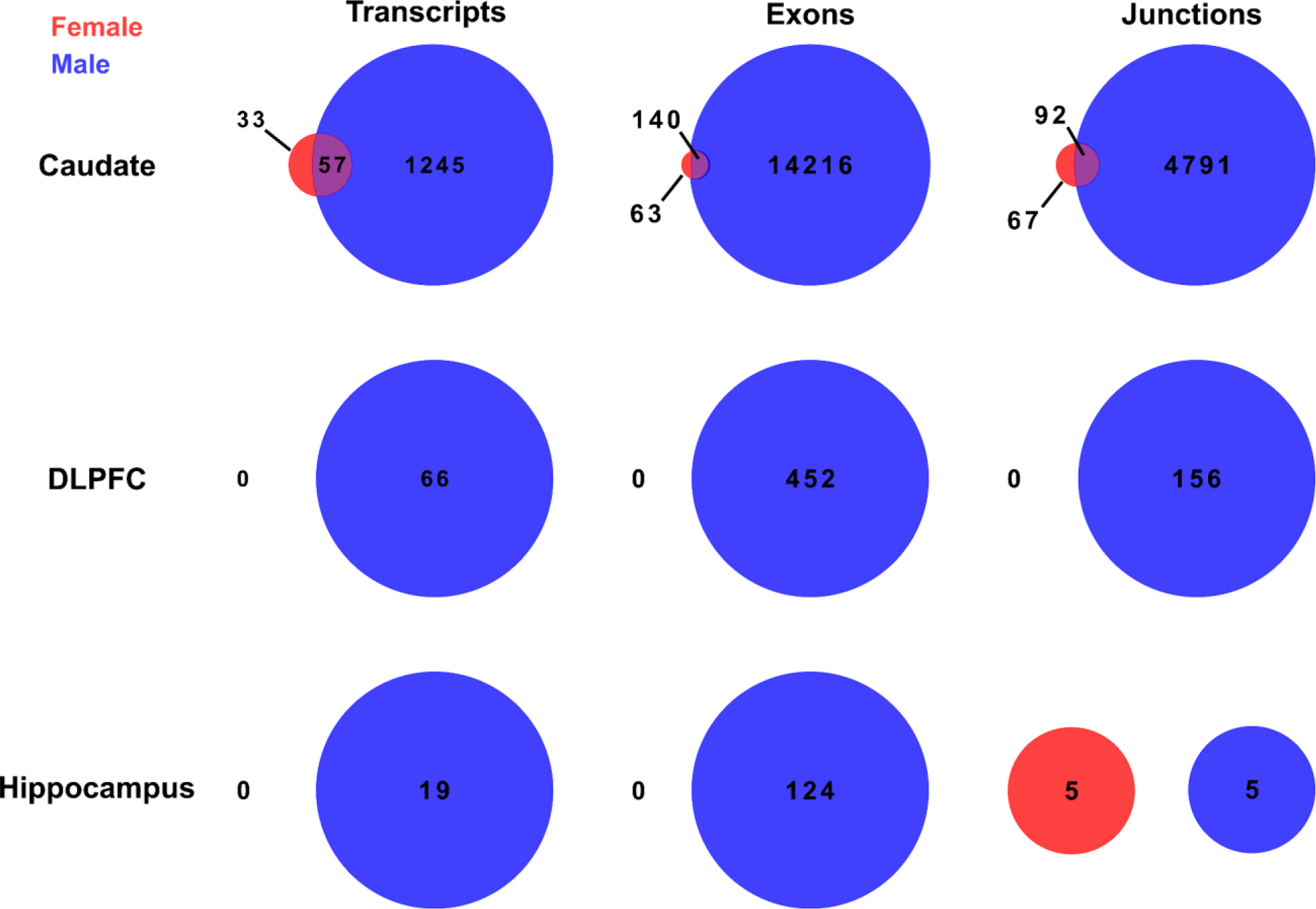
Little to no identification of female-specific schizophrenia differential features (transcripts, exons, and junctions) across the caudate nucleus, DLPFC, and hippocampus. Venn diagram showing little to no overlap for DLPFC and hippocampus between female (red) and male (blue) schizophrenia differentially expressed transcript, exon, and junction.

**Fig. S20.**
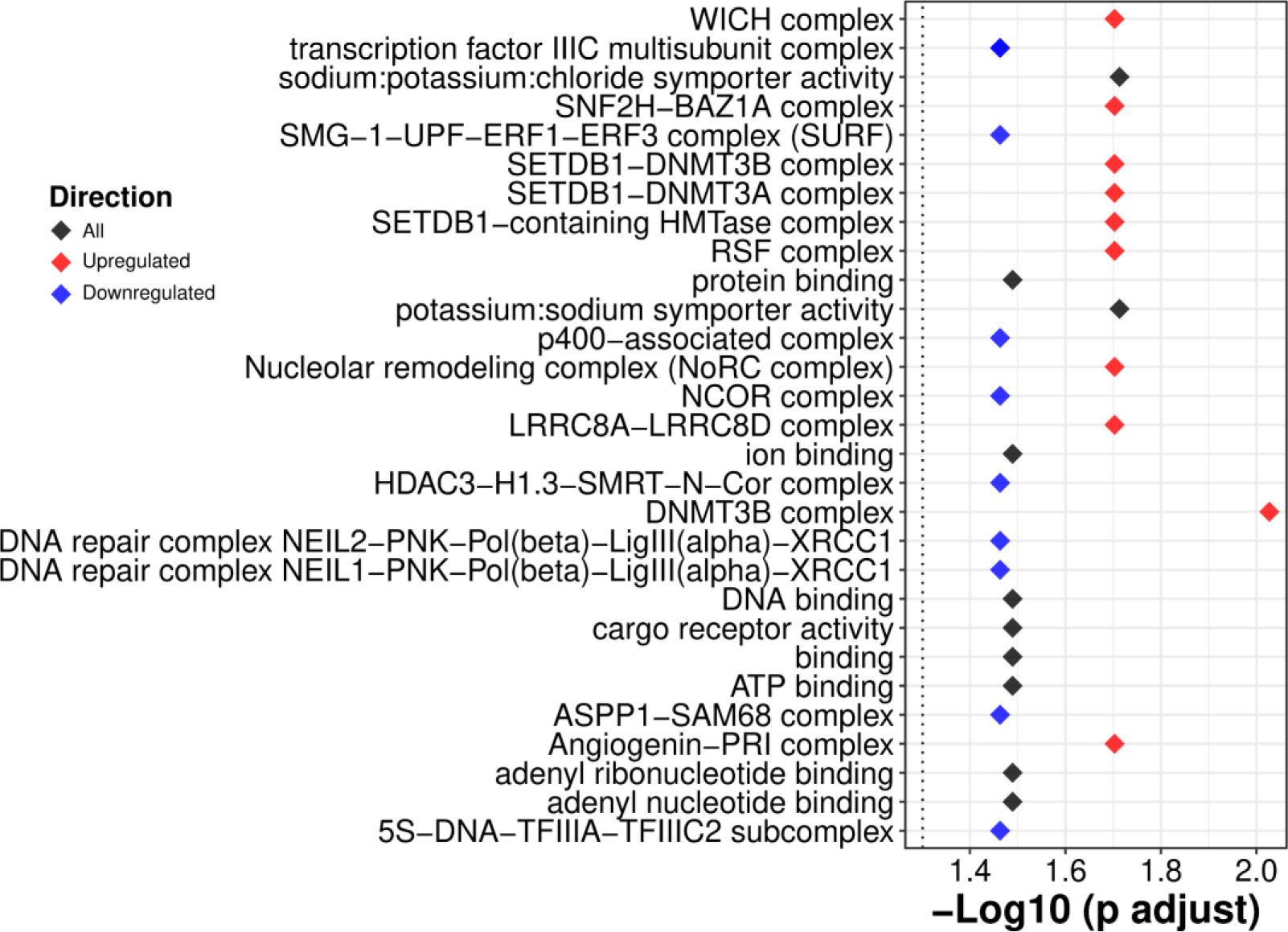
Female-specific schizophrenia DEGs for the caudate nucleus enriched in DNA repair and transcription complexes. Plot of the most significant by p-value gene terms from the gene term enrichment analysis. Black: all DEGs, red: up-regulated in schizophrenic individuals, and blue: down-regulated in schizophrenic individuals.

**Fig. S21.**
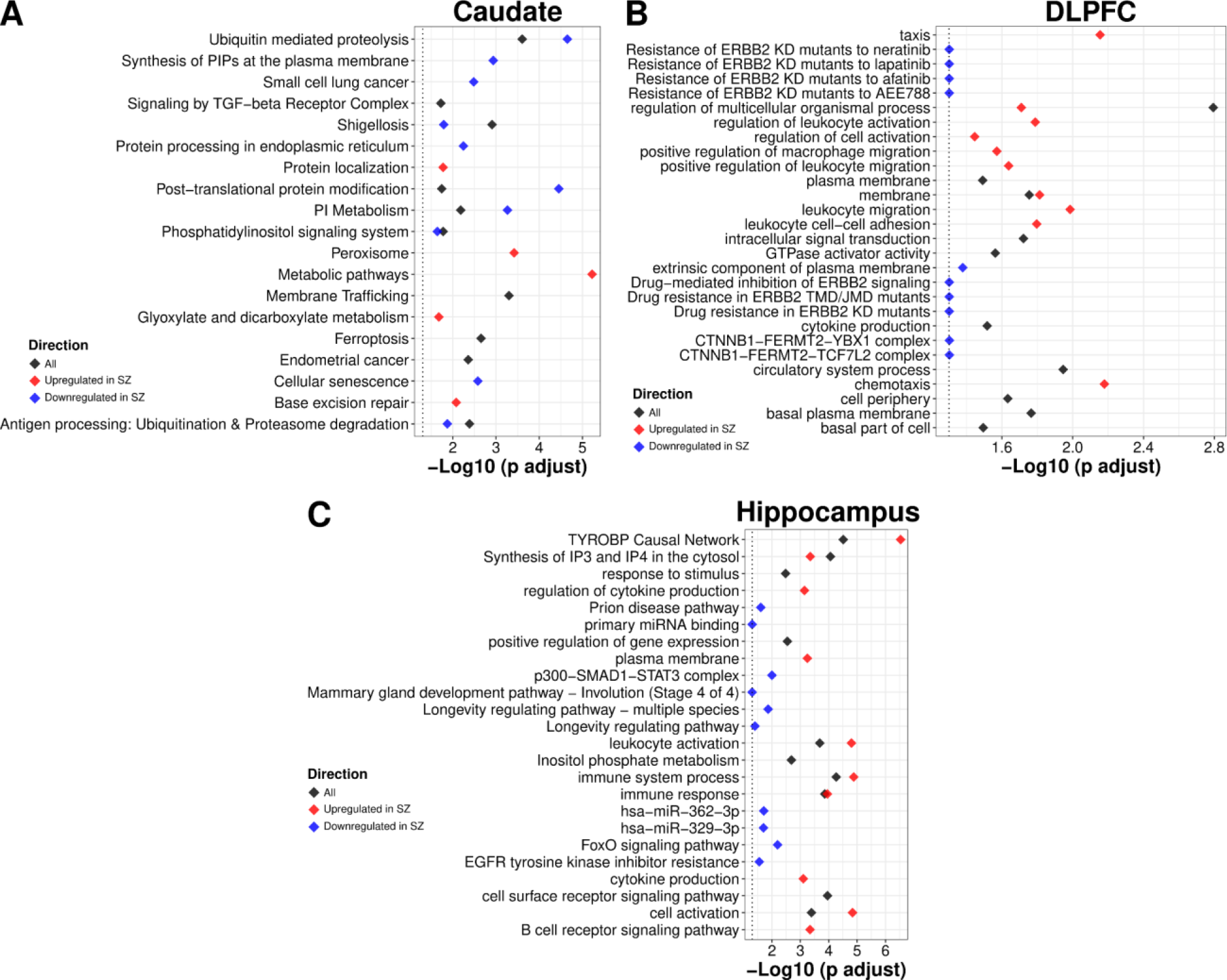
Gene term enrichment for male-specific schizophrenia DEGs across brain regions. A. The 10 most enriched KEGG pathways for the caudate nucleus. The 10 most enriched pathways (KEGG and non-KEGG) for **B.** the DLPFC and **C.** the hippocampus. Black: all DEGs for schizophrenia, red: up-regulated DEGs for schizophrenia, and blue: down-regulated DEGs for schizophrenia.

**Fig. S22.**
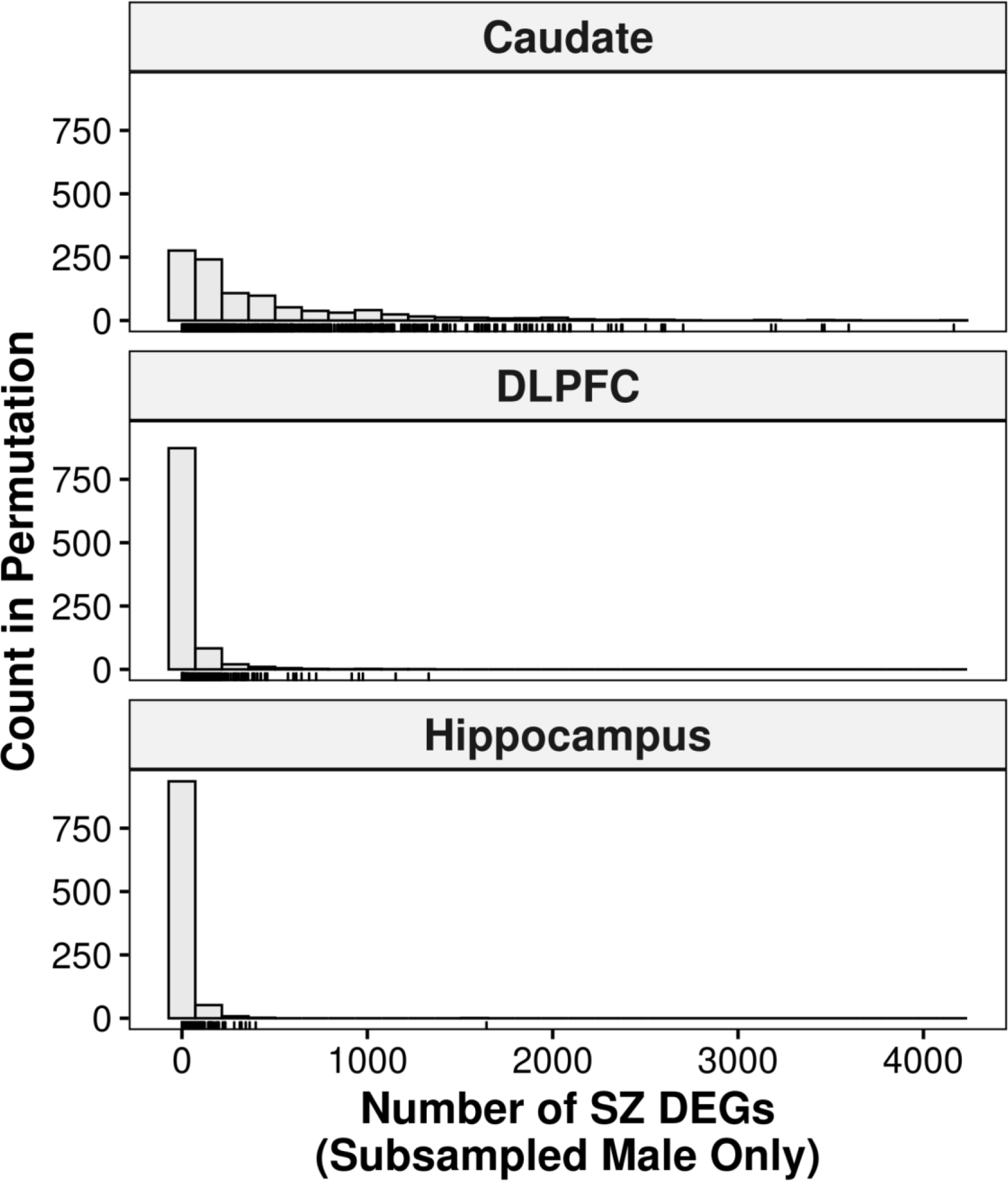
Reduction of the detected schizophrenia DEGs in male-only analysis at smaller sample sizes. Histogram of permutation analysis of male-only differential expression analysis at female sample size levels (n=121, 114, and 121 for the caudate nucleus, DLPFC, and hippocampus, respectively) shows a reduction of detected schizophrenia DEGs similar to female-only schizophrenia analysis.

**Fig. S23.**
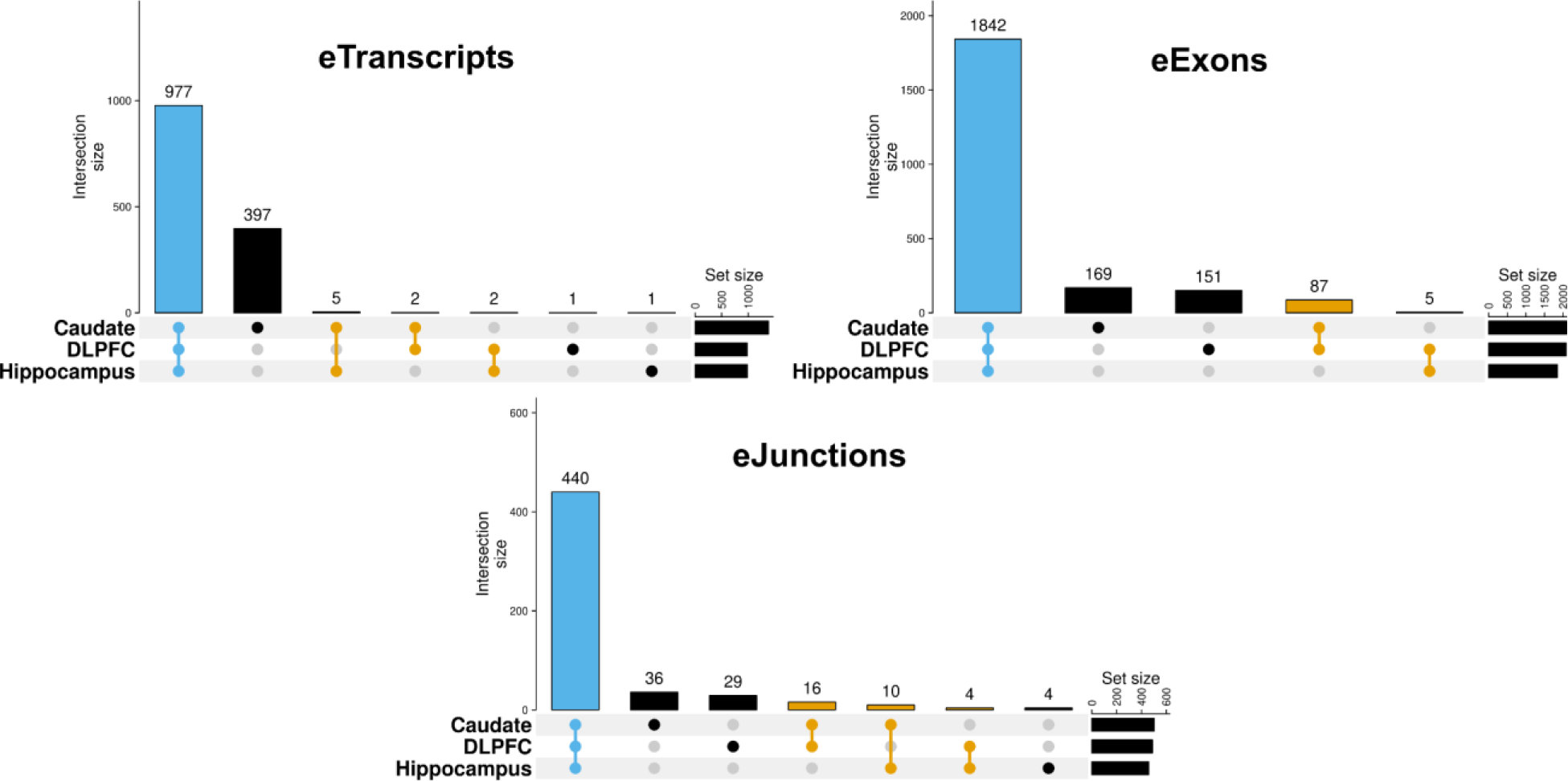
si-eQTLs are shared across brain regions. UpSet plots showing the majority of si-eQTL are shared across features for eTranscripts (si-eQTL associated with unique transcripts), eExons (si-eQTL associated with unique exons), and eJunctions (si-eQTL associated with unique junctions).

**Fig. S24.**
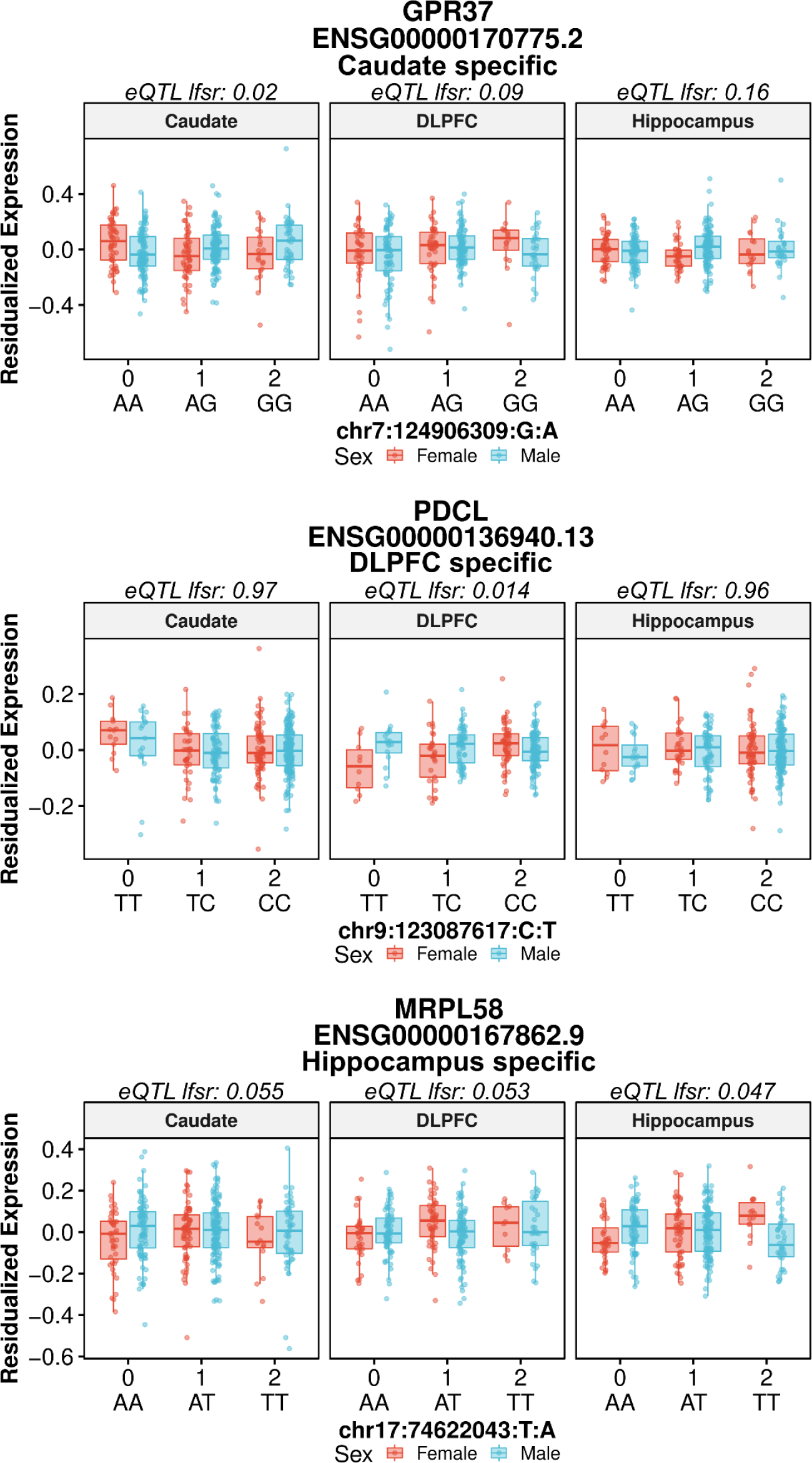
Example boxplots of si-eQTL. The most significant (local false sign rate (lfsr^72^)) si-eQTL with brain-region specific association for the caudate nucleus (top), DLPFC (middle), and hippocampus (bottom).

**Fig. S25.**
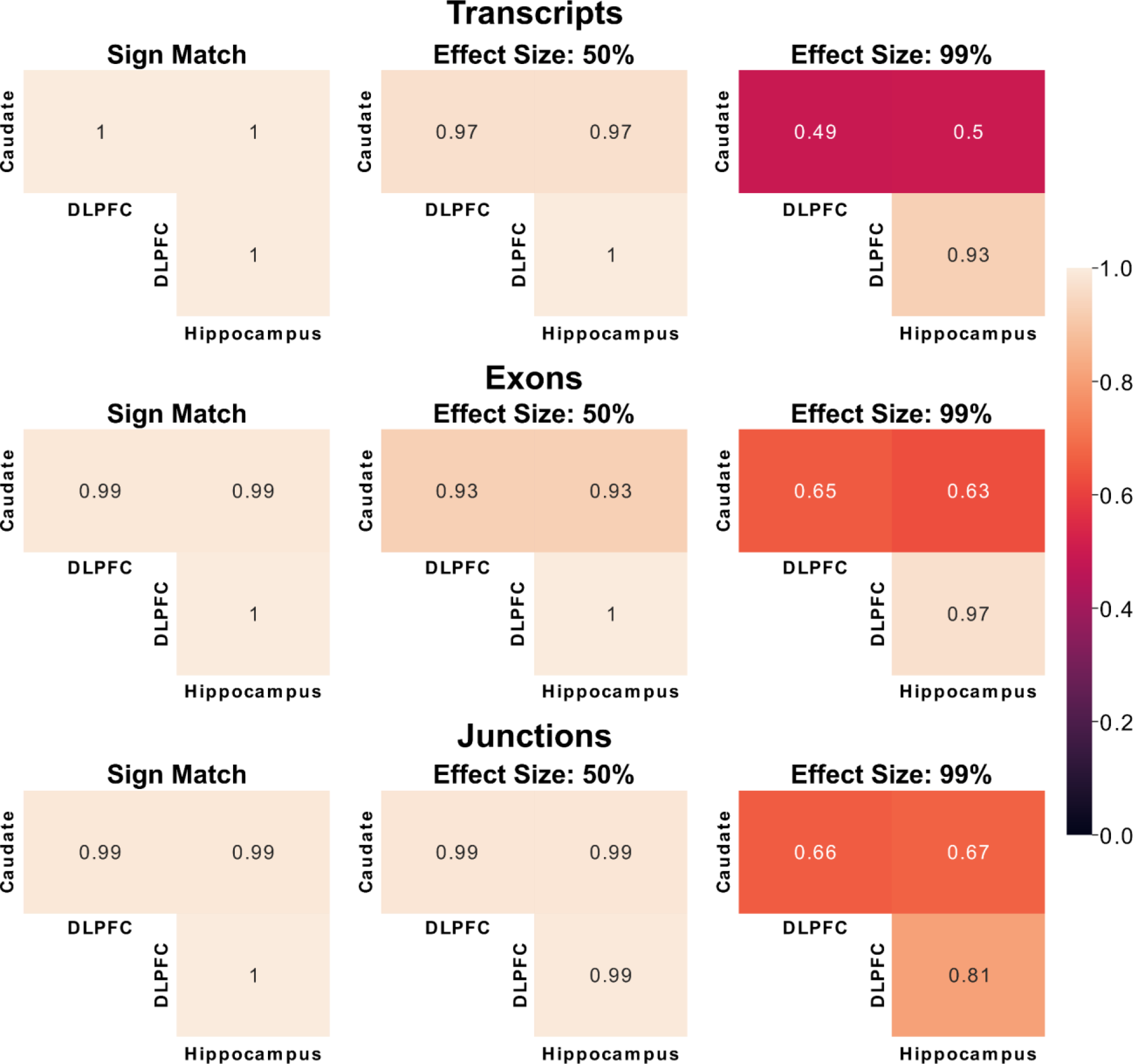
si-eQTL SNP-gene pairs shared across brain regions demonstrate concordant directionality. Heatmap of proportion of feature level (transcripts, exons, and junctions) si-eQTL sharing with sign of si-eQTL matching (left), the same sign of si-eQTL and within a factor of 0.5 effect size (middle), and the same sign of si-eQTL and within a factor 0.99 effect size (right) using mashr^48^. A factor of one is a perfect effect size match.

**Fig. S26.**
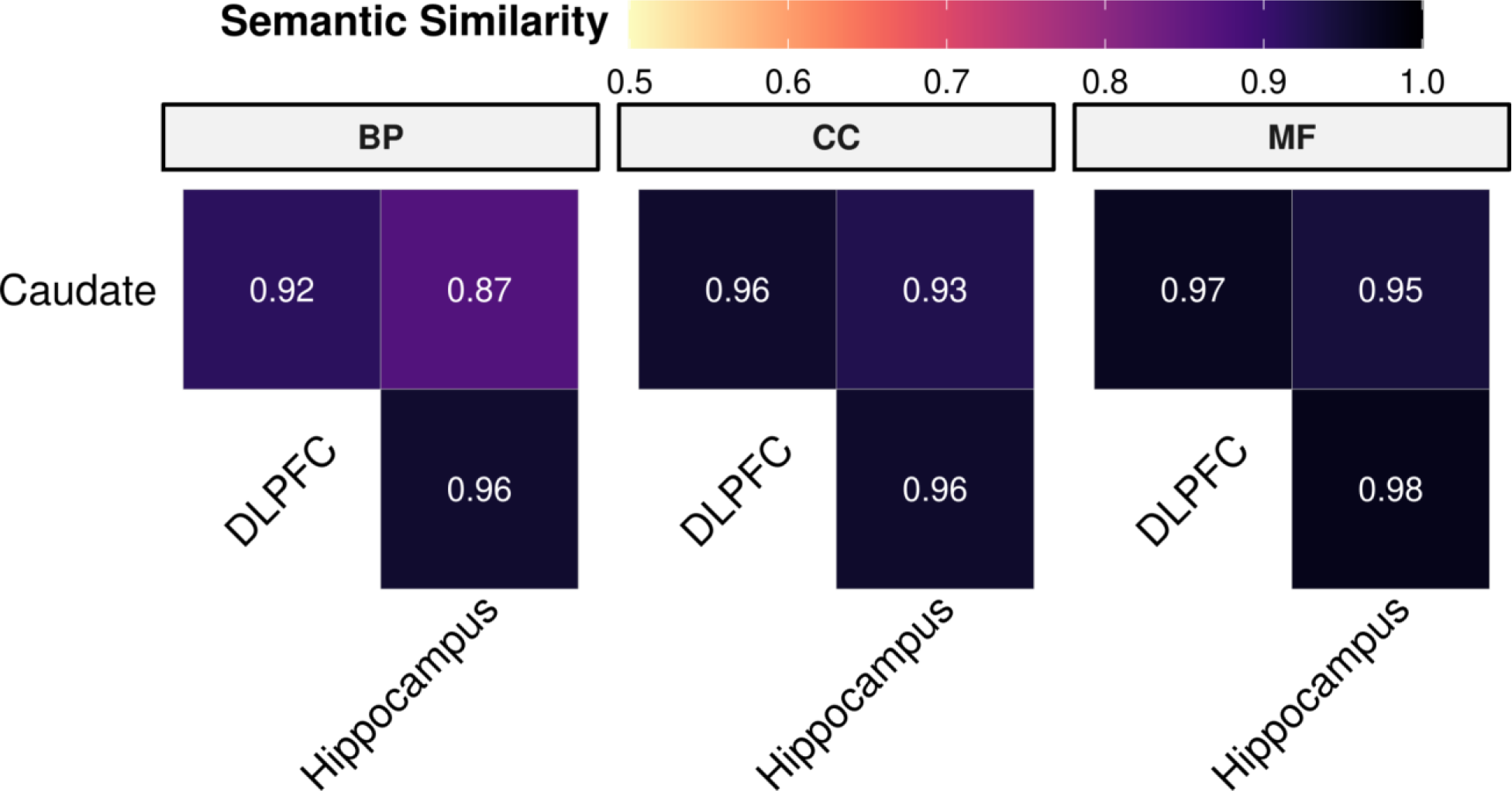
Sex-interacting eGenes share functional semantic similarity across brain regions. Heatmap of significant term enrichment for the Gene Ontology database (BP: Biological Process, CC: Cellular Component, and MF: Molecular Function) across brain regions.

**Fig. S27.**
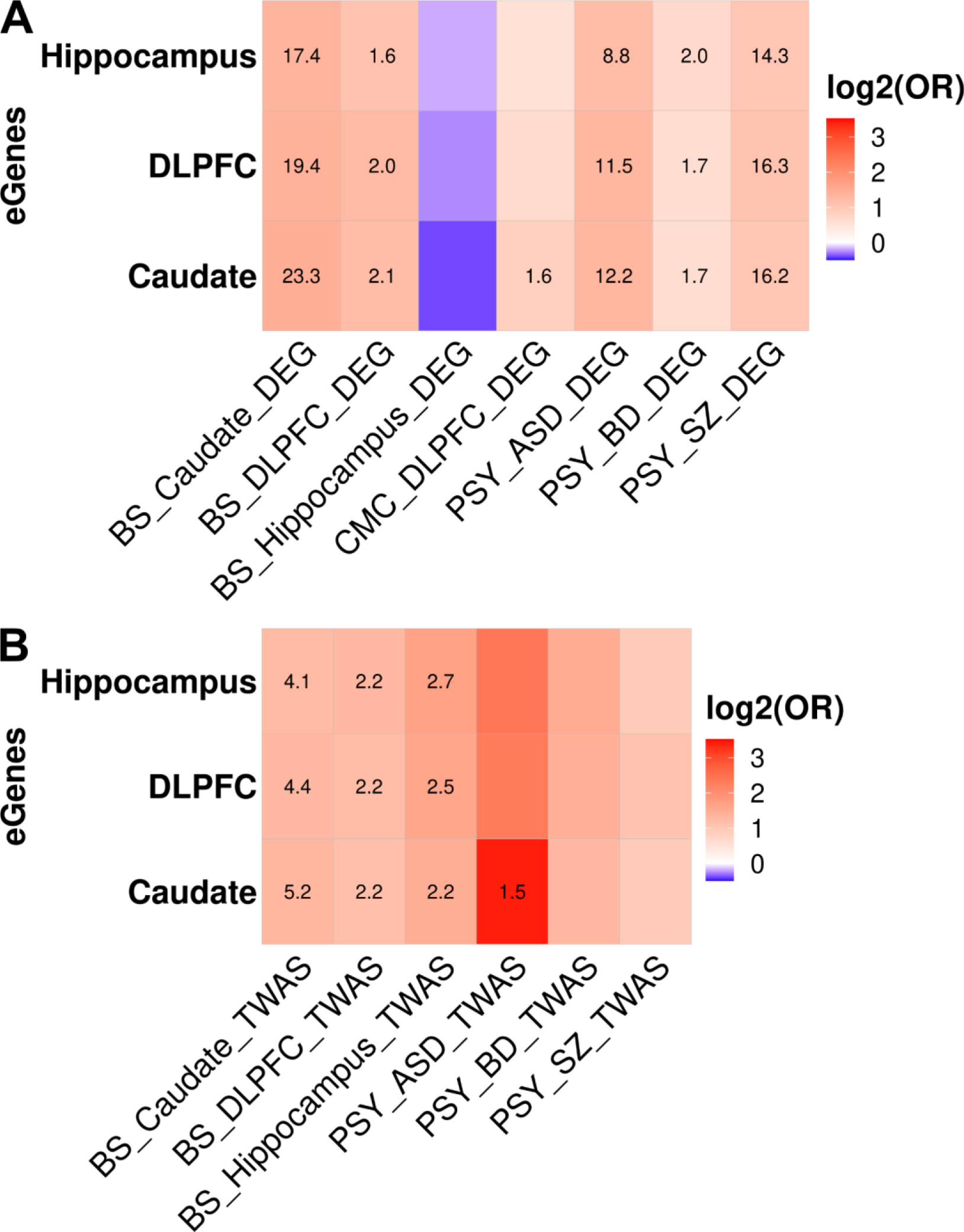
Sex-interacting eGenes are enriched for neuropsychiatric disorders. Heatmap of enrichment (red) / depletion (blue) for sex-interacting eGenes with **A.** differential expression genes (DEG) or **B.** transcriptome-wide association studies (TWAS) for neuropsychiatric disorders for the caudate nucleus, DLPFC, and hippocampus. Significant enrichments (Fisher’s exact test, FDR corrected p-values, -log10 transformed) annotated within tiles. BS: BrainSeq Consortium, CMC: CommonMind Consortium, PSY: psychENCODE. For BrainSeq Consortium and CMC, DEG and TWAS are for schizophrenia (SZ). ASD: Autism spectrum disorder, BD: Bipolar disorder. eGenes: si-eQTL associated with unique genes. psychENCODE results are for DLPFC.

**Fig. S28:**
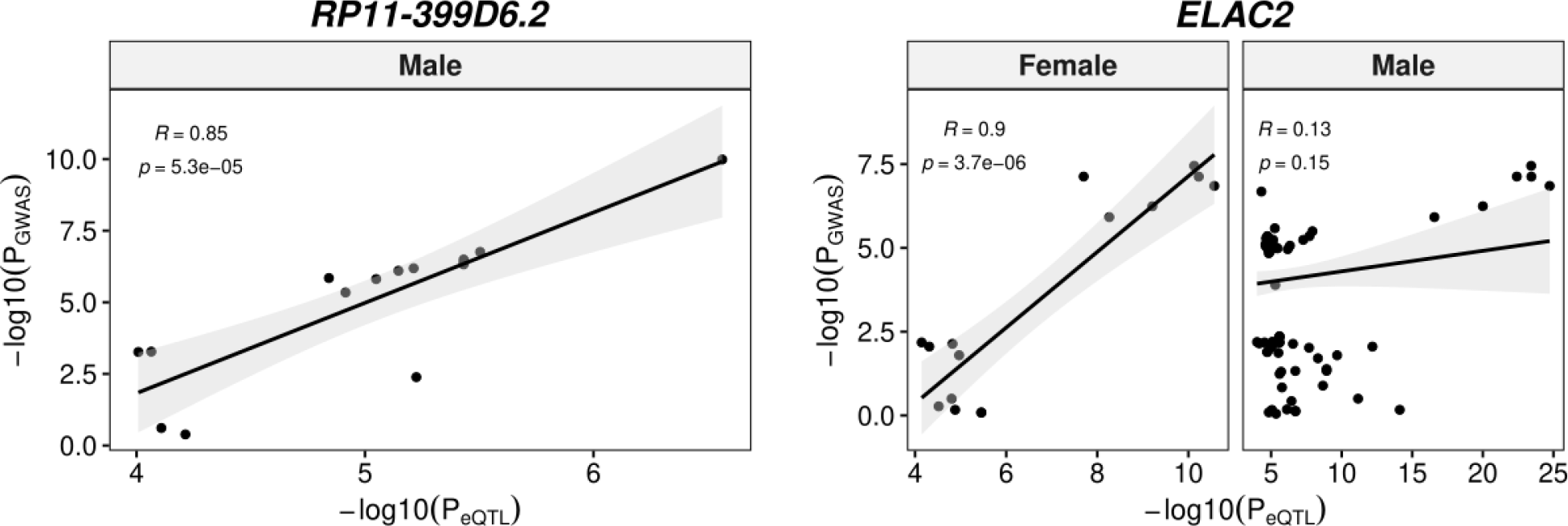
Colocalization of si-eQTL and schizophrenia risk in the caudate nucleus. P-P plots of the significant associations (RCP > 0.5).

**Fig. S29:**
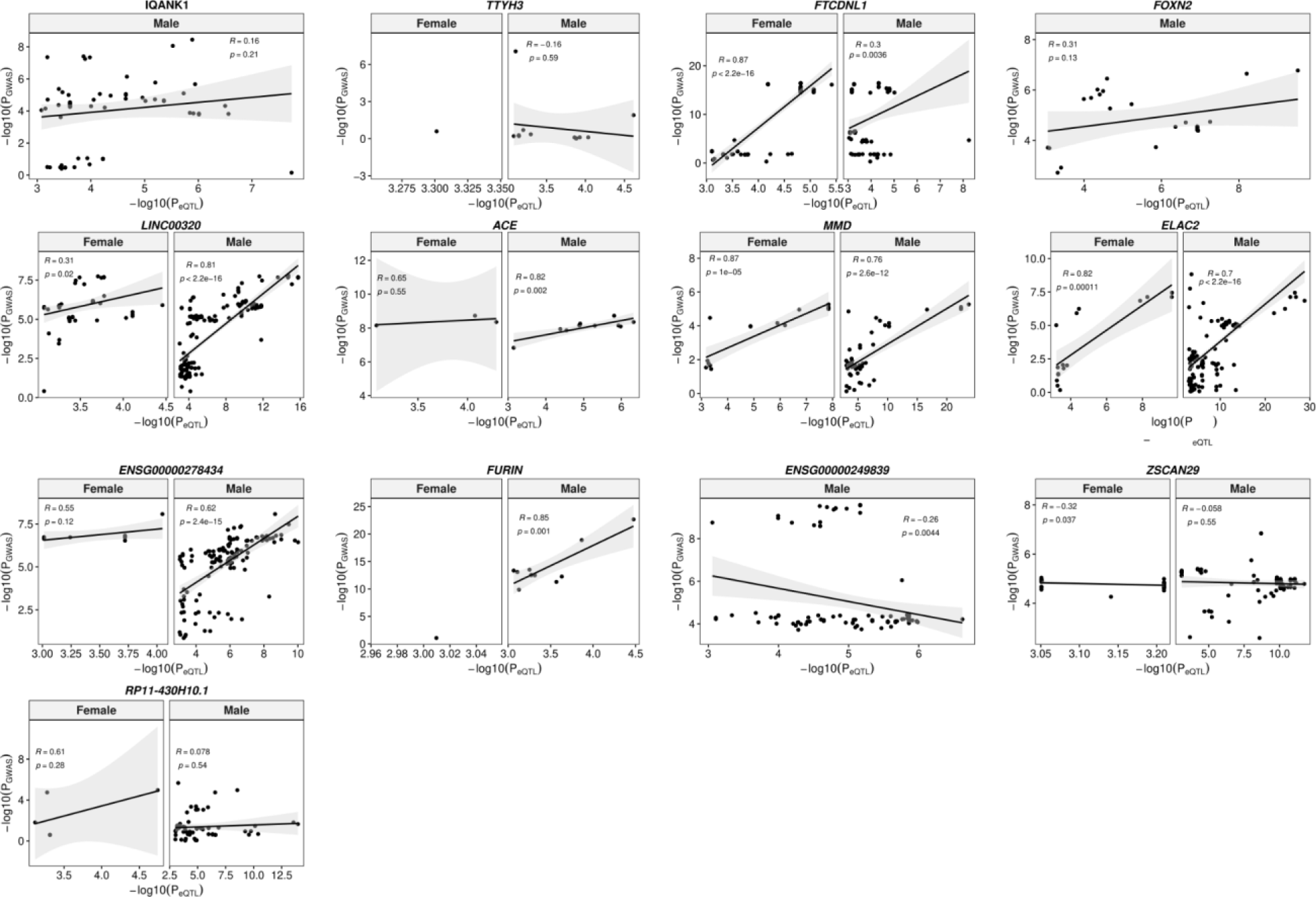
Colocalization of si-eQTL and schizophrenia risk in the DLPFC. P-P plots of the significant associations (RCP > 0.5).

**Fig. S30:**
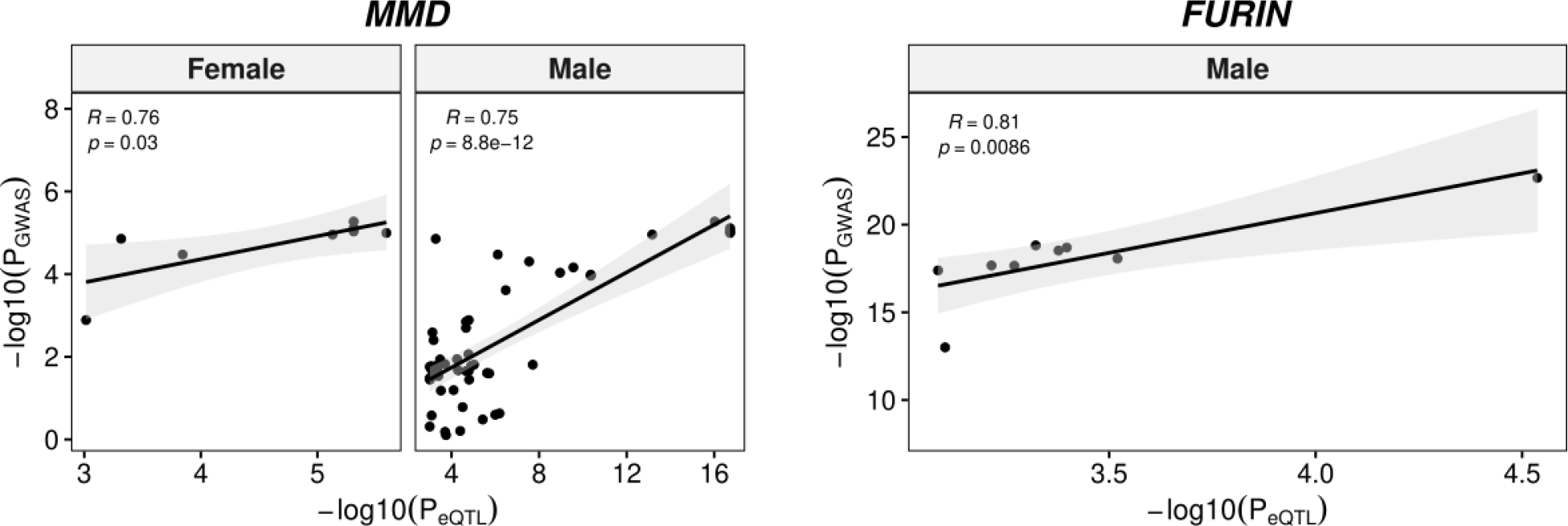
Colocalization of si-eQTL and schizophrenia risk in the hippocampus. P-P plots of the significant associations (RCP > 0.5).

### Tables

**Table S1.** Summary of differential expression results (FDR < 0.05) by feature (genes, transcripts, exons, and exon-exon junctions) for sex differences in the caudate nucleus, DLPFC, and hippocampus. Number of differentially expressed features is separated by chromosome location. The number of unique genes associated with transcript, exon, or junction in parenthesis.

| Brain Region | Chr Location | Gene | Transcript (Geneid) | Exon (Geneid) | Junction (Geneid) |
| --- | --- | --- | --- | --- | --- |
| Caudate Nucleus | Allosomal | 80 | 278 (110) | 987 (78) | 539 (53) |
|  | Autosomal | 300 | 184 (176) | 492 (189) | 236 (85) |
| DLPFC | Allosomal | 92 | 195 (73) | 1015 (100) | 564 (74) |
|  | Autosomal | 481 | 227 (288) | 2551 (801) | 1245 (626) |
| Hippocampus | Allosomal | 63 | 119 (70) | 819 (68) | 528 (50) |
|  | Autosomal | 42 | 71 (67) | 133 (46) | 218 (109) |

**Table S2.**
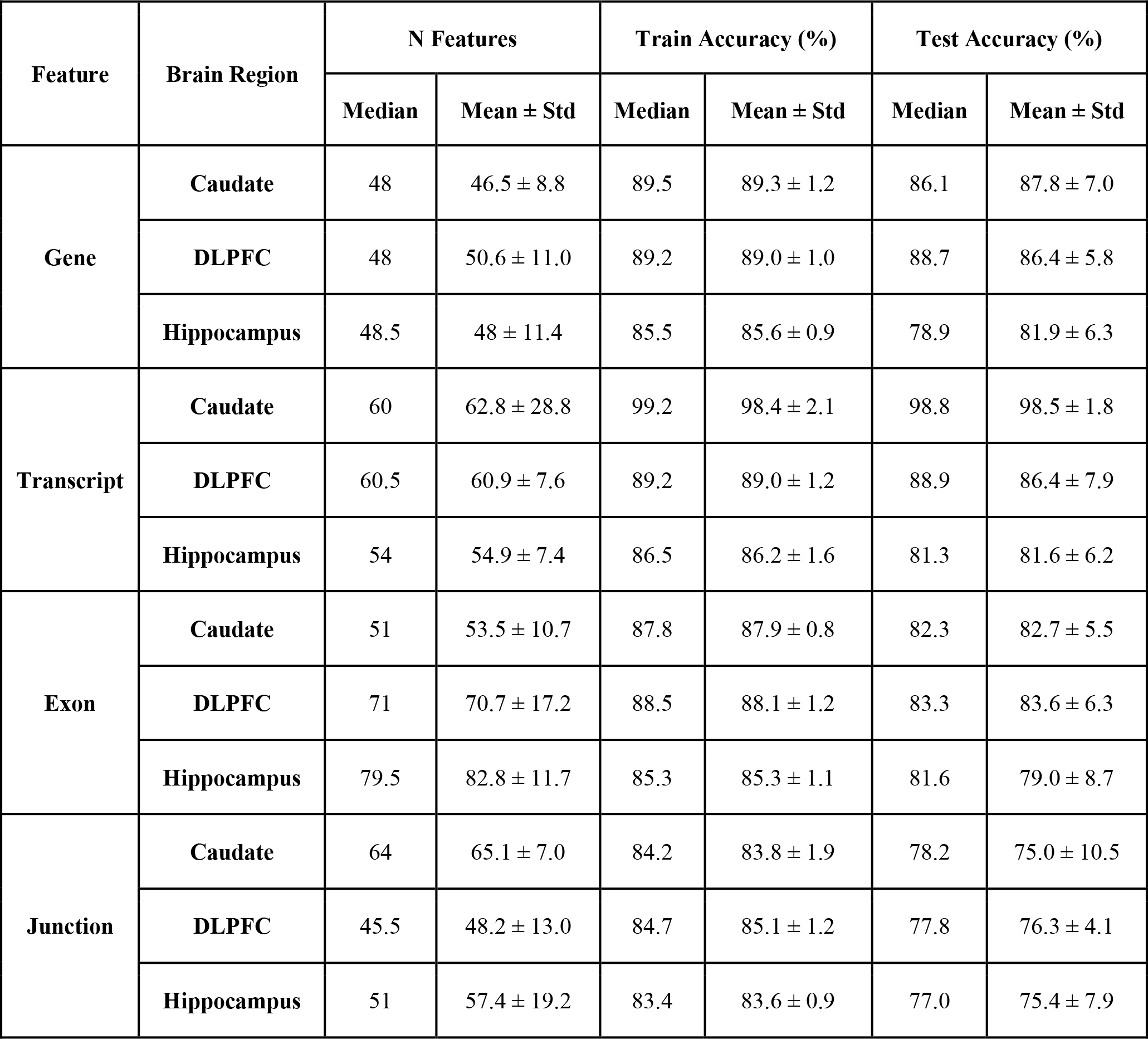
Summary of random forest classification with dynamic recursive feature elimination prediction for sex using autosomes accuracy and smallest number of features. Train, test, and smallest set metrics of mean, median, and standard deviation (std) across 10 folds for each feature (gene, transcript, exon, and junction) and brain region (caudate, DLPFC, and hippocampus).

**Table S3.** Summary results of differential expression analysis (FDR < 0.05) of interaction for sex and brain region for genes, transcripts, exons, and exon-exon junctions. The number of unique genes associated with transcript, exon, or junction in parenthesis.

| **Brain Regions** | **Gene** | **Transcript (Geneid)** | **Exon (Geneid)** | **Junction (Geneid)** |
| --- | --- | --- | --- | --- |
| Caudate Nucleus and DLPFC | 668 | 642 (569) | 2254 (649) | 324 (147) |
| Caudate Nucleus and Hippocampus | 23 | 173 (129) | 129 (38) | 47 (17) |
| DLPFC and Hippocampus | 26 | 28 (19) | 131 (24) | 58 (16) |

**Table S4.** Summary of stringent sex-specific differential expression analysis (FDR < 0.05) for schizophrenia by sex and feature (gene, transcript, exon, and exon-exon junction) for the caudate nucleus, DLPFC, and hippocampus. The number of unique genes associated with transcript, exon, or junction in parenthesis.

| Brain Region | Sex | Gene | Transcript (Geneid) | Exon (Geneid) | Junction (Geneid) |
| --- | --- | --- | --- | --- | --- |
| Caudate Nucleus | Female | 124 | 11 (7) | 19 (9) | 17 (11) |
|  | Male | 1858 | 480 (436) | 5834 (1767) | 2064 (1073) |
| DLPFC | Female | 0 | 0 (0) | 0 (0) | 0 (0) |
|  | Male | 122 | 22 (22) | 116 (80) | 49 (41) |
| Hippocampus | Female | 0 | 0 (0) | 0 (0) | 5 (3) |
|  | Male | 104 | 17 (15) | 113 (50) | 3 (3) |

**Table S5.** Summary of shared genes between two brain regions for stringent male-specific schizophrenia DEGs. The genes shared across brain regions are bolded.

| Brain Regions | # of Genes | Gene Names |
| --- | --- | --- |
| Caudate Nucleus and DLPFC | 14 | *BCO1*, ***CARD11***, *PTGS1*, *RP11-214K3.18*, *DPY19L2*, *FCGR3B*, ***RP11-489O18.1***, *STXBP6*, *CNTNAP3*, *PALD1*, *CARTPT*, *MAF*, *ECT2L*, and *DOCK8* |
| Caudate Nucleus and Hippocampus | 14 | ***RP11-489O18.1***, ***CARD11***, *SUSD3*, *OSMR-AS1*, *SELPLG*, *AC008697.1*, *IGSF6*, *ADGRG5*, *GCKR*, *USE1*, *KIAA0040*, *UBR5*, *TRAF3IP3*, and *RP11-214K3.22* |
| DLPFC and Hippocampus | 7 | *TNFRSF13C*, ***CARD11***, *GPR34*, ***RP11-489O18.1***, *CLEC7A*, *LPAR5*, and *CSF1R* |

### Data

**Data S1. BrainSeq_bySex_genes_3region_eQTL_results.tar.gz:** Compressed tar file containing gene eQTL results by sex per brain region including all nominal p-values as well as permutation analysis.

**Data S2. differential_expression_analysis_4features_sex.txt.gz**: Compressed text file of differential expression analysis for sex across the caudate nucleus, DLPFC, and hippocampus for four features (gene, transcript, exon, and junction).

**Data S3. functional_enrichment_analysis_3brain_regions_sex.txt**: Text file of functional enrichment analysis for sex across the caudate nucleus, DLPFC, and hippocampus, separated by sex bias (all, female bias, male bias).

**Data S4. BrainSeq_sex_WGCNA_results.tar.gz**: Compressed tar file containing with WGCNA results for autosomal only and all genes sex networks including eigengenes, module membership text files, GO enrichment text and excel file results, enrichment results with sex DEG analysis for the caudate nucleus, DLPFC, and hippocampus.

**Data S5. differential_expression_region_interaction_sex_4features.txt**: Text file of differential expression analysis for the interaction between brain region and sex for the four features (gene, transcript, exon, and junction).

**Data S6. BrainSeq_male_biased_genes_XCI_status.tsv**: Text file of differentially expressed male-biased (up-regulated in male individuals) genes across the caudate nucleus, DLPFC, and hippocampus annotated for X-chromosome inactivation (XCI) status.

**Data S7. differential_expression_schizophrenia_by_sex_4features.txt.gz**: Compressed text file of differential expression analysis for schizophrenia by sex across the caudate nucleus, DLPFC, and hippocampus for four features (gene, transcript, exon, and junction).

**Data S8. functional_enrichment_analysis_maleSZ_3brain_regions.txt**: Functional enrichment of male-specific schizophrenia across three brain regions using stringent gene list (all DEGs, up- and down-regulated in schizophrenia).

**Data S9. BrainSeq_sexGenotypes_4features_3regions.txt.gz**: Compressed text file of sex-interacting eQTL results across the caudate nucleus, DLPFC, and hippocampus for four features (gene, transcript, exon, and junction) generated using mash modeling.

**Data S10. BrainSeq_sex_interacting_eGene_functional_enrichment.xlsx**: Excel file of functional enrichment of sex-interacting eQTL associated with unique genes for the caudate nucleus, DLPFC, and hippocampus.

**Data S11. BrainSeq_siEQTL_public_comparison.xlsx**: Excel file of si-eQTL shared with the public datasets^8, 18^.

**Data S12. dapg_sex_interacting_eQTL_results.tar.gz**: si-eQTL results after fine-mapping across the three brain regions for four features (gene, transcript, exon, and junction).

**Data S13. BrainSeq_colocalization_3regions.xlsx**: Excel file of gene level colocalization between dap-g fine-mapped sex-interacting eQTL and schizophrenia GWAS^49^ on the signal level and individual SNP level using fastENLOC.

**Data S14. BrainSeq_colocalization_eQTpLoTs.tar.gz**: Compressed text file of colocalization plots (eQTpLot) between gene-level, sex-only eQTL analysis and schizophrenia risk (PGC3^49^, p-value < 5e-8) for the caudate nucleus, DLPFC, and hippocampus.

