## Supplementary material for "Transcriptional and genetic sex differences for schizophrenia across the dorsolateral prefrontal cortex, hippocampus, and caudate nucleus": Data S4: module_trait_relationships.pdf

### Module kME−Trait Correlation

| | Sex | Schizo | Age | mitoRate | rRNA_rate | totalAssignedGene | RIN | overallMapRate | snpPC1 | snpPC2 | snpPC3 | sv1 | sv2 | sv3 | sv4 |
|---|---|---|---|---|---|---|---|---|---|---|---|---|---|---|---|
| MEpink | 0.032 (0.5) | 0.0064 (0.9) | −0.0072 (0.9) | −0.0022 (1) | −0.013 (0.8) | 0.034 (0.5) | 0.027 (0.6) | 0.028 (0.6) | −0.00027 (1) | 0.0065 (0.9) | −0.0015 (1) | −0.041 (0.4) | 0.0056 (0.9) | 0.0035 (0.9) | −0.0074 (0.9) |
| MEred | 0.034 (0.5) | 0.0031 (1) | −0.016 (0.8) | −0.0097 (0.9) | −0.002 (1) | 0.025 (0.6) | 0.018 (0.7) | 0.018 (0.7) | 0.0069 (0.9) | 0.0098 (0.8) | 0.003 (1) | −0.036 (0.5) | 0.0023 (1) | 0.00063 (1) | 0.0033 (0.9) |
| MEblue | 0.019 (0.7) | −0.0023 (1) | −0.02 (0.7) | −0.011 (0.8) | 0.014 (0.8) | 0.0094 (0.9) | 0.0047 (0.9) | 0.0018 (1) | 0.012 (0.8) | 0.0071 (0.9) | 0.0076 (0.9) | −0.019 (0.7) | 0.0098 (0.8) | −0.00056 (1) | 0.018 (0.7) |
| MEpurple | 0.019 (0.7) | −0.0094 (0.9) | −0.026 (0.6) | −0.033 (0.5) | 0.018 (0.7) | 0.01 (0.8) | −0.0015 (1) | 0.025 (0.6) | 0.025 (0.6) | 0.0046 (0.9) | 0.014 (0.8) | −0.037 (0.5) | −0.0024 (1) | −0.0013 (1) | 0.031 (0.5) |
| MEbrown | −0.073 (0.2) | −0.025 (0.6) | −0.00029 (1) | −0.081 (0.1) | −0.0071 (0.9) | 0.016 (0.8) | 0.029 (0.6) | 0.078 (0.1) | 0.0041 (0.9) | 0.0094 (0.9) | −0.008 (0.9) | −0.067 (0.2) | −0.066 (0.2) | 0.022 (0.7) | −0.03 (0.6) |
| MElightyellow | −0.046 (0.4) | −0.027 (0.6) | −0.005 (0.9) | −0.12 (0.02) | −0.027 (0.6) | 0.047 (0.4) | 0.072 (0.2) | 0.14 (0.008) | 0.025 (0.6) | 0.015 (0.8) | 0.01 (0.8) | −0.13 (0.01) | −0.096 (0.06) | 0.037 (0.5) | −0.025 (0.6) |
| MEdarkgrey | −0.026 (0.6) | −0.016 (0.7) | −0.015 (0.8) | −0.046 (0.4) | 0.022 (0.7) | −0.0056 (0.9) | 0.023 (0.6) | 0.011 (0.8) | 0.021 (0.7) | 0.0029 (1) | 0.016 (0.8) | −0.019 (0.7) | −0.012 (0.8) | 0.028 (0.6) | 0.027 (0.6) |
| MEdarkorange | −0.15 (0.003) | −0.0038 (0.9) | 0.0021 (1) | −0.04 (0.4) | −0.0062 (0.9) | 0.0074 (0.9) | 0.021 (0.7) | 0.048 (0.4) | −0.0032 (1) | −0.0043 (0.9) | −0.0093 (0.9) | −0.041 (0.4) | −0.048 (0.3) | 0.055 (0.3) | −0.01 (0.8) |
| MEblack | −0.046 (0.4) | −0.021 (0.7) | −0.0075 (0.9) | −0.069 (0.2) | 0.0074 (0.9) | −0.0021 (1) | 0.03 (0.6) | 0.039 (0.5) | 0.025 (0.6) | 0.00089 (1) | 0.024 (0.6) | −0.028 (0.6) | −0.012 (0.8) | 0.011 (0.8) | 0.035 (0.5) |
| MElightgreen | −0.04 (0.4) | −0.017 (0.7) | 0.012 (0.8) | −0.12 (0.02) | −0.022 (0.7) | 0.021 (0.7) | 0.076 (0.1) | 0.1 (0.04) | 0.022 (0.7) | −0.00046 (1) | 0.023 (0.7) | −0.078 (0.1) | −0.05 (0.3) | 0.011 (0.8) | 0.021 (0.7) |
| MEmidnightblue | −0.013 (0.8) | 0.00088 (1) | 0.016 (0.8) | −0.054 (0.3) | −0.009 (0.9) | −0.013 (0.8) | 0.033 (0.5) | 0.019 (0.7) | 0.012 (0.8) | −0.018 (0.7) | 0.02 (0.7) | 0.011 (0.8) | 0.021 (0.7) | −0.017 (0.7) | 0.064 (0.2) |
| MEdarkred | −0.025 (0.6) | −0.014 (0.8) | 0.012 (0.8) | −0.039 (0.4) | −0.0015 (1) | −0.0019 (1) | 0.005 (0.9) | 0.049 (0.3) | −0.0066 (0.9) | −0.0052 (0.9) | −0.016 (0.7) | −0.025 (0.6) | −0.052 (0.3) | 0.031 (0.5) | −0.031 (0.5) |
| MEyellow | 0.0058 (0.9) | −0.007 (0.9) | 0.012 (0.8) | −0.028 (0.6) | 0.0016 (1) | −0.017 (0.7) | 0.006 (0.9) | 0.0093 (0.9) | 0.0026 (1) | −0.016 (0.7) | 0.00033 (1) | 0.015 (0.8) | 0.0018 (1) | −0.00014 (1) | 0.025 (0.6) |
| MEskyblue | −0.06 (0.2) | −0.014 (0.8) | 0.0049 (0.9) | −0.022 (0.7) | −0.0089 (0.9) | 0.0086 (0.9) | 0.0021 (1) | 0.043 (0.4) | −0.00015 (1) | 0.0059 (0.9) | −0.011 (0.8) | −0.034 (0.5) | −0.061 (0.2) | 0.04 (0.4) | −0.045 (0.4) |
| MEtan | 0.029 (0.6) | −0.0035 (0.9) | 0.0016 (1) | −0.05 (0.3) | −0.02 (0.7) | 0.043 (0.4) | 0.039 (0.5) | 0.051 (0.3) | 0.0067 (0.9) | −0.008 (0.9) | 0.003 (1) | −0.053 (0.3) | −0.0014 (1) | 0.025 (0.6) | 0.00052 (1) |
| MEgrey60 | 0.056 (0.3) | −0.0062 (0.9) | −0.021 (0.7) | −0.059 (0.3) | −0.019 (0.7) | 0.06 (0.2) | 0.052 (0.3) | 0.068 (0.2) | 0.011 (0.8) | 0.0037 (0.9) | 0.011 (0.8) | −0.083 (0.1) | −0.0027 (1) | 0.044 (0.4) | 0.0038 (0.9) |
| MEsaddlebrown | −0.00039 (1) | −0.028 (0.6) | −0.023 (0.7) | −0.11 (0.03) | −0.0031 (1) | 0.025 (0.6) | 0.053 (0.3) | 0.1 (0.05) | 0.029 (0.6) | 0.0068 (0.9) | 0.011 (0.8) | −0.092 (0.07) | −0.057 (0.3) | 0.034 (0.5) | 0.018 (0.7) |
| MEsalmon | −0.046 (0.4) | −0.014 (0.8) | 0.0062 (0.9) | 0.059 (0.3) | 0.036 (0.5) | −0.046 (0.4) | −0.046 (0.4) | −0.085 (0.1) | −0.017 (0.7) | −0.013 (0.8) | −0.028 (0.6) | 0.074 (0.1) | 0.022 (0.7) | −0.0089 (0.9) | 0.0013 (1) |
| MEdarkgreen | 0.039 (0.4) | 0.028 (0.6) | 0.012 (0.8) | 0.067 (0.2) | 0.011 (0.8) | −0.033 (0.5) | −0.039 (0.4) | −0.09 (0.08) | −0.02 (0.7) | −0.024 (0.6) | 0.0026 (1) | 0.099 (0.05) | 0.097 (0.06) | −0.044 (0.4) | 0.053 (0.3) |
| MEorange | 0.0025 (1) | 4.4e−05 (1) | 0.018 (0.7) | 0.011 (0.8) | 0.0082 (0.9) | −0.035 (0.5) | −0.0087 (0.9) | −0.04 (0.4) | −0.016 (0.8) | −0.022 (0.7) | −0.012 (0.8) | 0.059 (0.3) | 0.027 (0.6) | −0.0085 (0.9) | 0.029 (0.6) |
| MEviolet | 0.048 (0.3) | 0.038 (0.5) | 0.0027 (1) | 0.033 (0.5) | −0.014 (0.8) | 0.023 (0.7) | 0.0041 (0.9) | −0.035 (0.5) | 0.006 (0.9) | −0.014 (0.8) | 0.043 (0.4) | 0.029 (0.6) | 0.11 (0.03) | −0.058 (0.3) | 0.073 (0.2) |
| MEwhite | 0.061 (0.2) | 0.042 (0.4) | 0.0033 (0.9) | 0.11 (0.03) | 0.0086 (0.9) | −0.0042 (0.9) | −0.058 (0.3) | −0.1 (0.05) | −0.026 (0.6) | −0.013 (0.8) | 0.0032 (0.9) | 0.087 (0.09) | 0.12 (0.02) | −0.06 (0.2) | 0.024 (0.6) |
| MEgreen | 0.063 (0.2) | 0.035 (0.5) | −0.0056 (0.9) | 0.12 (0.02) | 0.038 (0.5) | −0.038 (0.5) | −0.11 (0.03) | −0.15 (0.004) | −0.005 (0.9) | −0.023 (0.7) | 0.024 (0.6) | 0.13 (0.009) | 0.16 (0.001) | −0.11 (0.04) | 0.088 (0.09) |
| MEgreenyellow | 0.085 (0.1) | 0.031 (0.5) | −0.012 (0.8) | 0.099 (0.05) | 0.04 (0.4) | −0.034 (0.5) | −0.086 (0.09) | −0.12 (0.02) | −0.0098 (0.8) | −0.014 (0.8) | 0.0034 (0.9) | 0.11 (0.03) | 0.12 (0.02) | −0.065 (0.2) | 0.069 (0.2) |
| MElightcyan | 0.028 (0.6) | −0.02 (0.7) | 0.0072 (0.9) | 0.0016 (1) | −0.04 (0.4) | 0.055 (0.3) | −0.011 (0.8) | 0.093 (0.07) | −0.0024 (1) | −0.0039 (0.9) | −0.039 (0.4) | −0.079 (0.1) | 0.0089 (0.9) | 0.018 (0.7) | 0.0033 (0.9) |
| MEpaleturquoise | 0.041 (0.4) | 0.0059 (0.9) | 0.017 (0.7) | −0.088 (0.09) | −0.053 (0.3) | 0.067 (0.2) | 0.075 (0.1) | 0.12 (0.02) | 0.019 (0.7) | 0.0013 (1) | 0.03 (0.6) | −0.11 (0.04) | −0.027 (0.6) | −0.0075 (0.9) | −0.0013 (1) |
| MEsteelblue | 0.023 (0.7) | 0.0044 (0.9) | 0.012 (0.8) | −0.061 (0.2) | −0.052 (0.3) | 0.08 (0.1) | 0.068 (0.2) | 0.11 (0.03) | 0.02 (0.7) | 0.015 (0.8) | 0.031 (0.5) | −0.12 (0.02) | −0.034 (0.5) | −0.0068 (0.9) | −0.036 (0.5) |
| MEturquoise | 0.073 (0.2) | 0.025 (0.6) | 0.017 (0.7) | 0.0069 (0.9) | −0.026 (0.6) | 0.046 (0.4) | 0.027 (0.6) | 0.051 (0.3) | −0.002 (1) | −0.0042 (0.9) | 0.0034 (0.9) | −0.043 (0.4) | 0.02 (0.7) | −0.0035 (0.9) | −0.0018 (1) |
| MEcyan | 0.035 (0.5) | 0.0065 (0.9) | 0.029 (0.6) | −0.048 (0.3) | −0.026 (0.6) | 0.0031 (1) | 0.041 (0.4) | 0.05 (0.3) | −0.015 (0.8) | −0.017 (0.7) | −0.012 (0.8) | −0.015 (0.8) | −0.017 (0.7) | 0.0073 (0.9) | 0.011 (0.8) |
| MEmagenta | 0.065 (0.2) | 0.016 (0.8) | 0.021 (0.7) | 0.0074 (0.9) | −0.023 (0.7) | 0.015 (0.8) | 0.025 (0.6) | 0.012 (0.8) | −0.02 (0.7) | −0.0089 (0.9) | −0.0094 (0.9) | −0.0021 (1) | 0.012 (0.8) | 0.01 (0.8) | −0.0061 (0.9) |
| MEdarkturquoise | 0.026 (0.6) | 0.0046 (0.9) | 0.022 (0.7) | −0.05 (0.3) | −0.038 (0.5) | 0.035 (0.5) | 0.066 (0.2) | 0.079 (0.1) | −0.011 (0.8) | −0.0083 (0.9) | 0.0036 (0.9) | −0.054 (0.3) | −0.015 (0.8) | −0.00042 (1) | −0.0039 (0.9) |
| MEroyalblue | 0.012 (0.8) | −0.0029 (1) | 0.03 (0.6) | −0.085 (0.1) | −0.035 (0.5) | 0.011 (0.8) | 0.072 (0.2) | 0.082 (0.1) | −0.0023 (1) | −0.014 (0.8) | 0.0049 (0.9) | −0.041 (0.4) | −0.028 (0.6) | −0.00088 (1) | 0.021 (0.7) |
| MEgrey | 0.05 (0.3) | 0.011 (0.8) | 0.0066 (0.9) | −0.05 (0.3) | −0.046 (0.4) | 0.061 (0.2) | 0.053 (0.3) | 0.096 (0.06) | 0.02 (0.7) | 0.0055 (0.9) | 0.033 (0.5) | −0.091 (0.08) | −0.024 (0.6) | −0.0083 (0.9) | −0.0065 (0.9) |

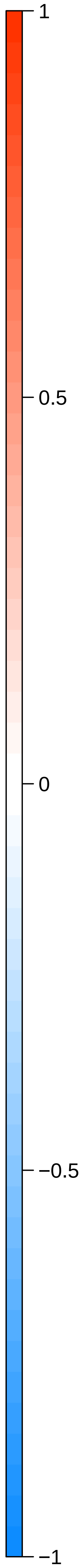
