## Supplementary figures and images for "Transcriptional and genetic sex differences for schizophrenia across the dorsolateral prefrontal cortex, hippocampus, and caudate nucleus"

### cluster_dendrogram.pdf

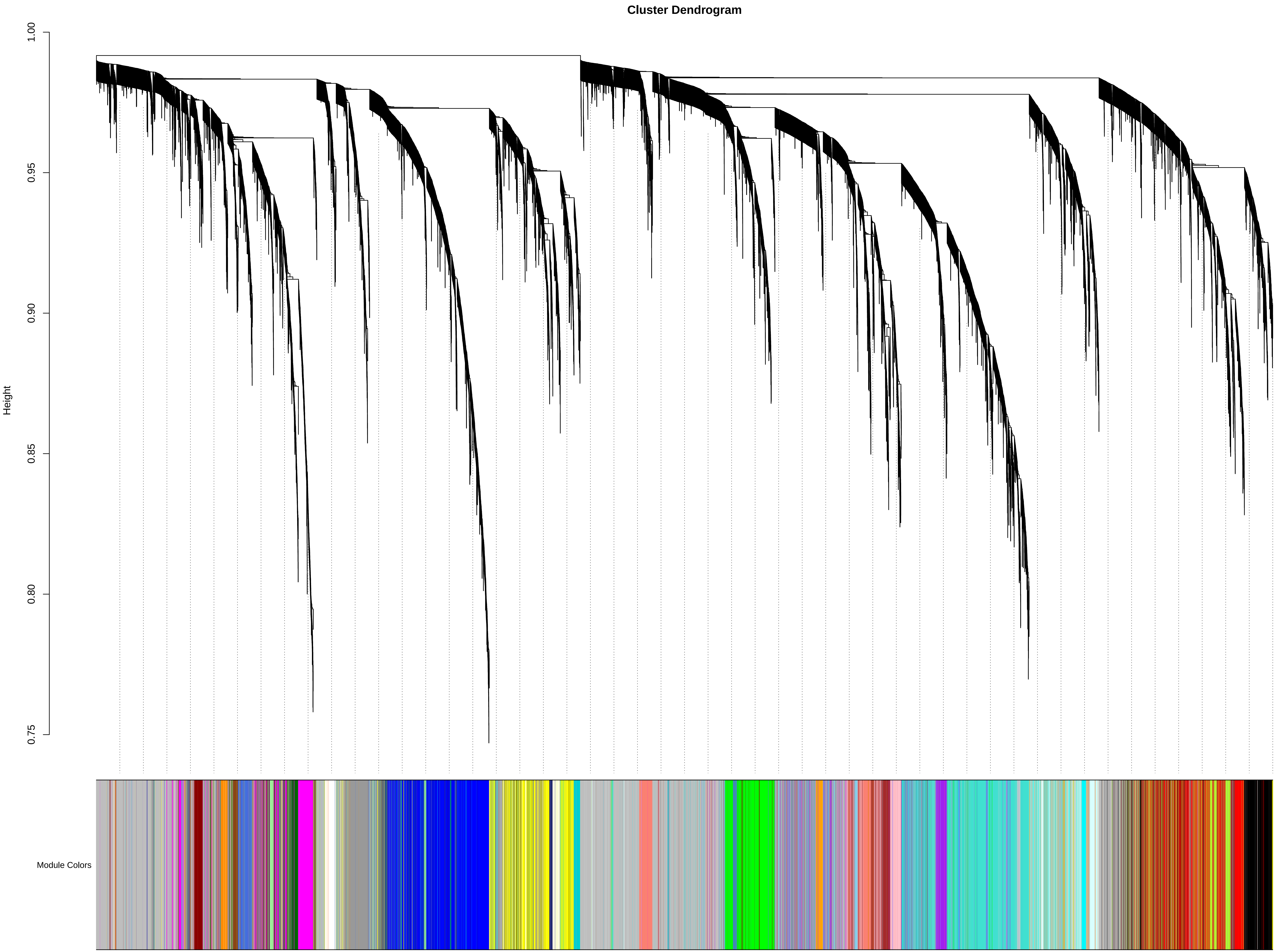

### cluster_dendrogram.pdf

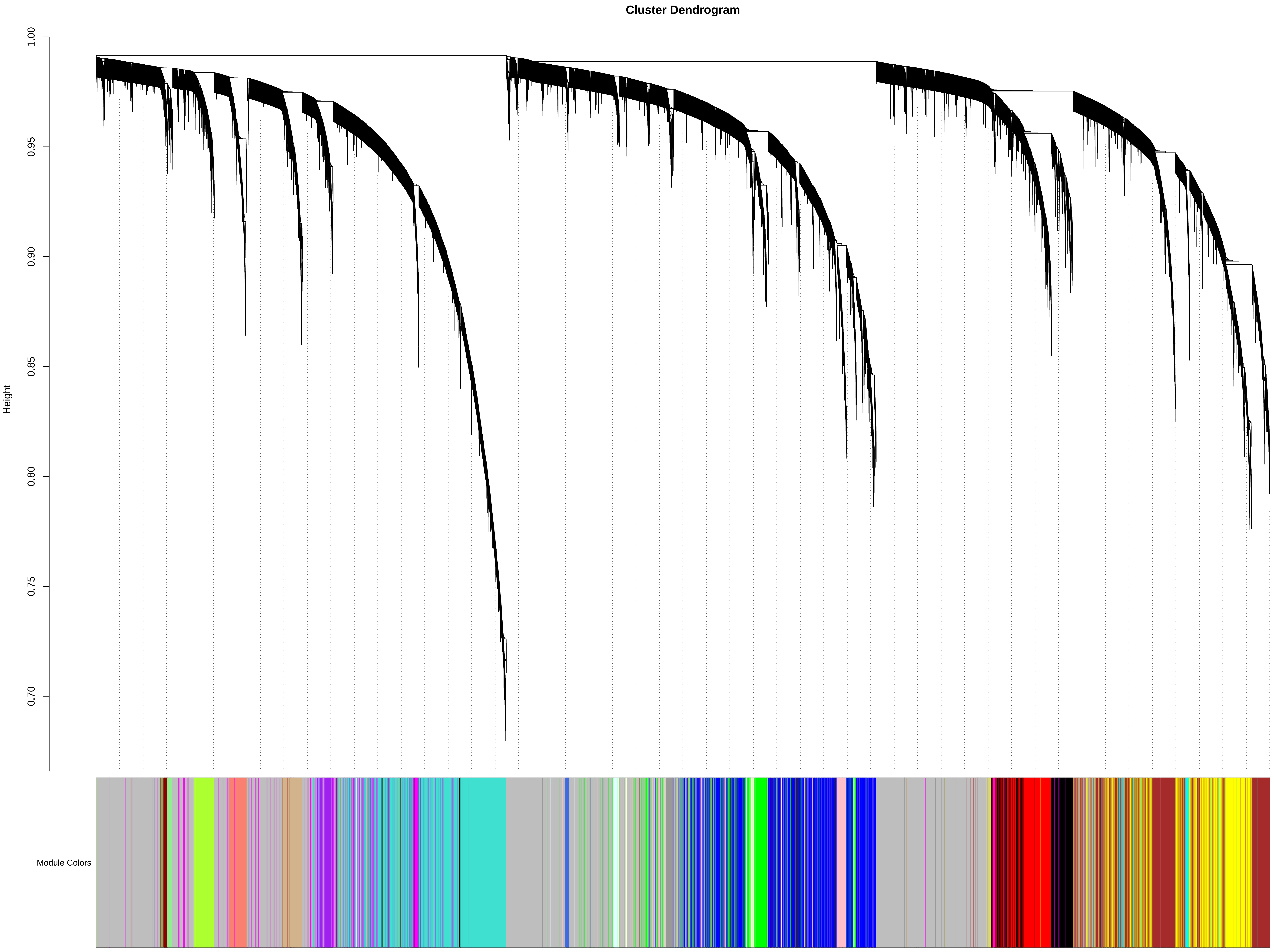

### cluster_dendrogram.pdf

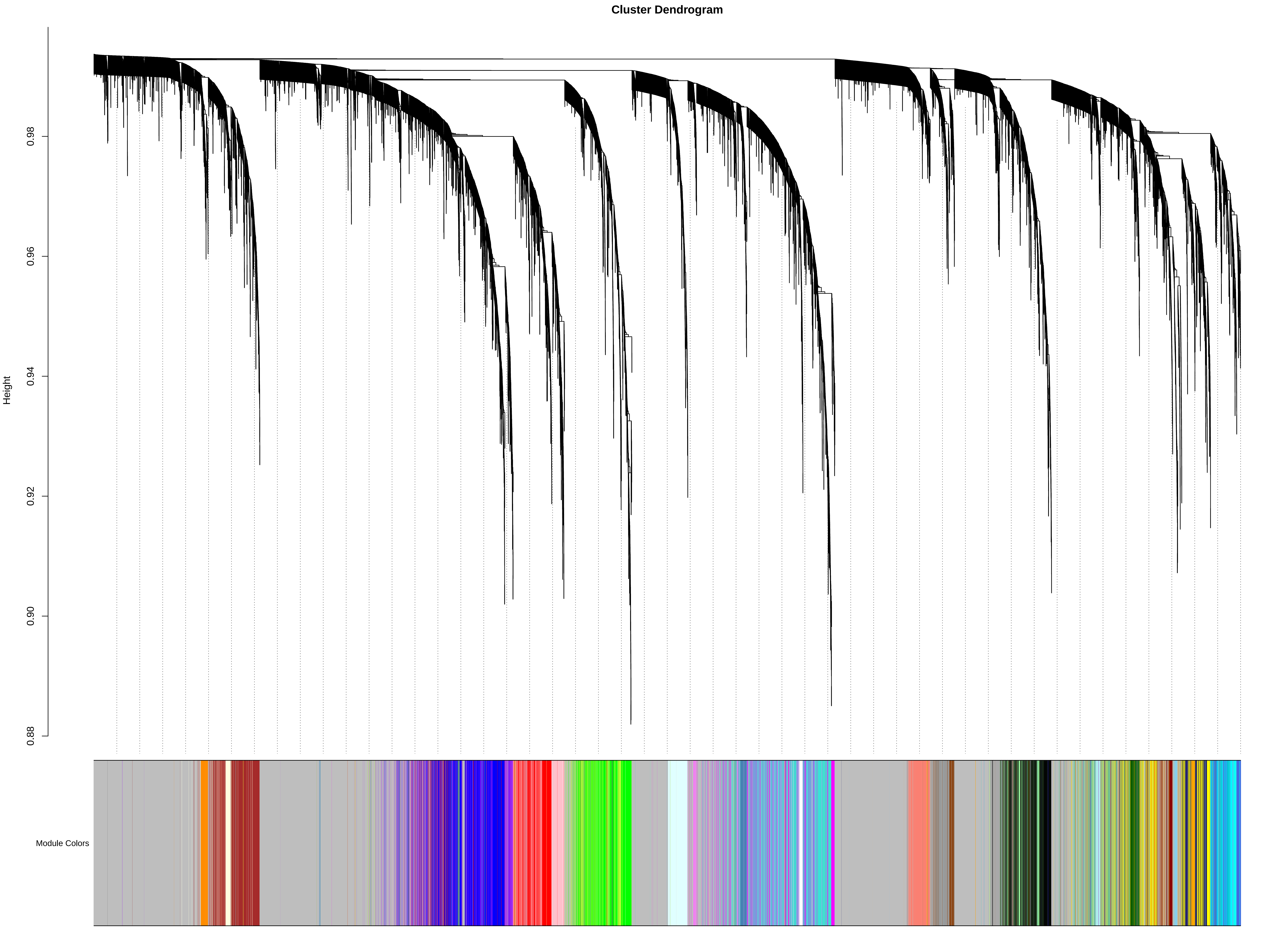

### ENSG00000006744.18.SCZD.dlpfc.WithoutCongruenceData.WithLinkageData.eQTpLot.png

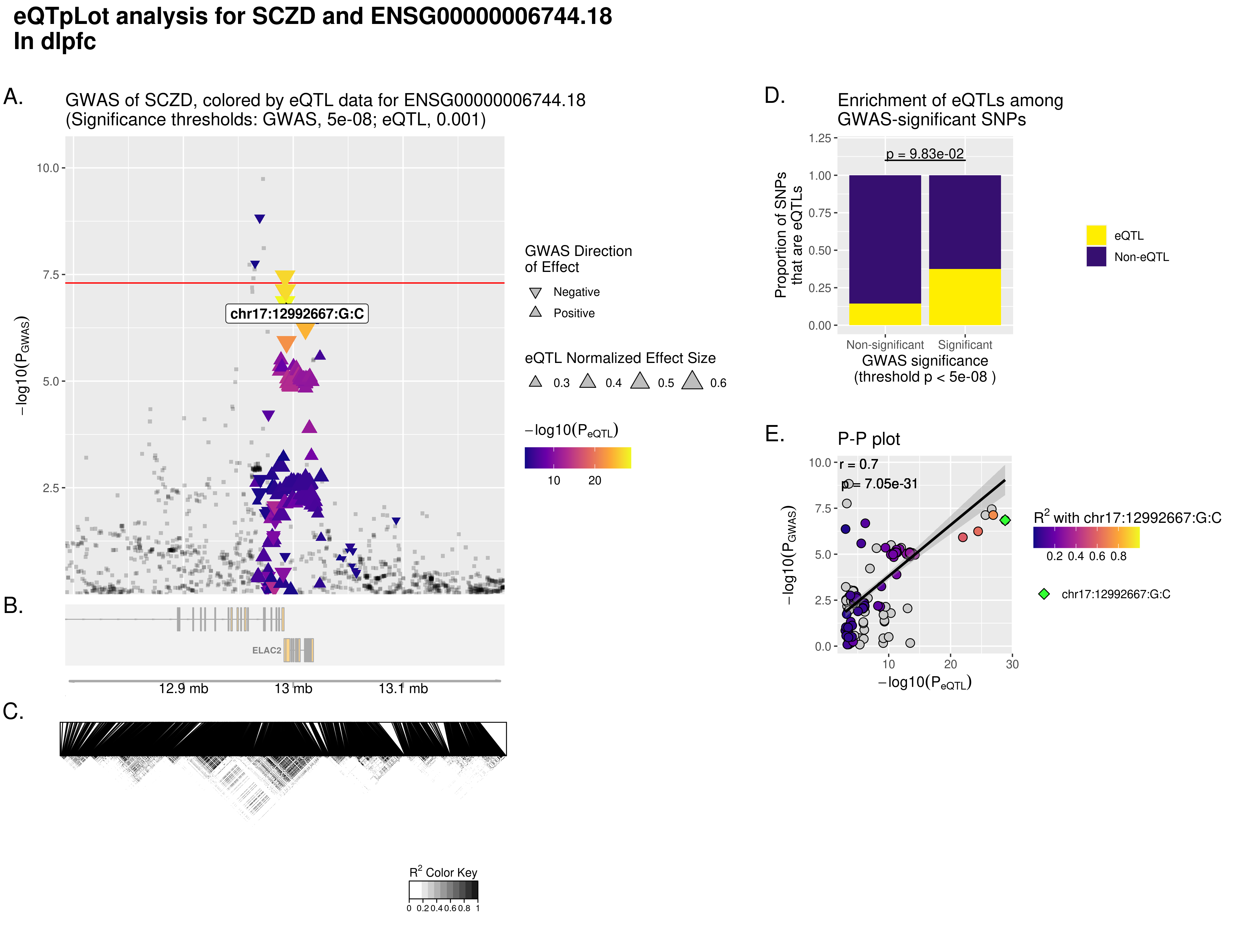

### ENSG00000006744.18.SCZD.dlpfc.WithoutCongruenceData.WithLinkageData.eQTpLot.png

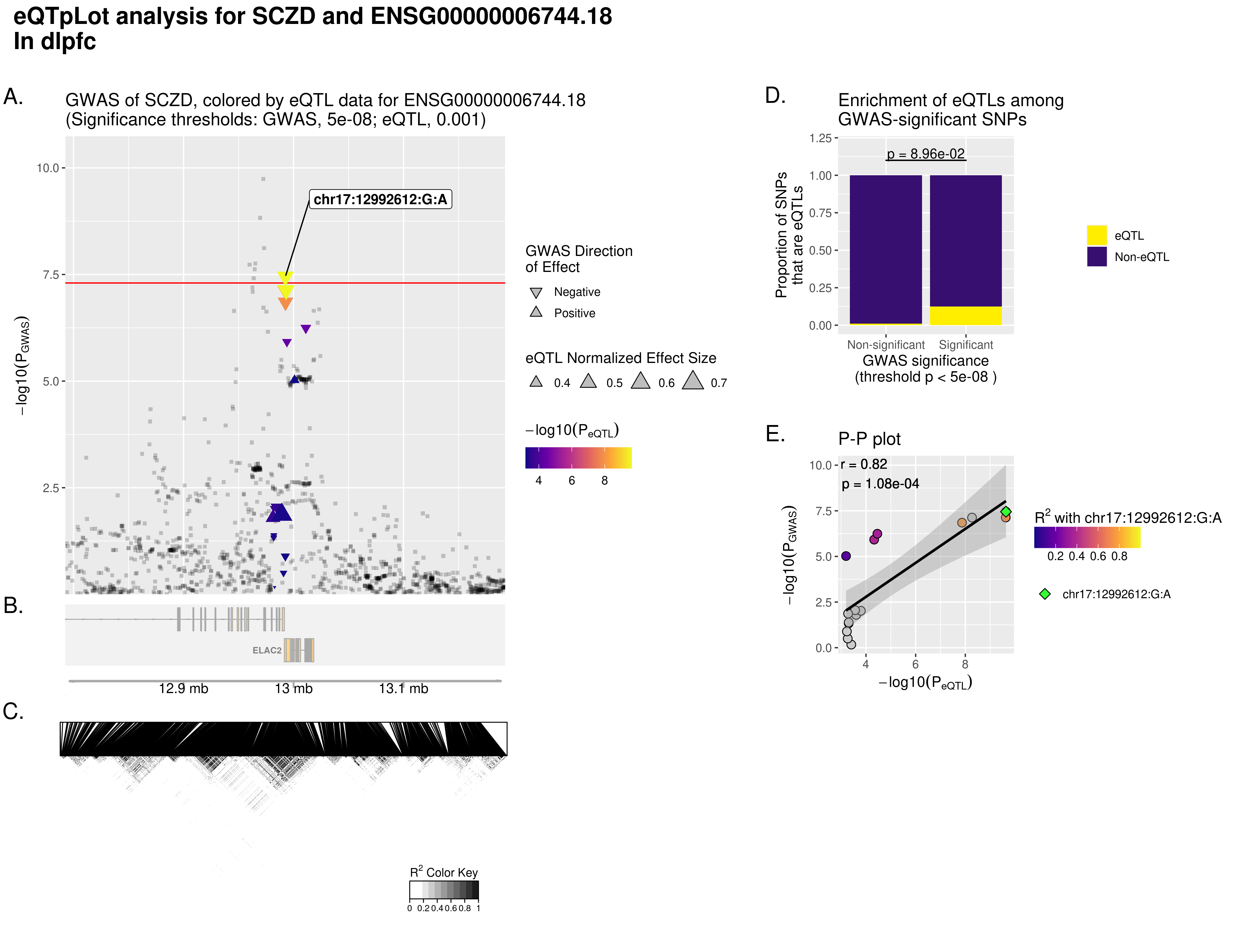

### ENSG00000108960.7.SCZD.dlpfc.WithoutCongruenceData.WithLinkageData.eQTpLot.png

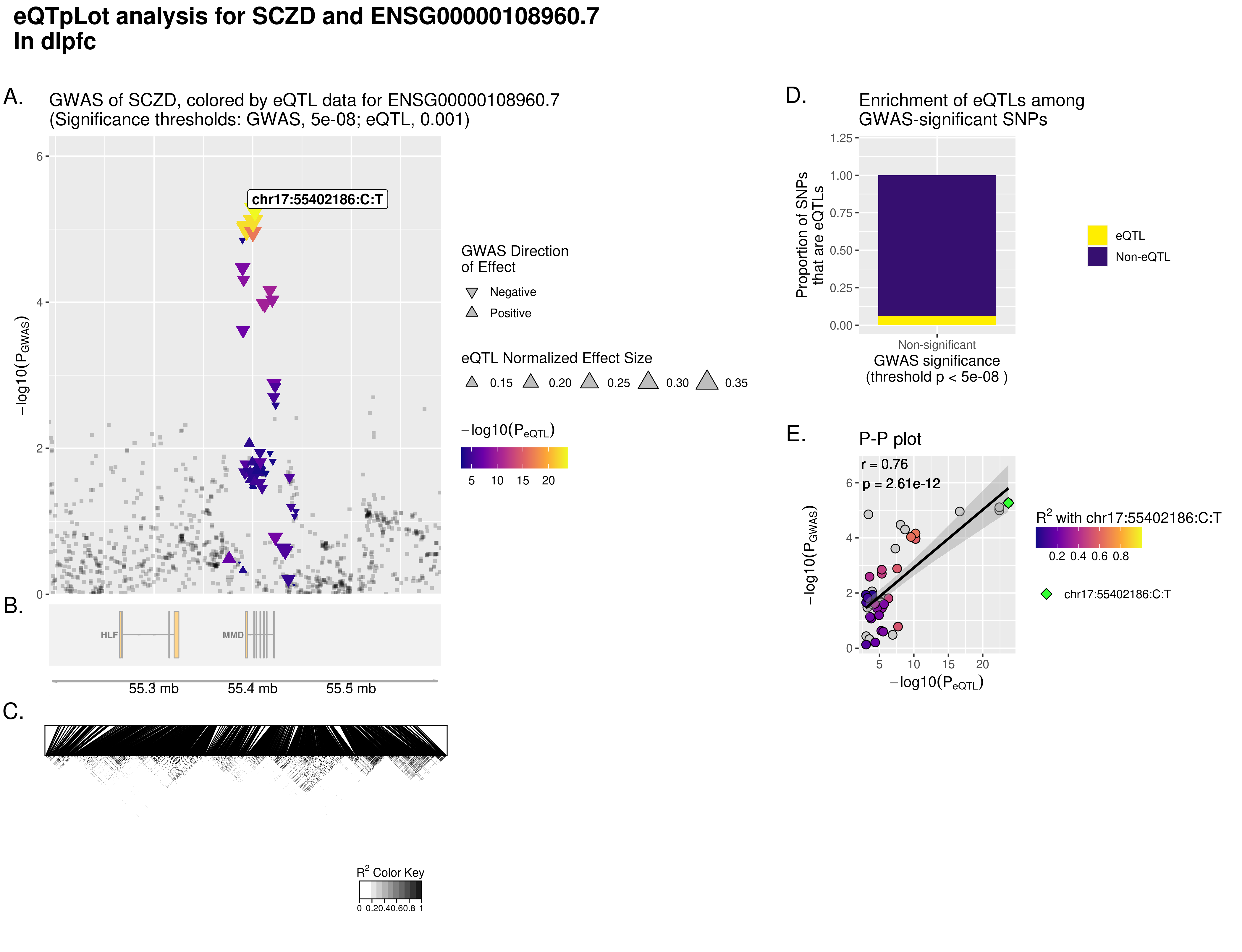

### ENSG00000108960.7.SCZD.dlpfc.WithoutCongruenceData.WithLinkageData.eQTpLot.png

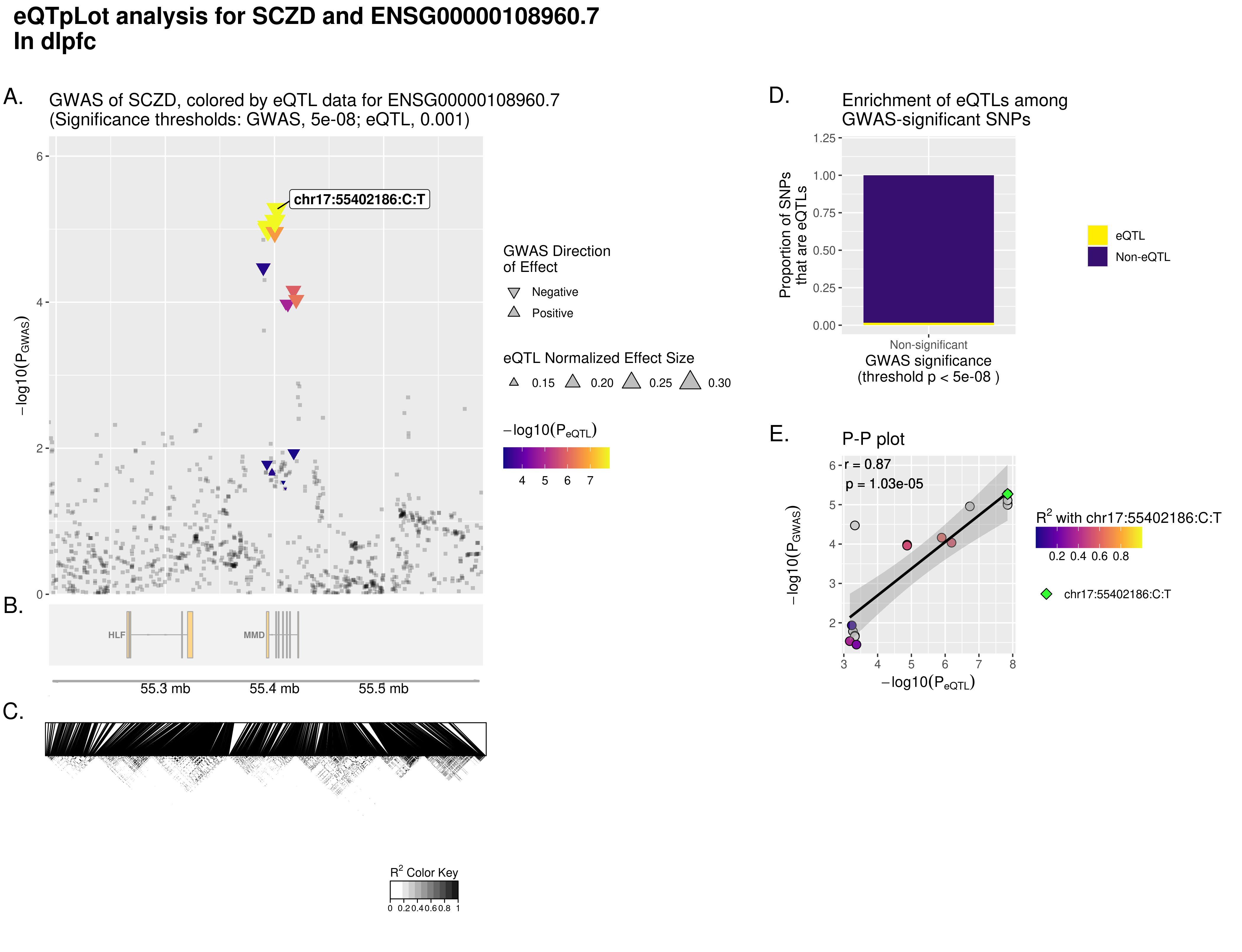

### ENSG00000108960.7.SCZD.hippocampus.WithoutCongruenceData.WithLinkageData.eQTpLot.png

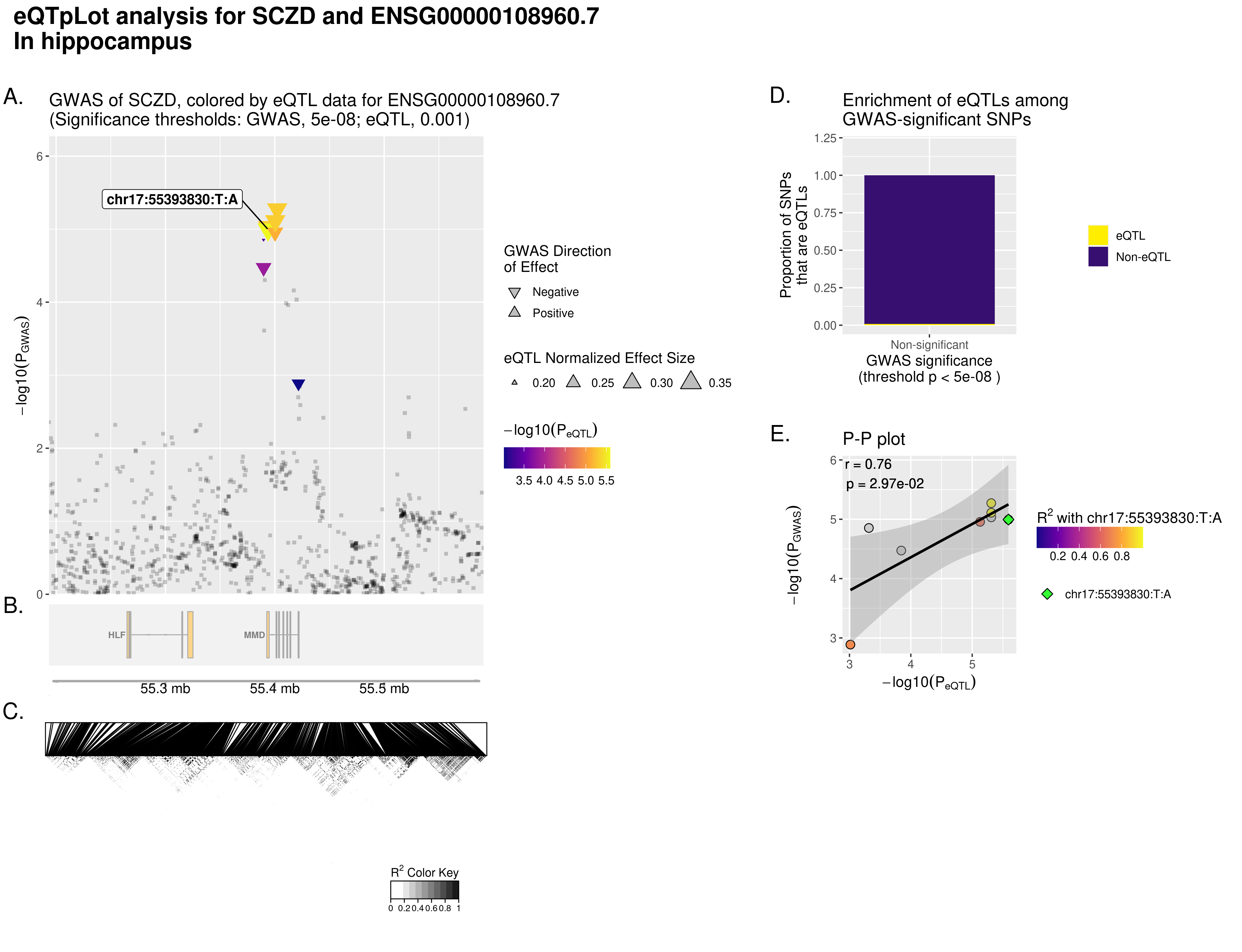

### ENSG00000108960.7.SCZD.hippocampus.WithoutCongruenceData.WithLinkageData.eQTpLot.png

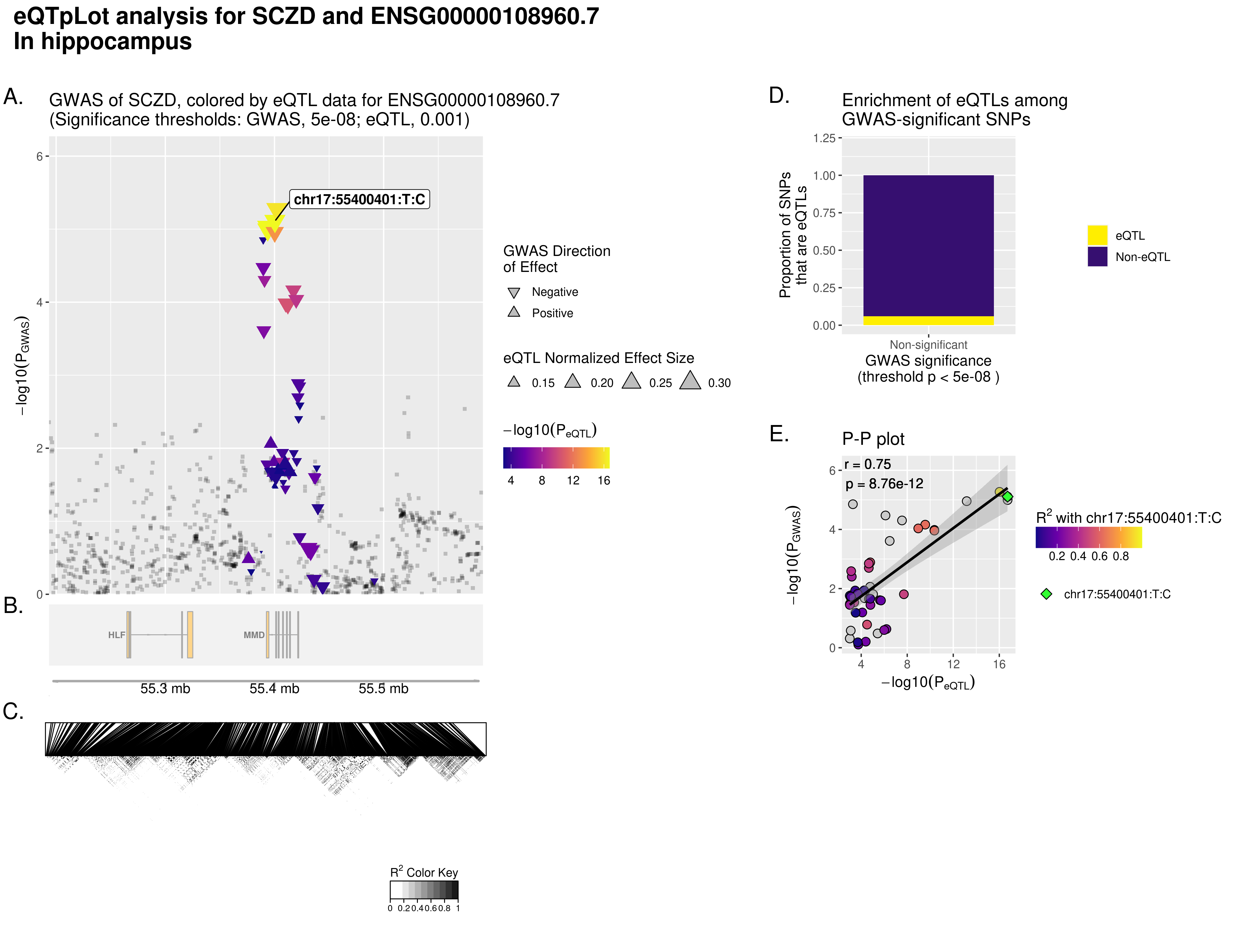

### ENSG00000136295.14.SCZD.dlpfc.WithoutCongruenceData.WithLinkageData.eQTpLot.png

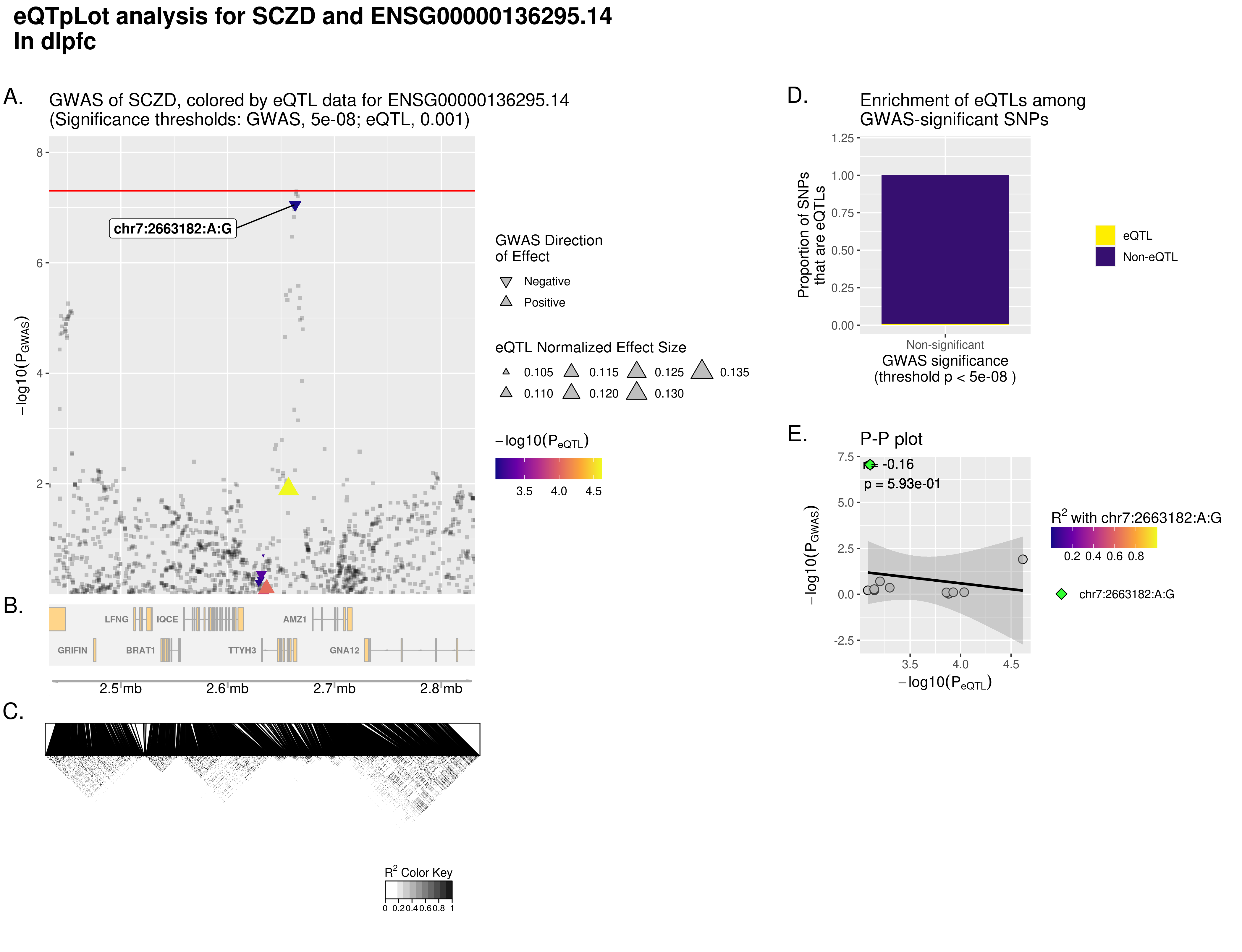

### ENSG00000140265.12.SCZD.dlpfc.WithoutCongruenceData.WithLinkageData.eQTpLot.png

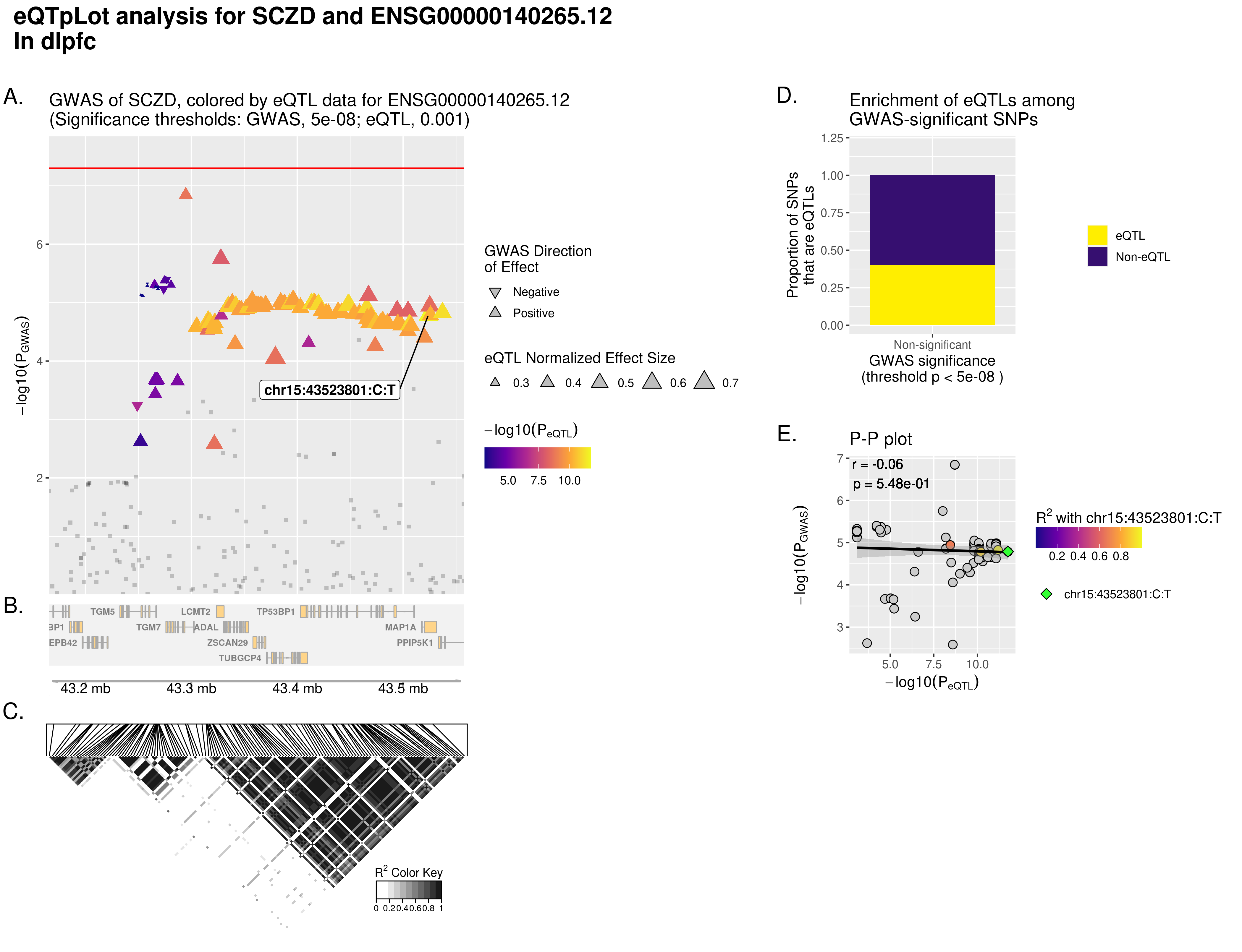

### ENSG00000140265.12.SCZD.dlpfc.WithoutCongruenceData.WithLinkageData.eQTpLot.png

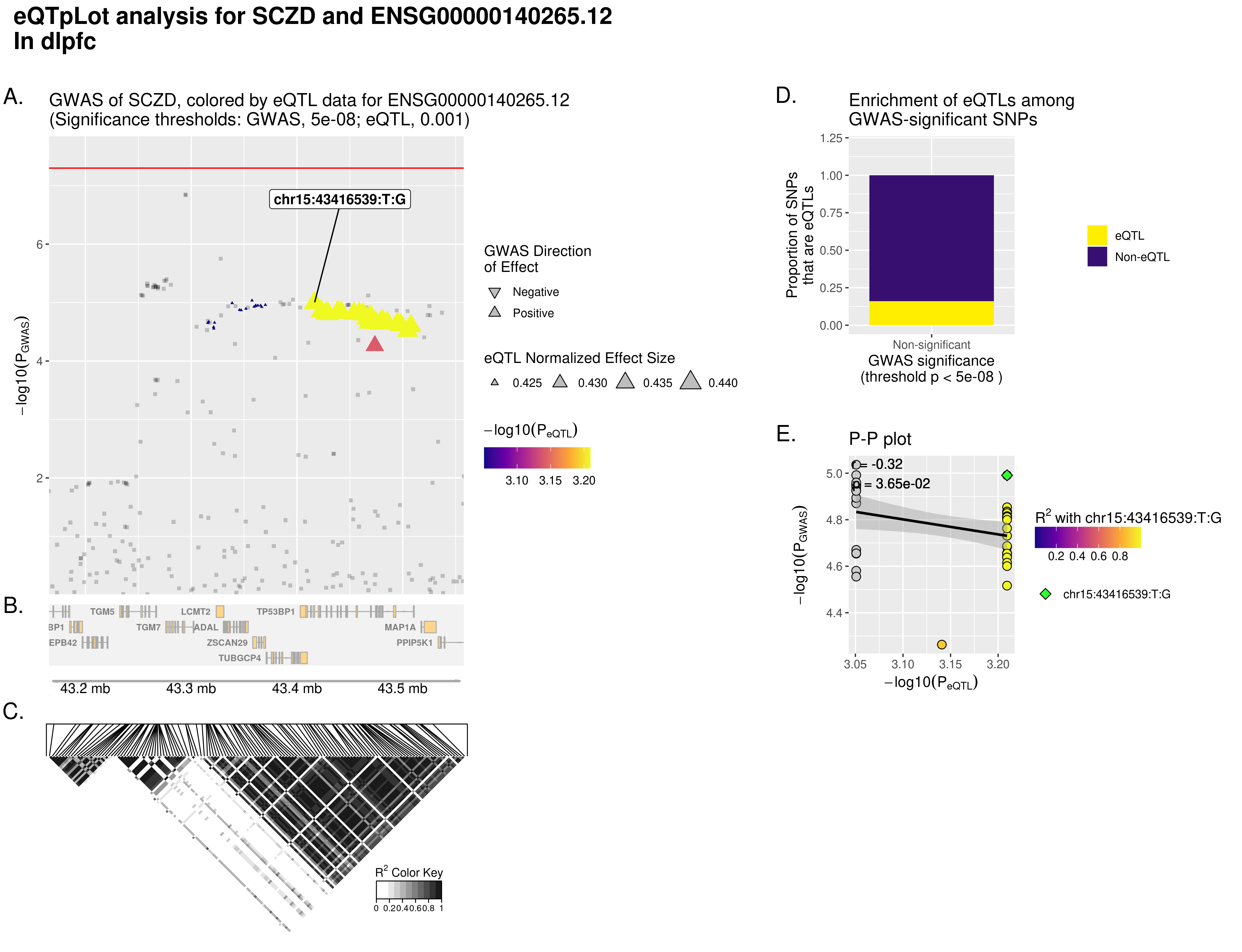

### ENSG00000140564.10.SCZD.dlpfc.WithoutCongruenceData.WithLinkageData.eQTpLot.png

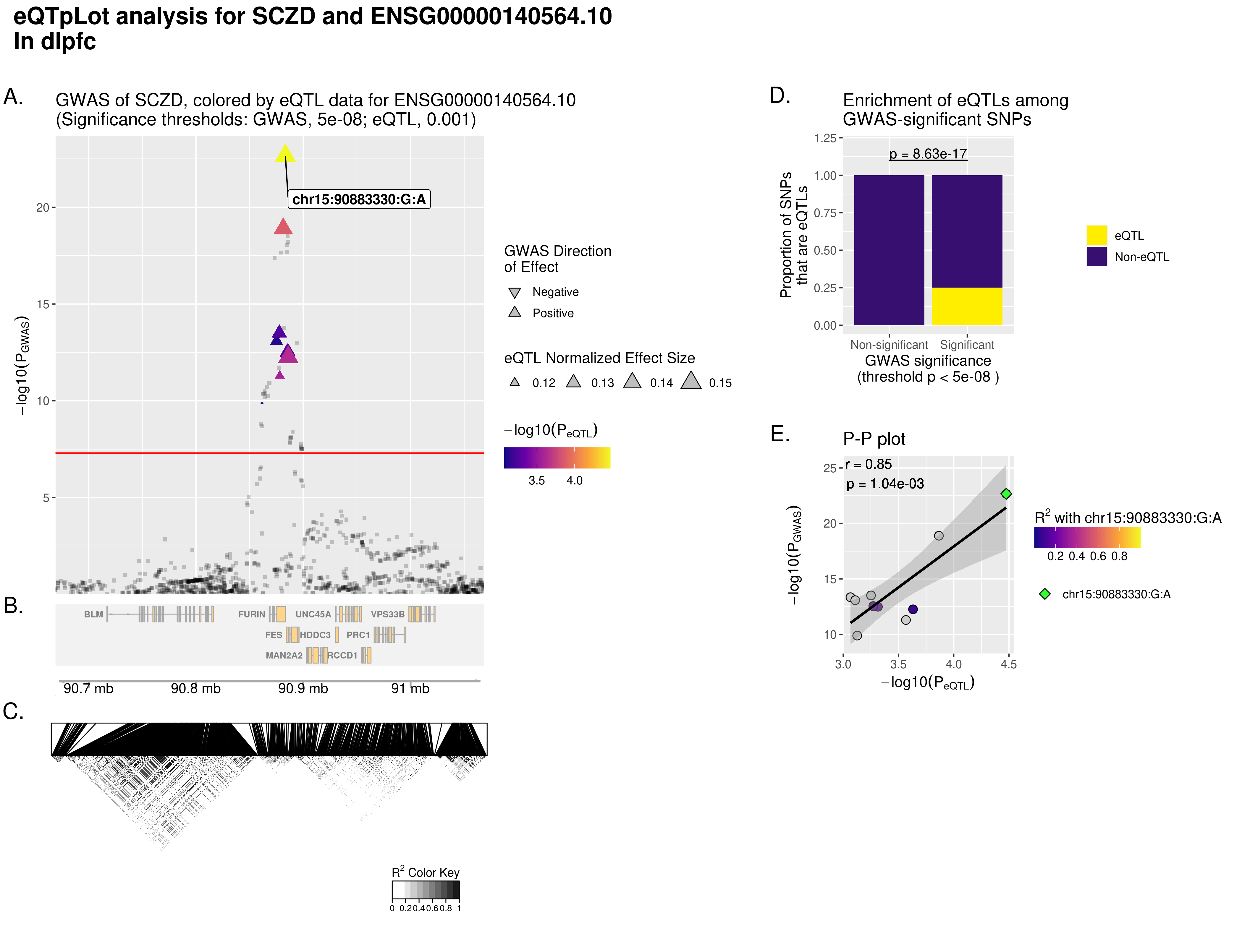

### ENSG00000140564.10.SCZD.hippocampus.WithoutCongruenceData.WithLinkageData.eQTpLot.png

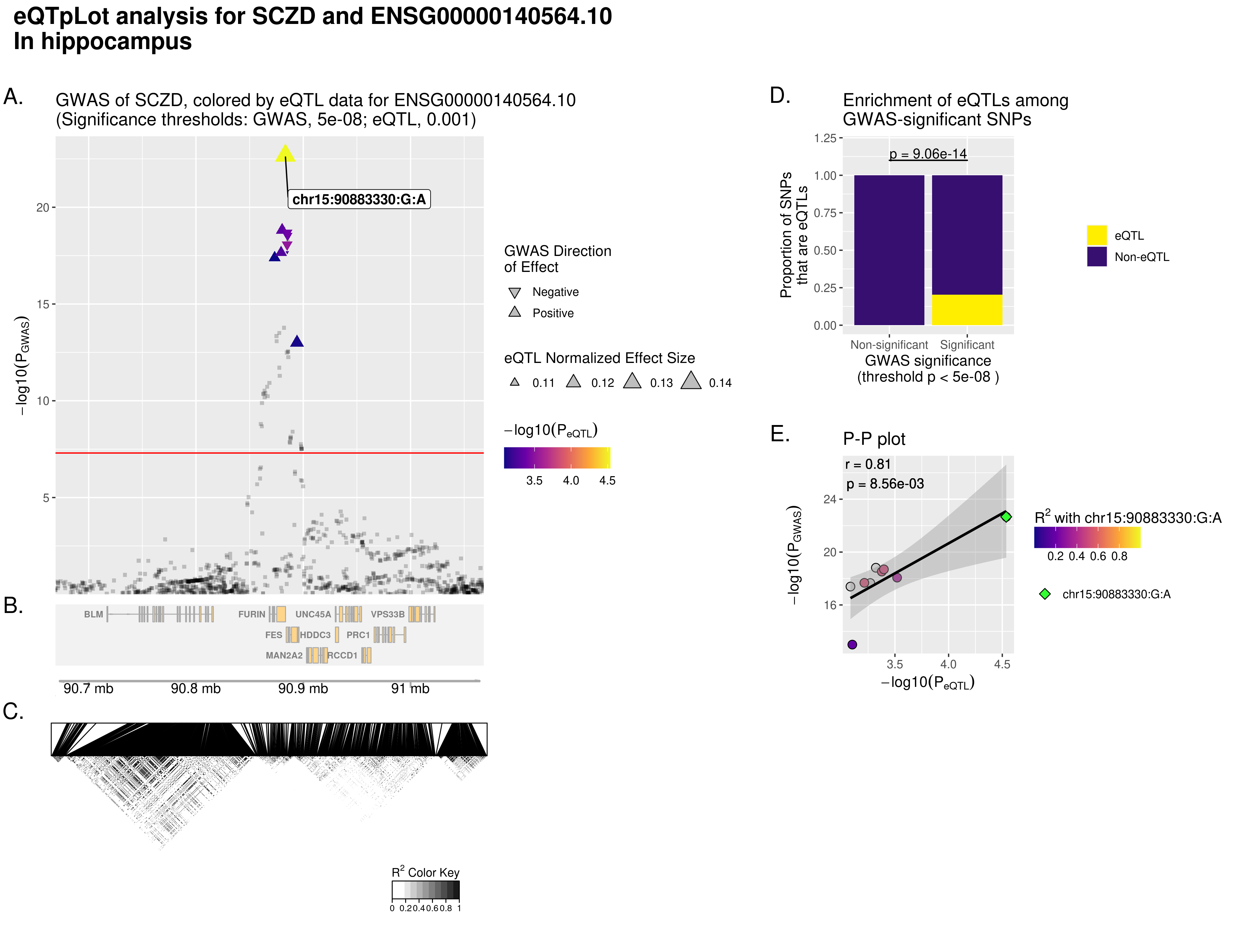

### ENSG00000159640.15.SCZD.dlpfc.WithoutCongruenceData.WithLinkageData.eQTpLot.png

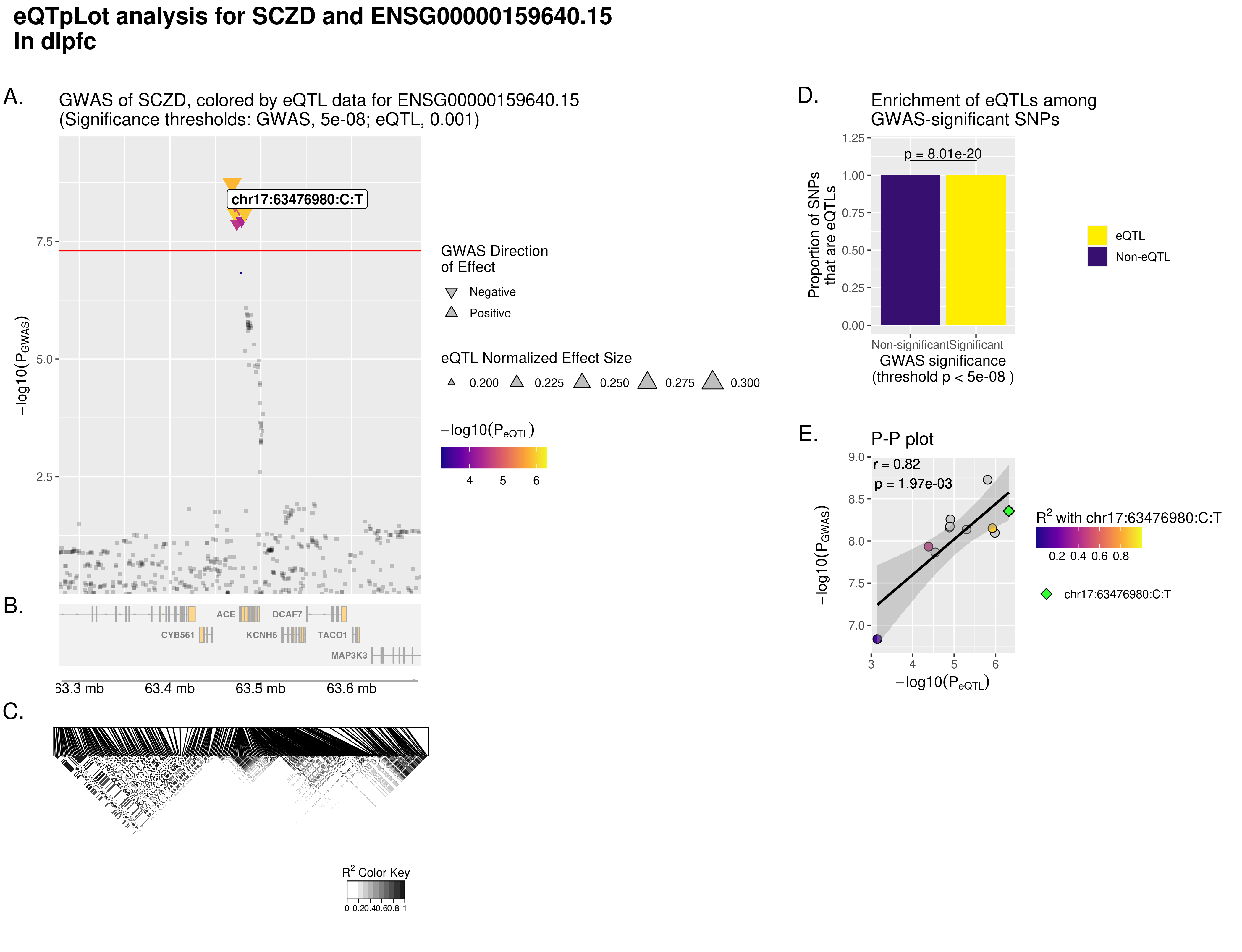

### ENSG00000159640.15.SCZD.dlpfc.WithoutCongruenceData.WithLinkageData.eQTpLot.png

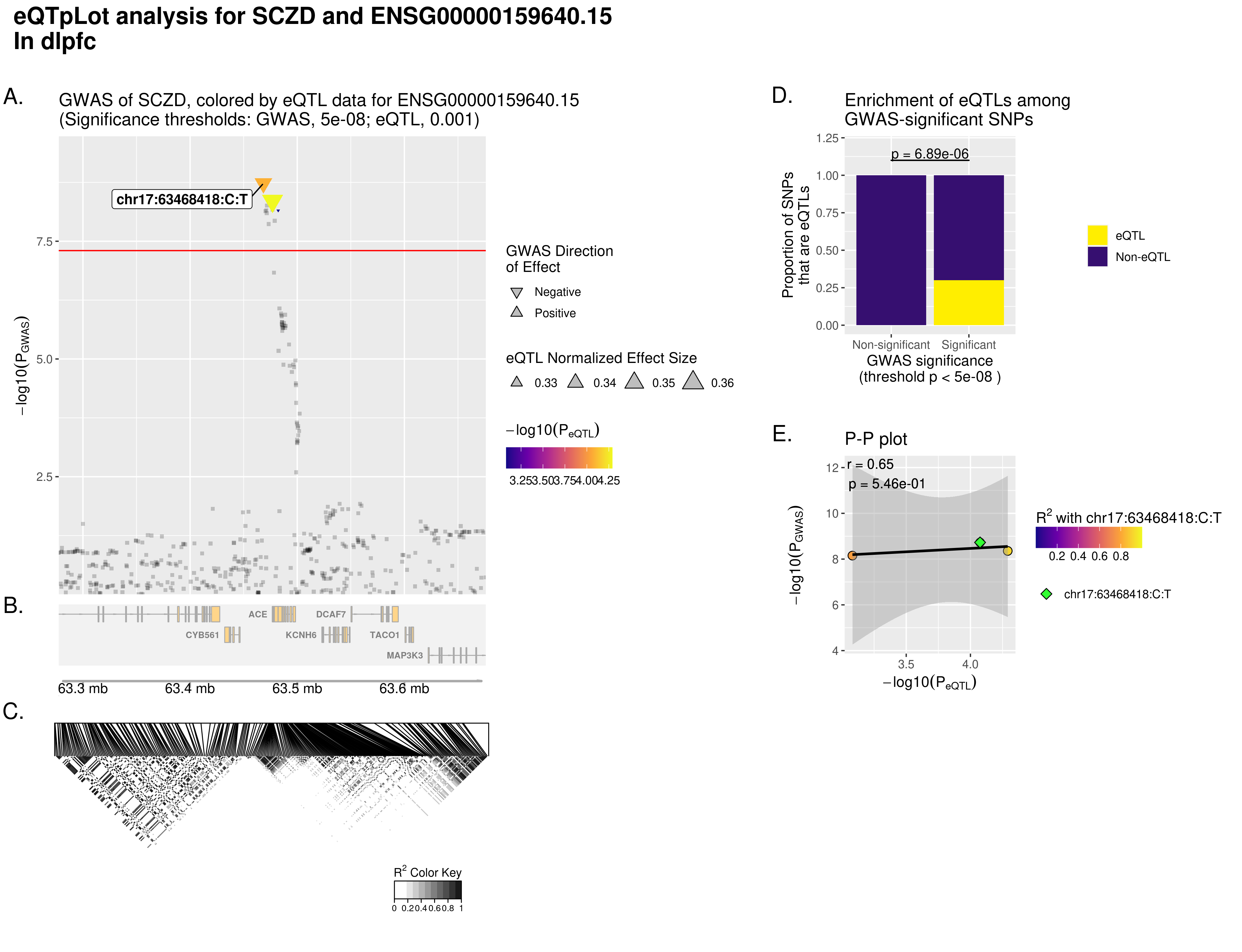

### ENSG00000170802.15.SCZD.dlpfc.WithoutCongruenceData.WithLinkageData.eQTpLot.png

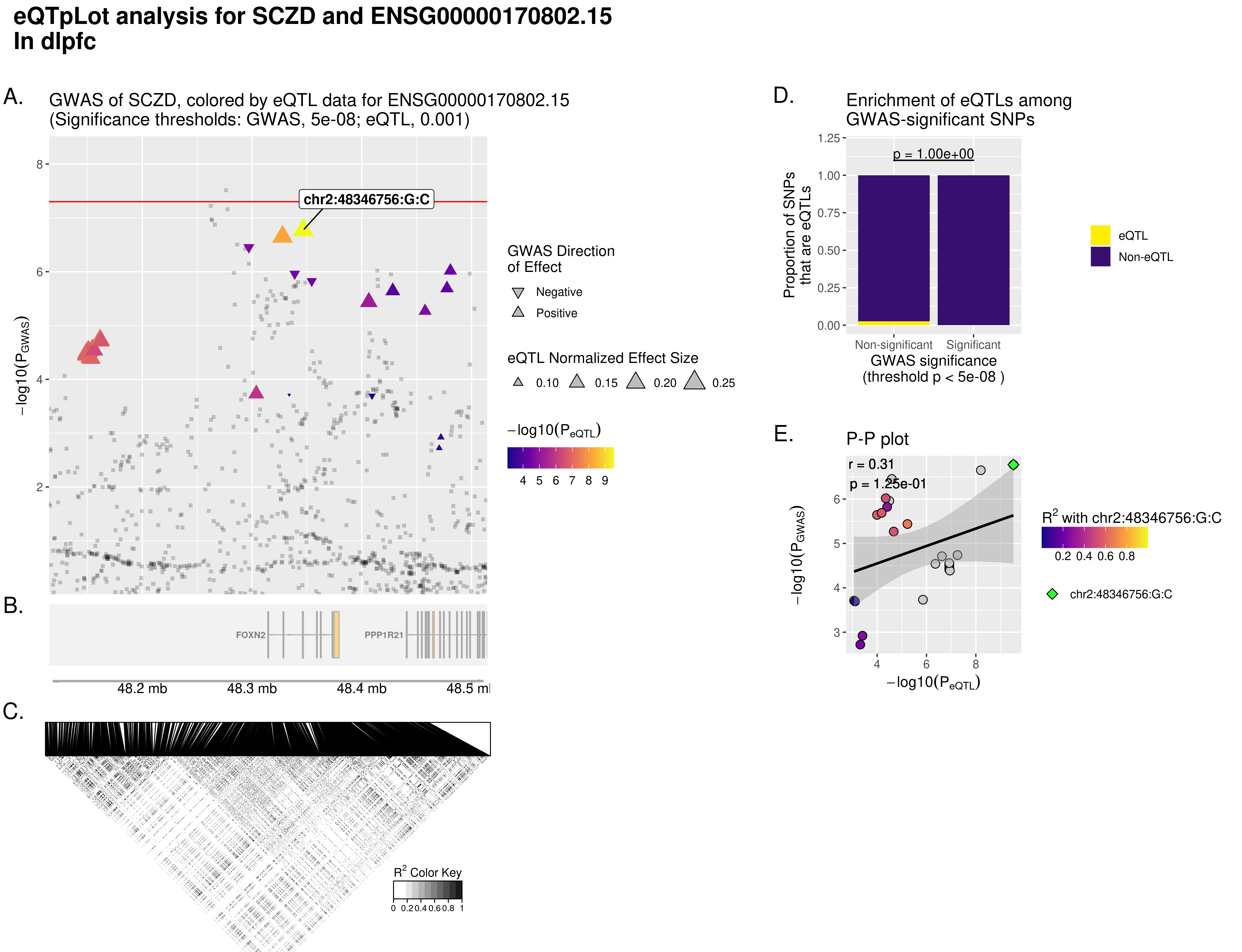

### ENSG00000203499.11.SCZD.dlpfc.WithoutCongruenceData.WithLinkageData.eQTpLot.png

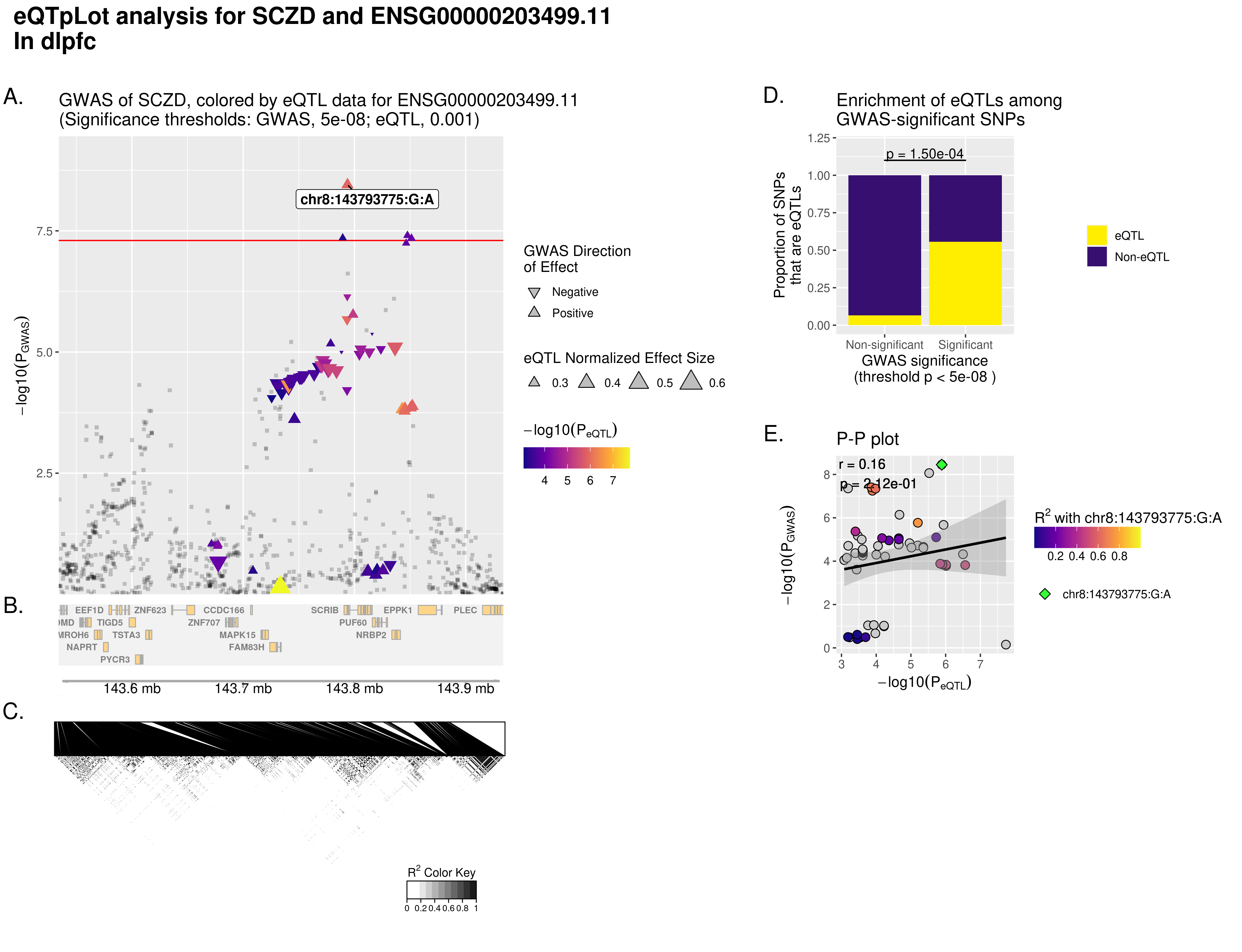

### ENSG00000224924.6.SCZD.dlpfc.WithoutCongruenceData.WithLinkageData.eQTpLot.png

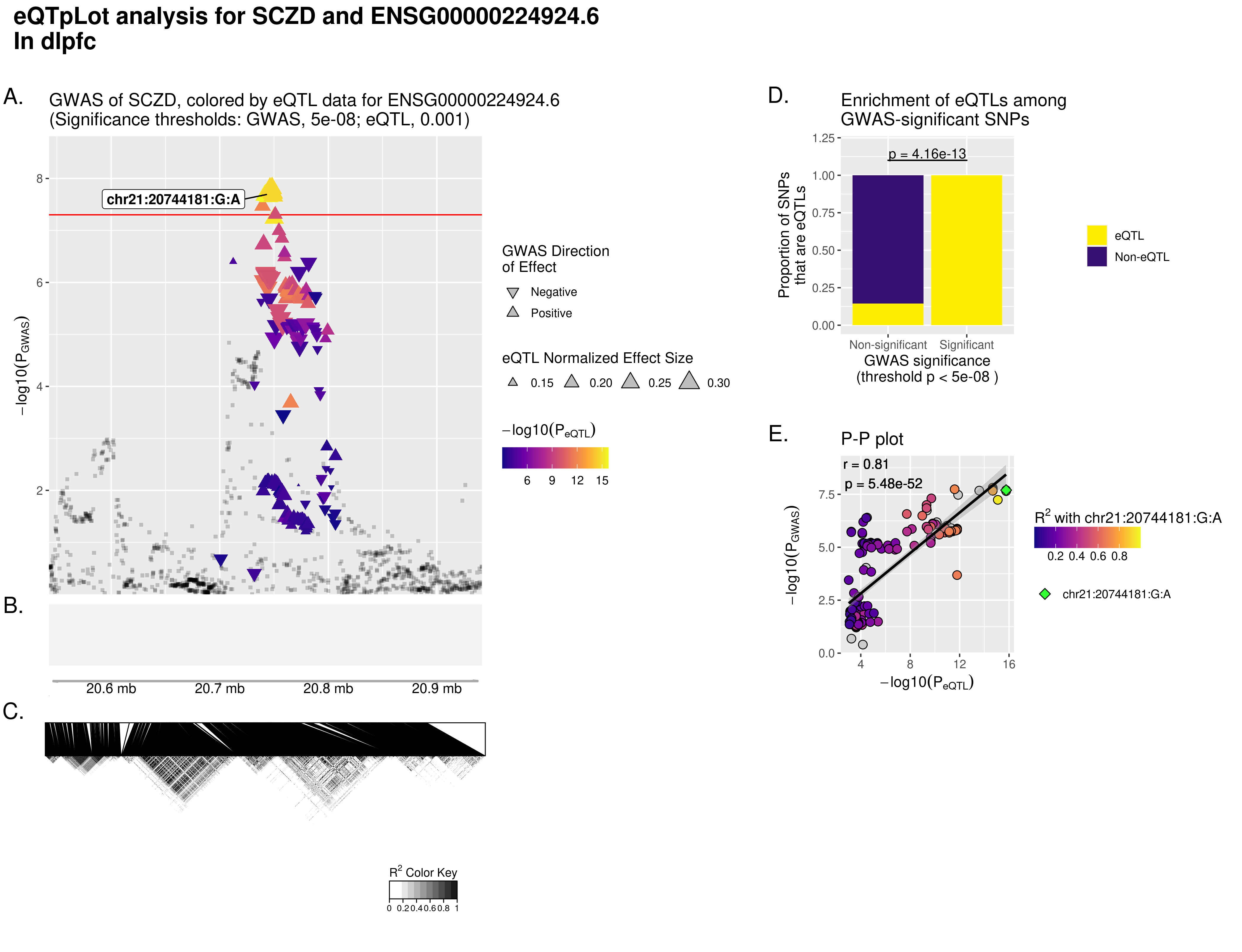

### ENSG00000224924.6.SCZD.dlpfc.WithoutCongruenceData.WithLinkageData.eQTpLot.png

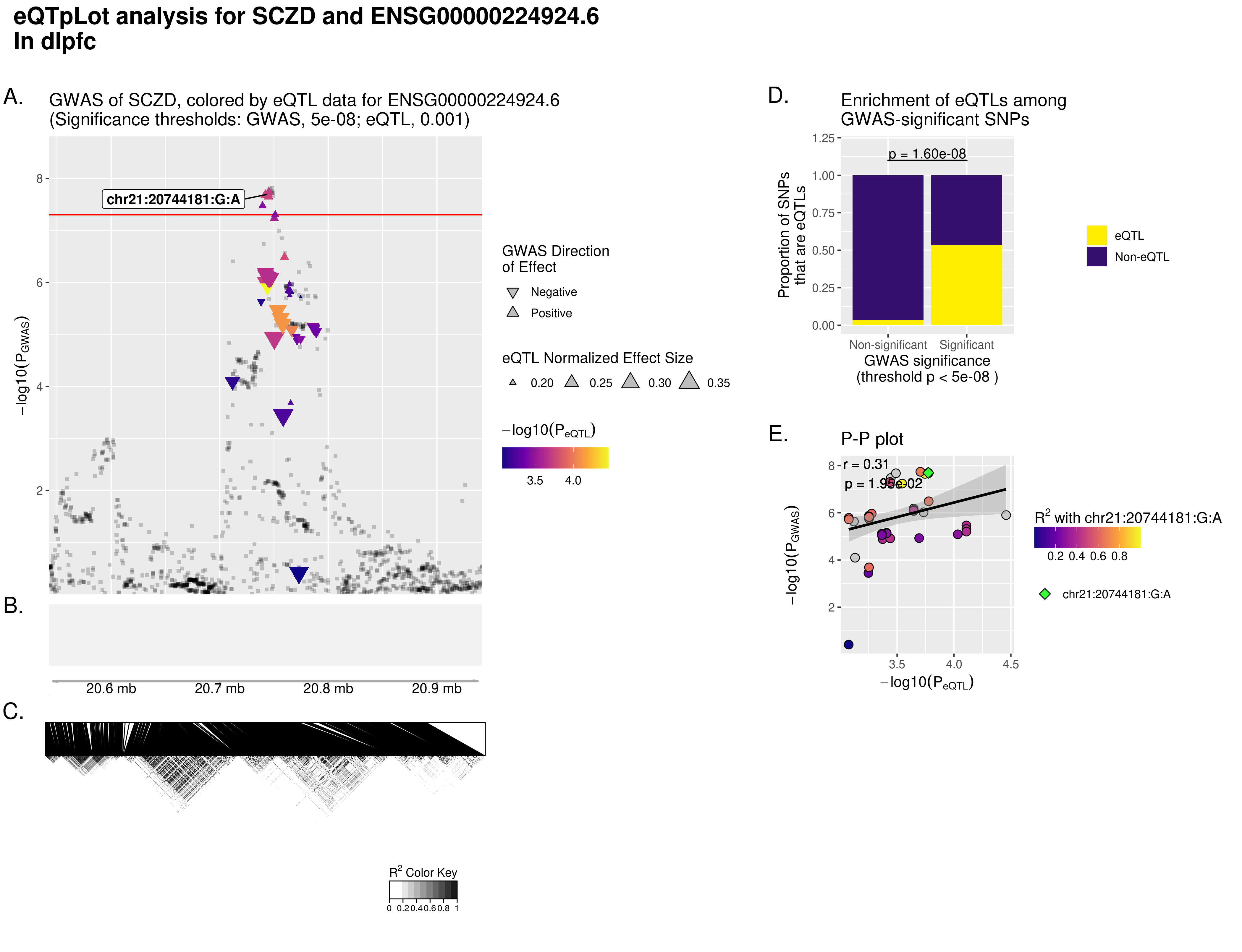

### ENSG00000226124.6.SCZD.dlpfc.WithoutCongruenceData.WithLinkageData.eQTpLot.png

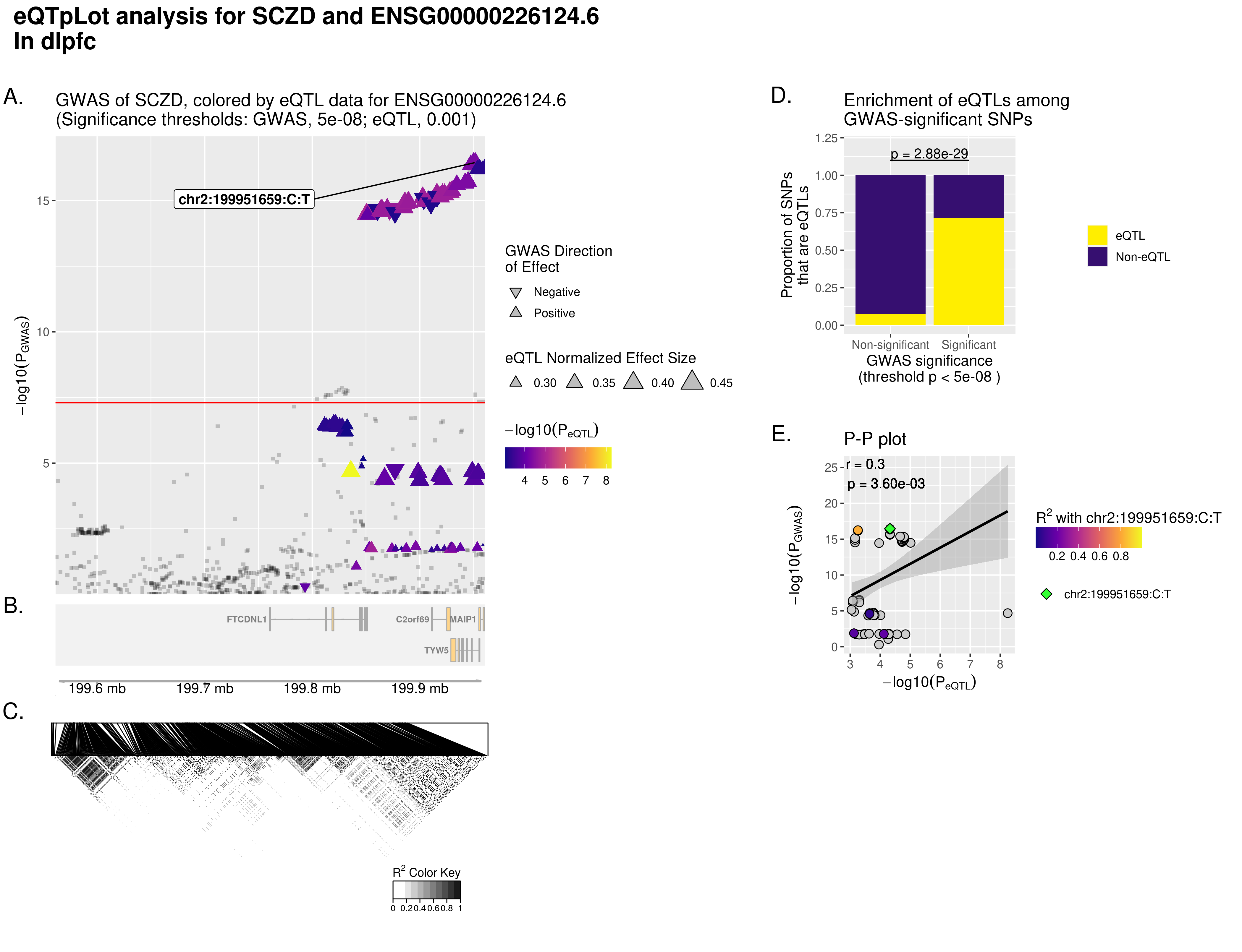

### ENSG00000226124.6.SCZD.dlpfc.WithoutCongruenceData.WithLinkageData.eQTpLot.png

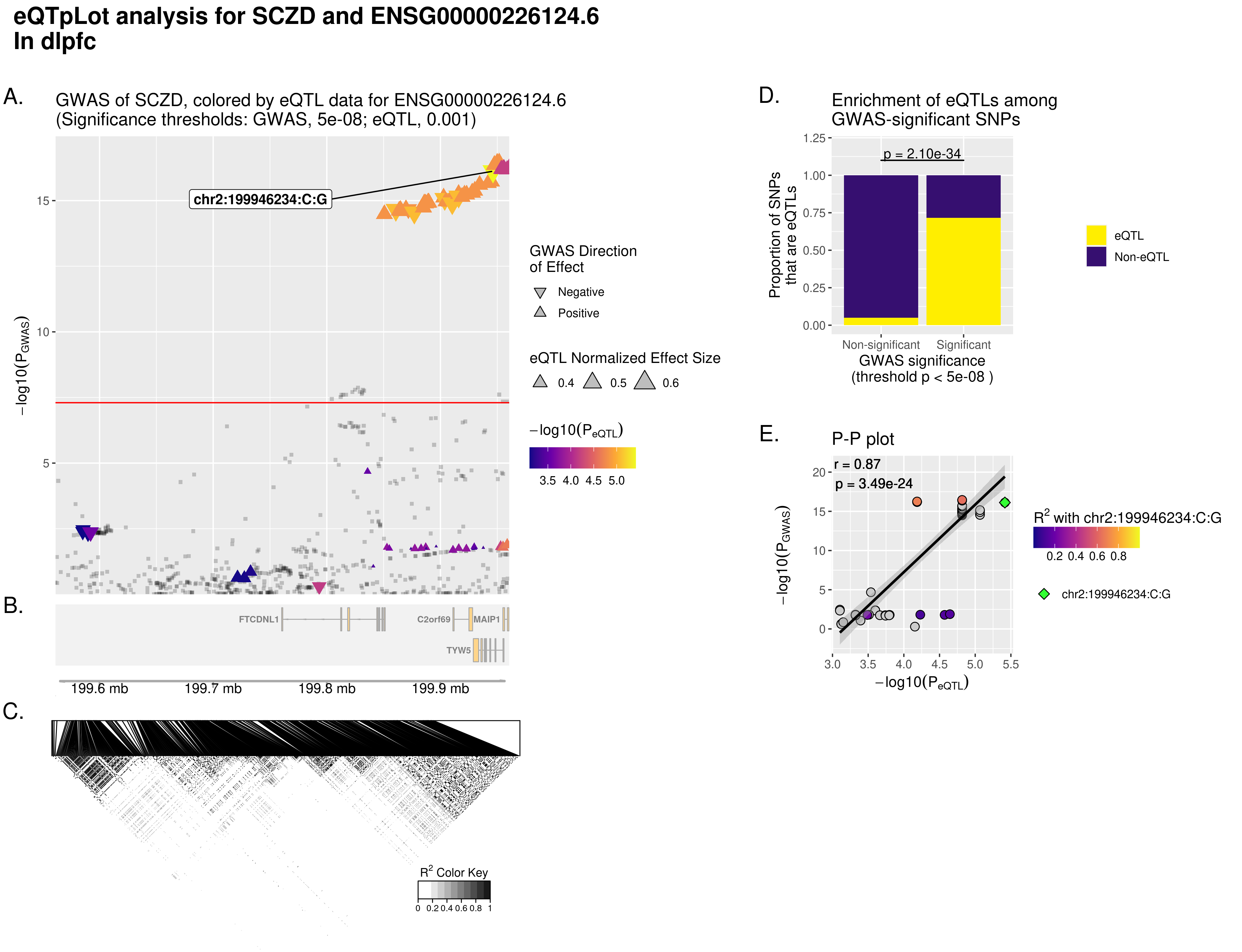

### ENSG00000249839.1.SCZD.dlpfc.WithoutCongruenceData.WithLinkageData.eQTpLot.png

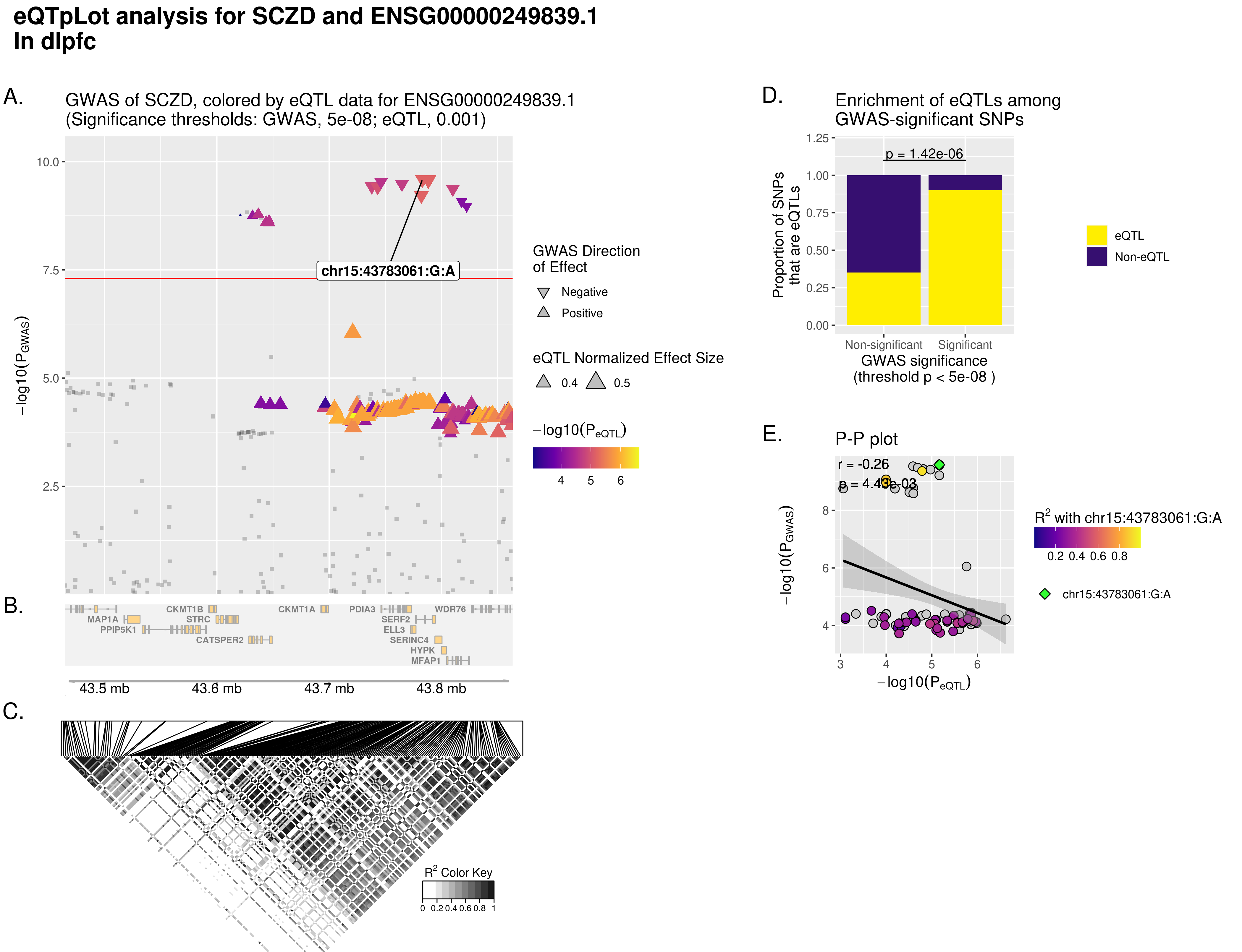

### ENSG00000254427.1.SCZD.dlpfc.WithoutCongruenceData.WithLinkageData.eQTpLot.png

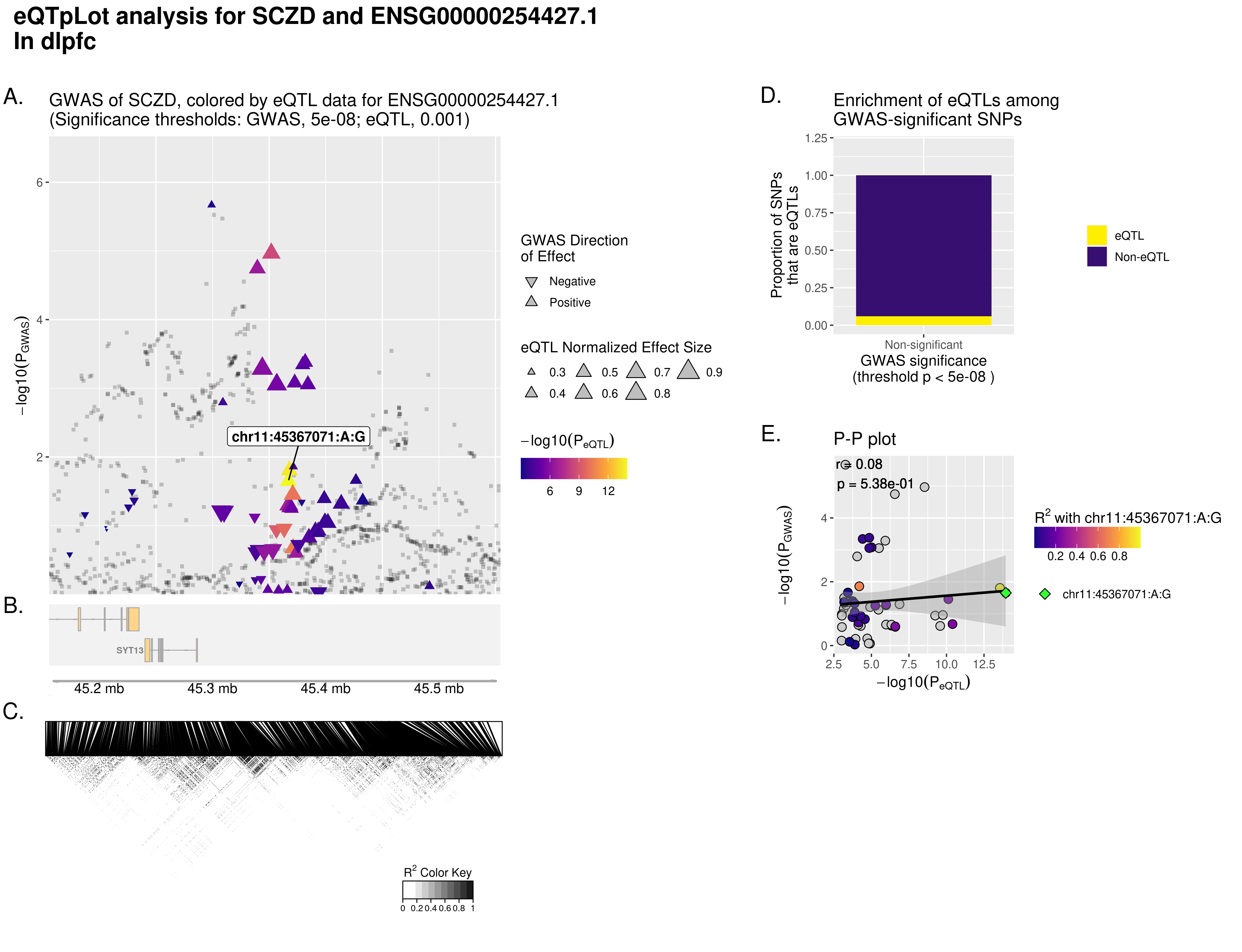

### ENSG00000254427.1.SCZD.dlpfc.WithoutCongruenceData.WithLinkageData.eQTpLot.png

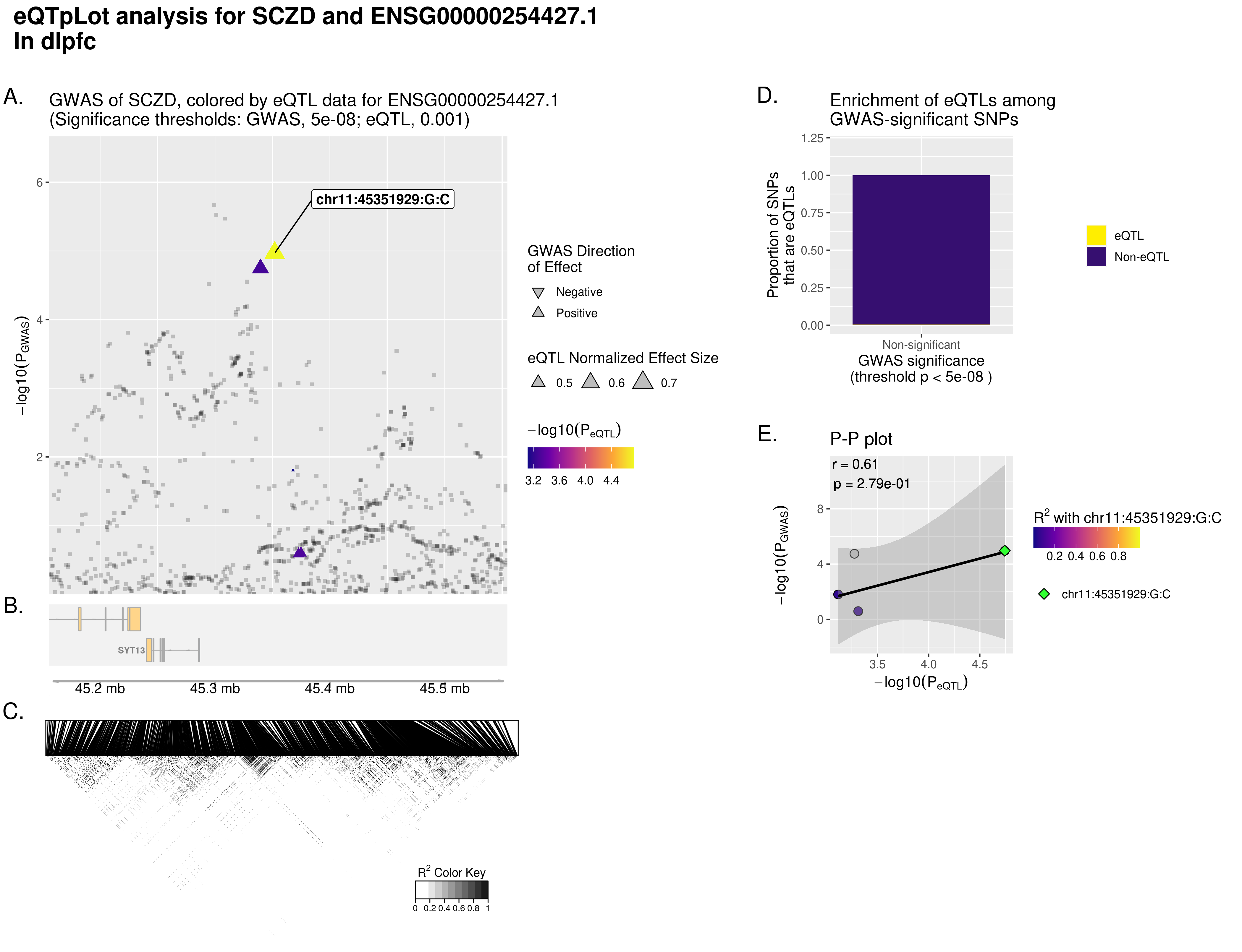

### ENSG00000278434.1.SCZD.dlpfc.WithoutCongruenceData.WithLinkageData.eQTpLot.png

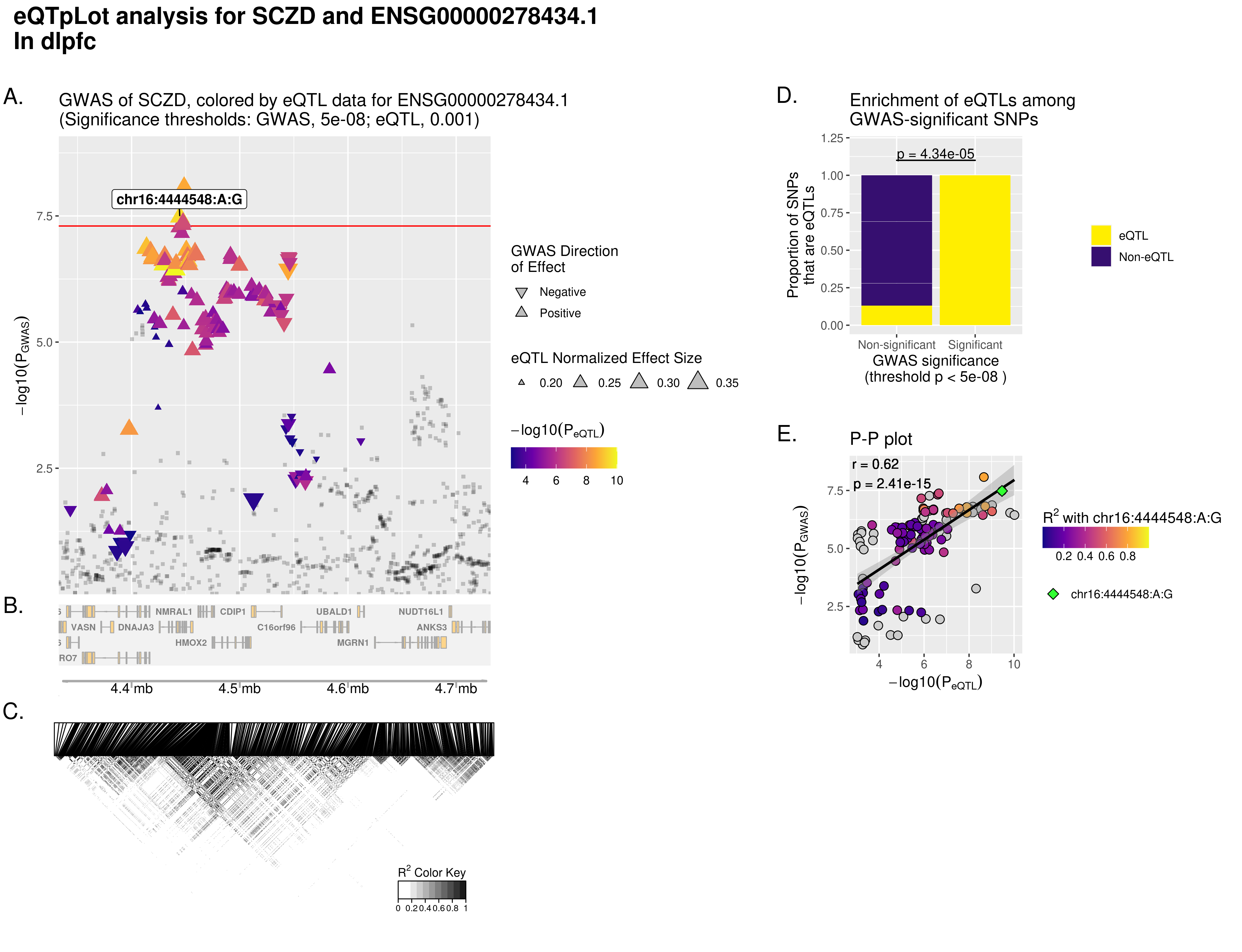

### ENSG00000278434.1.SCZD.dlpfc.WithoutCongruenceData.WithLinkageData.eQTpLot.png

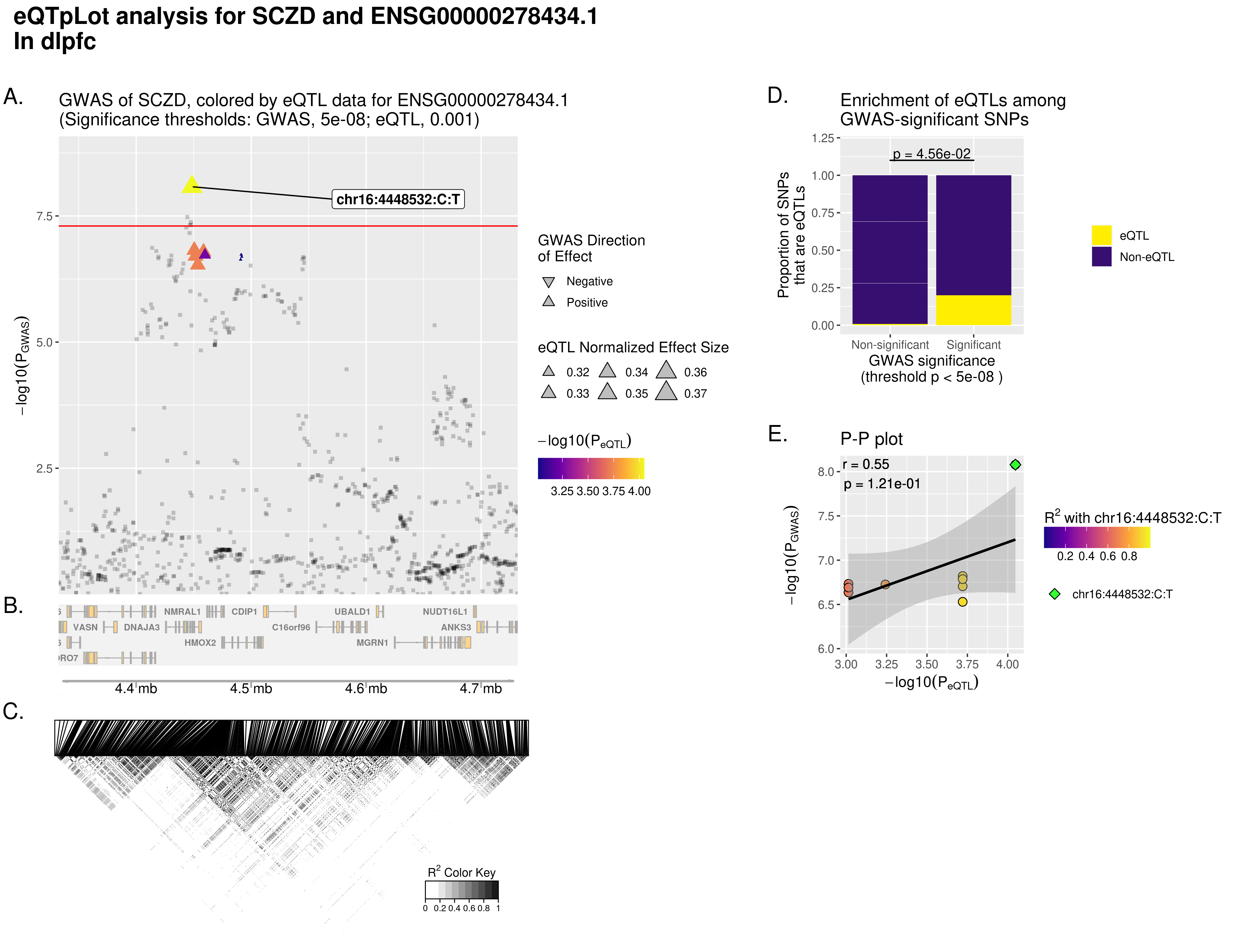

### module_trait_relationships.pdf

Module kME−Trait Correlation

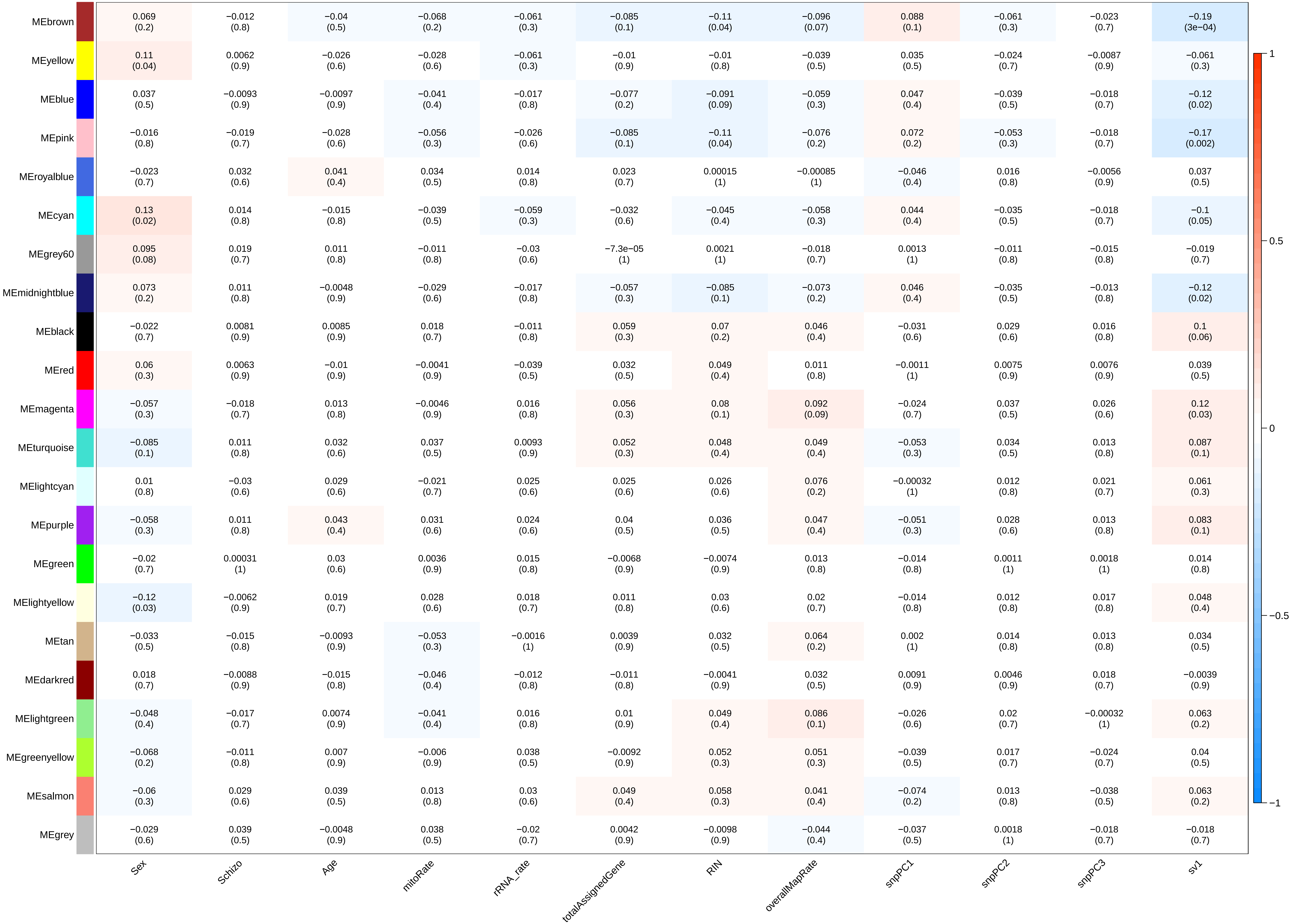

### power_parameter_selection.pdf

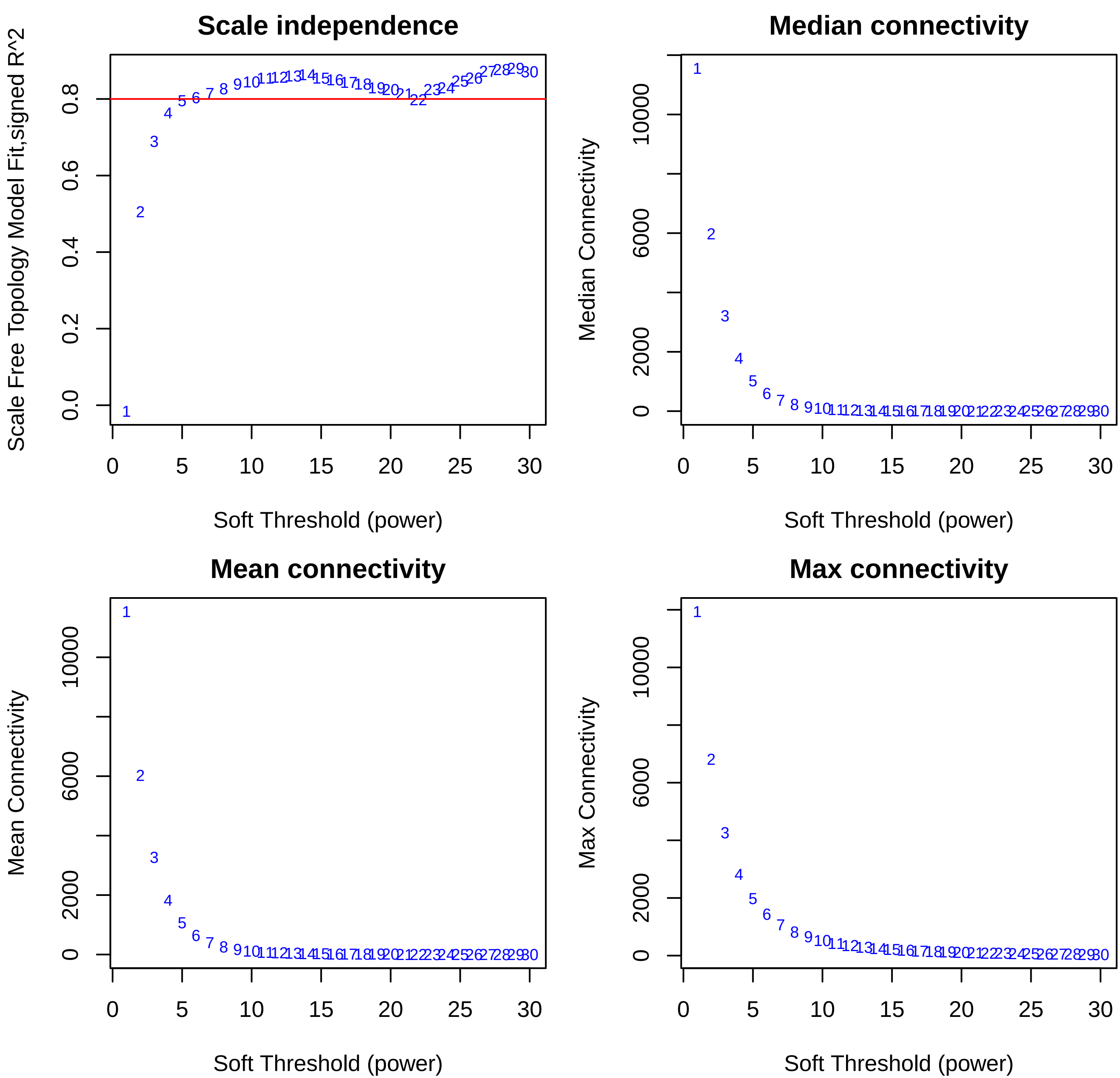

### power_parameter_selection.pdf

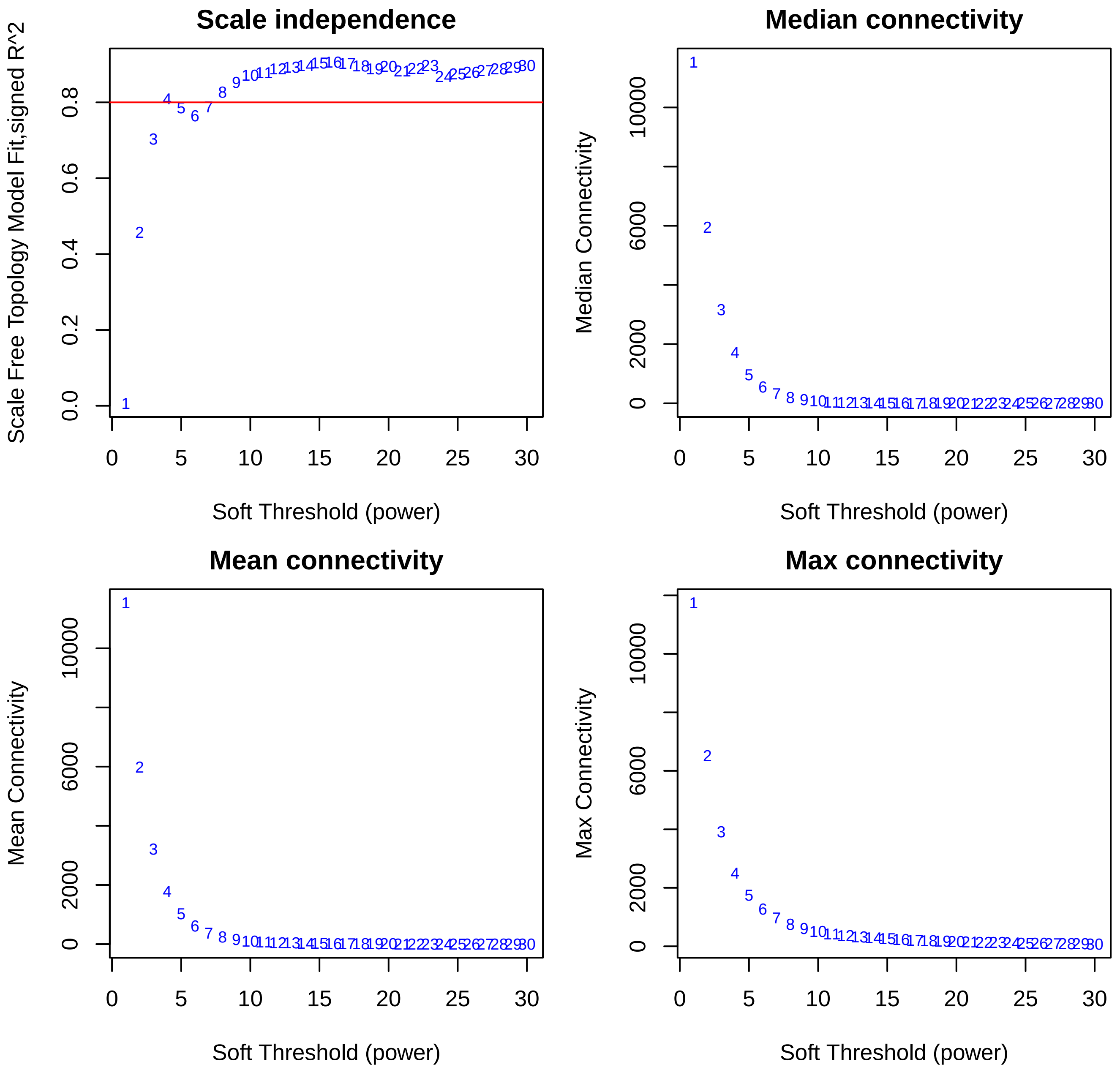

### power_parameter_selection.pdf

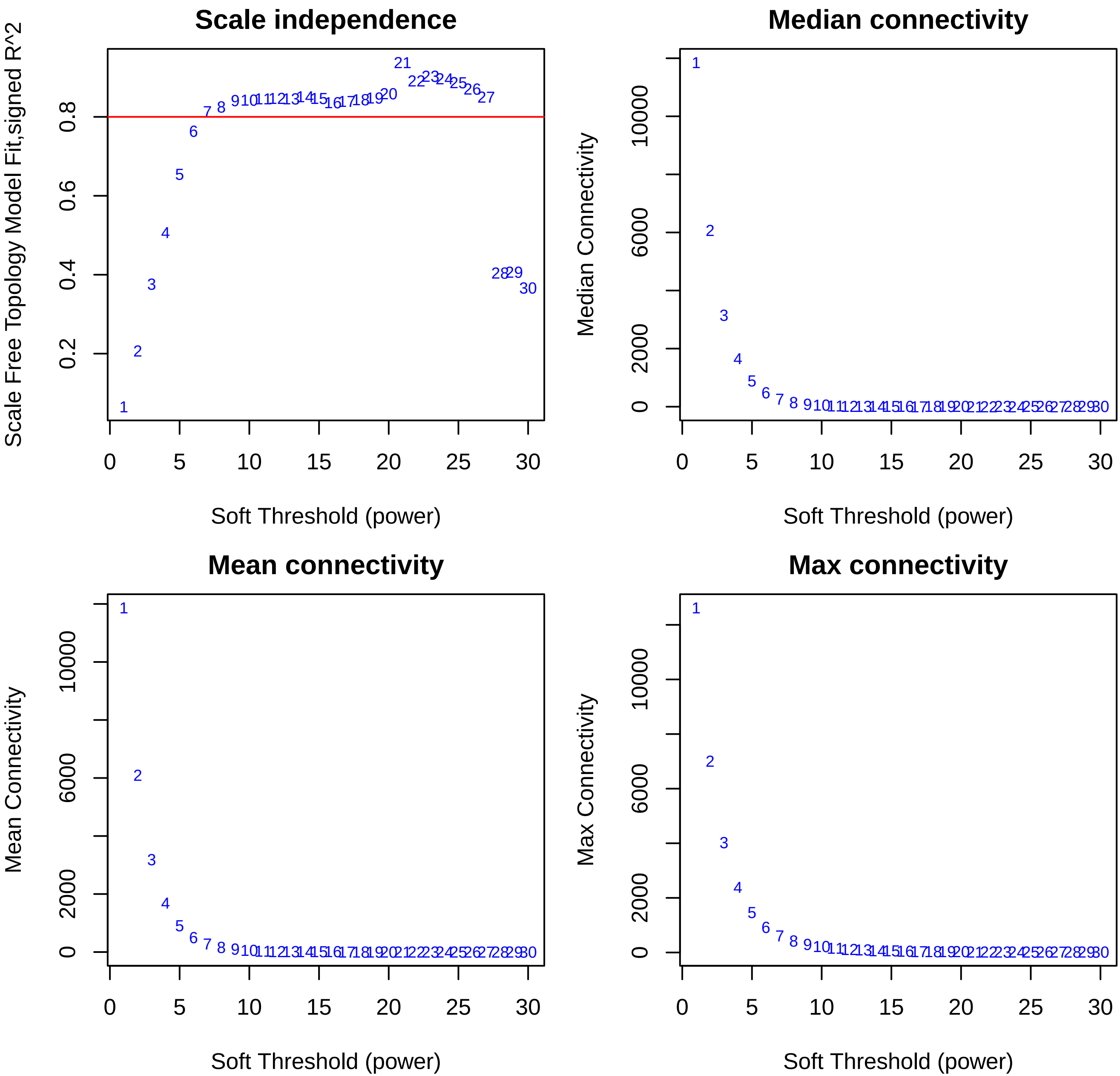

### sample_dendrogram_and_trait_heatmap.pdf

#### Sample dendrogram and trait heatmap

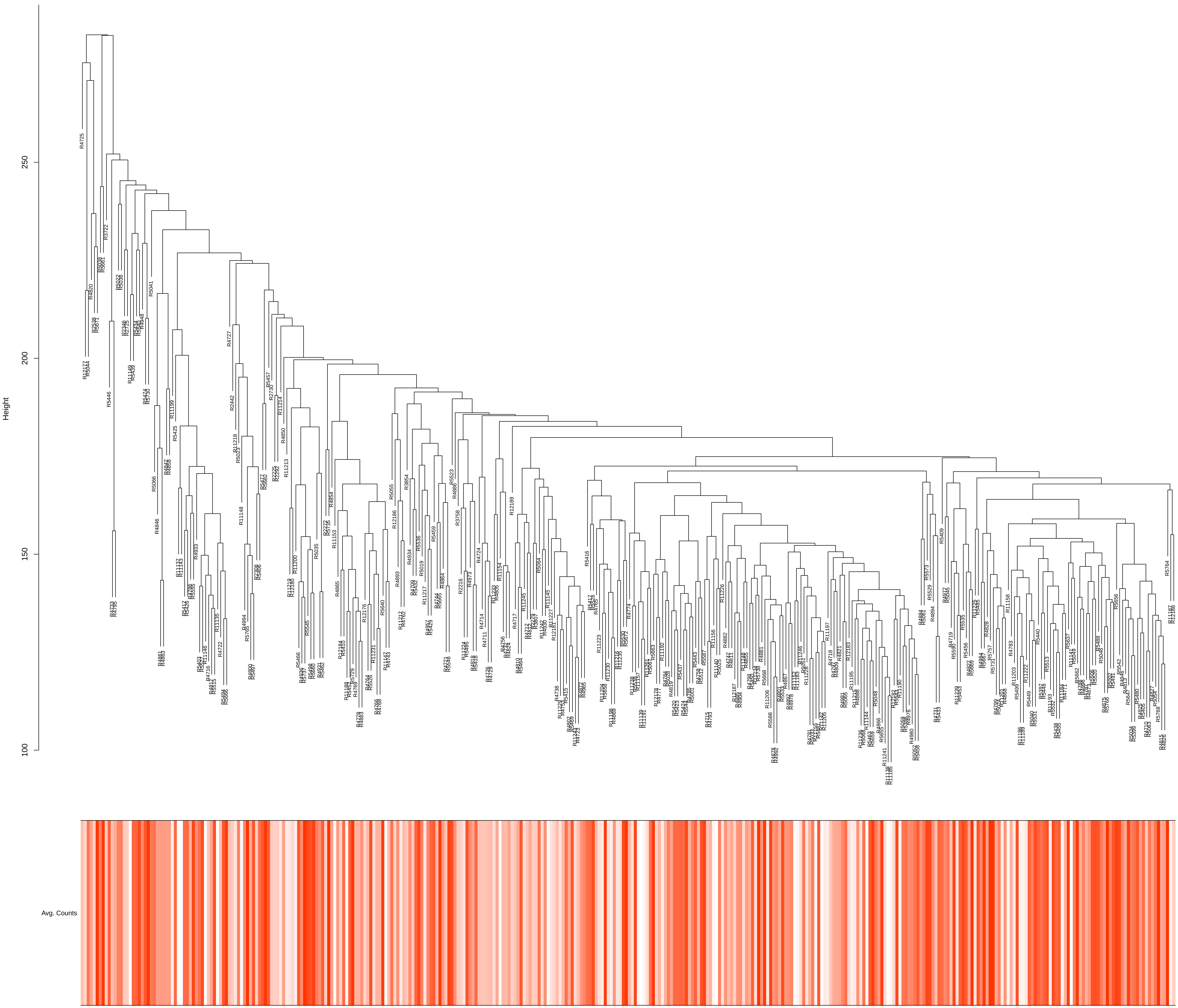

### sample_dendrogram_and_trait_heatmap.pdf

Sample dendrogram and trait heatmap

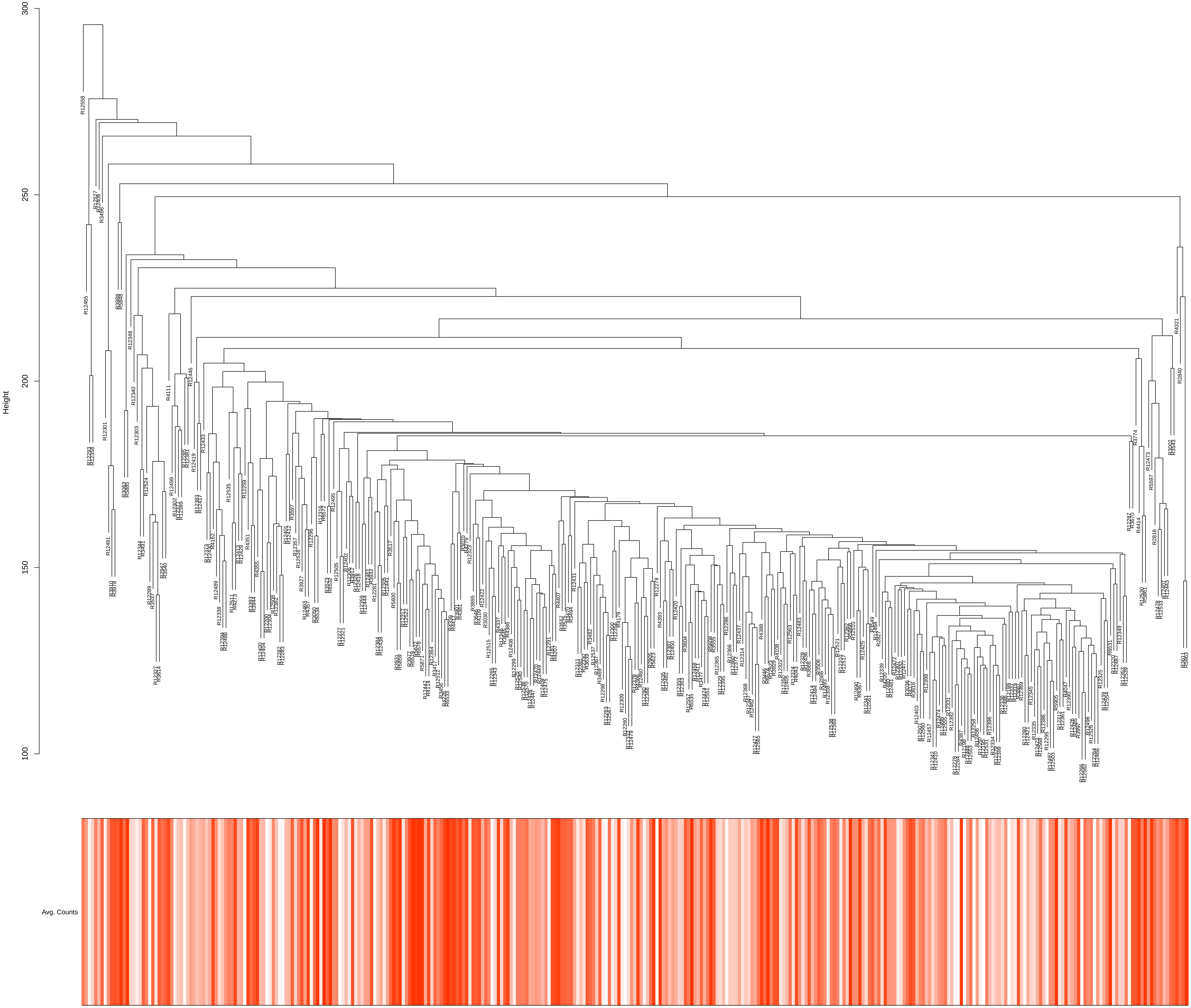

### sample_dendrogram_and_trait_heatmap.pdf

**Sample dendrogram and trait heatmap**

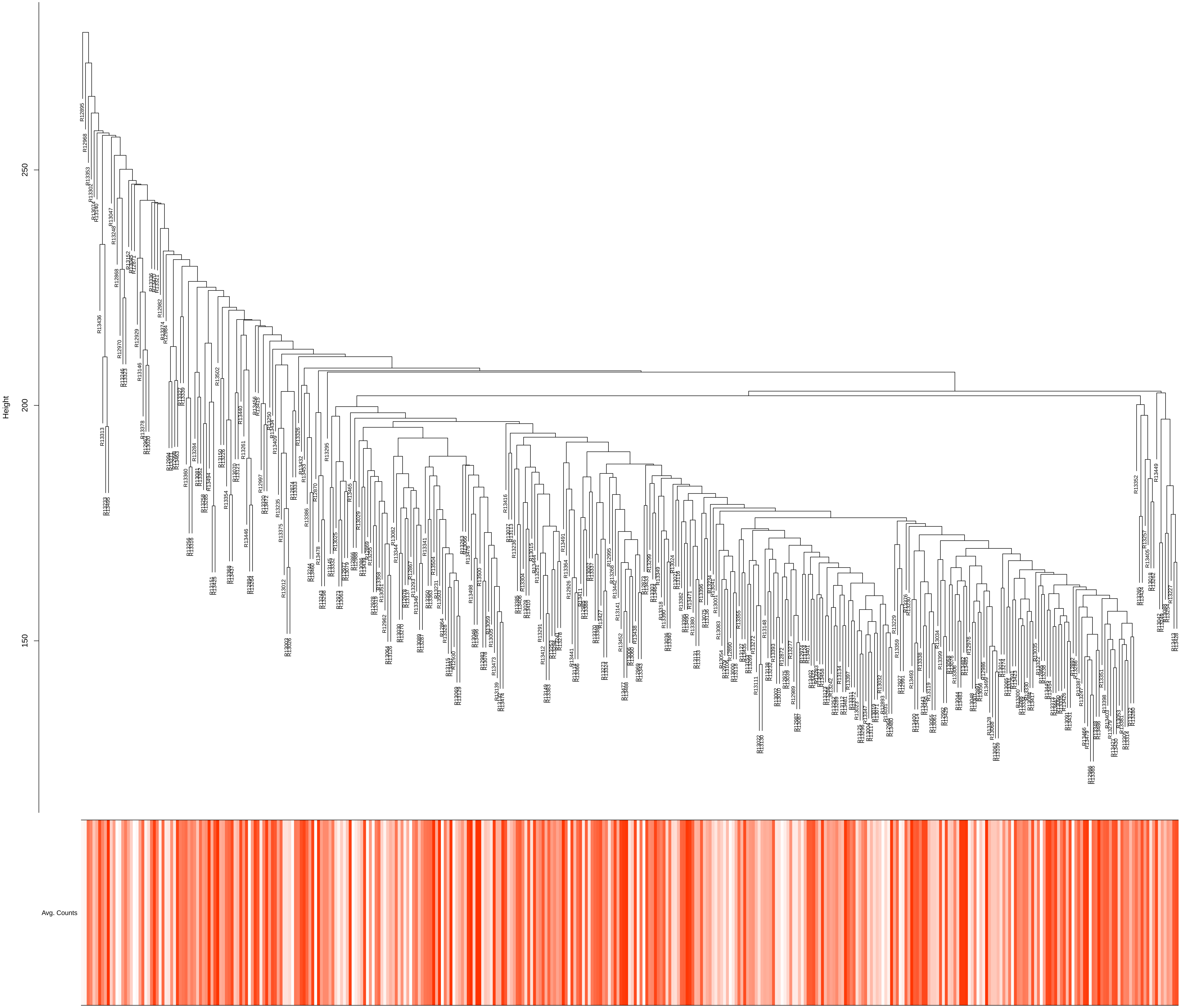

### wgcna_module_enrichment_DEG.pdf

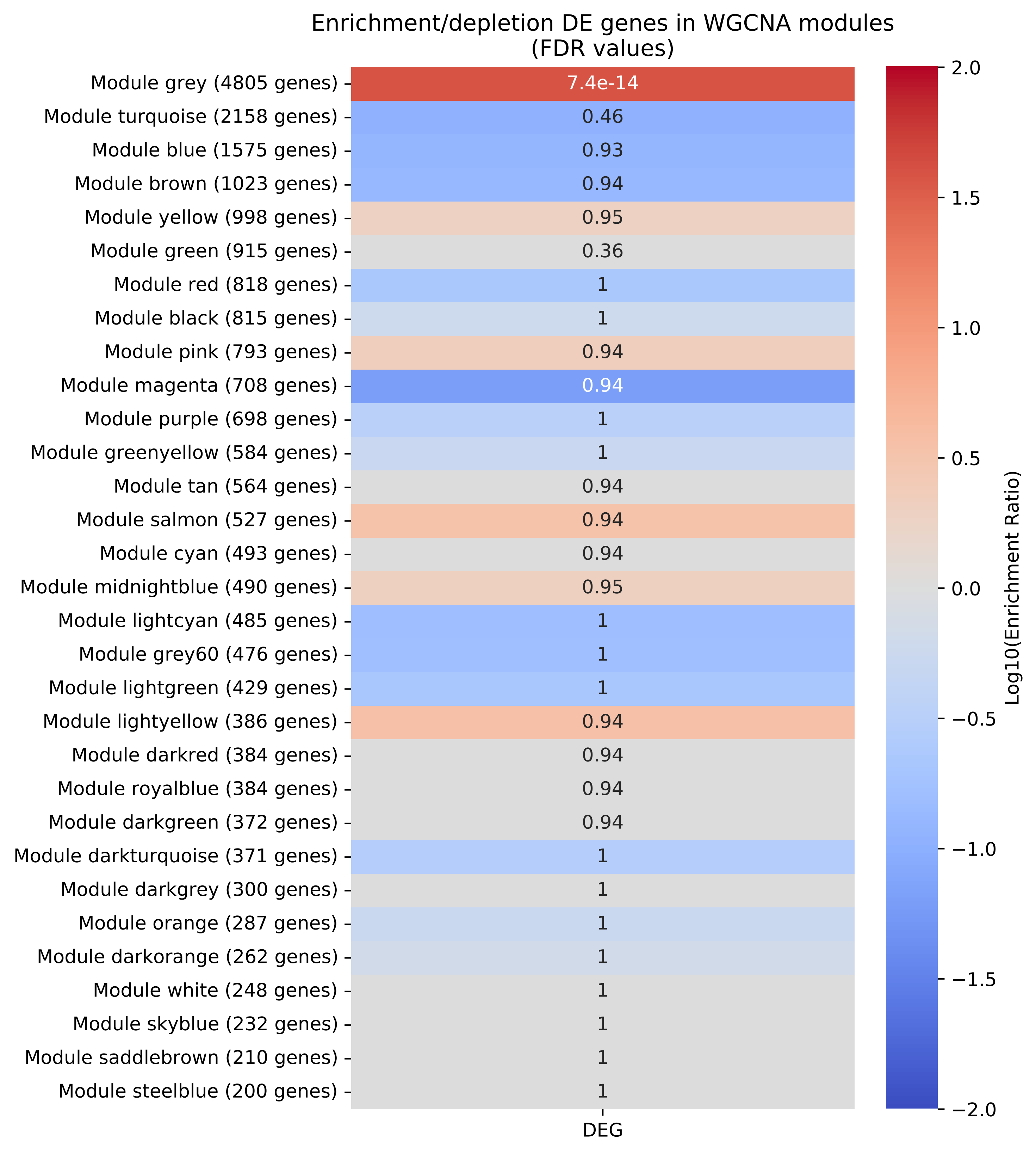

### wgcna_module_enrichment_DEG.pdf

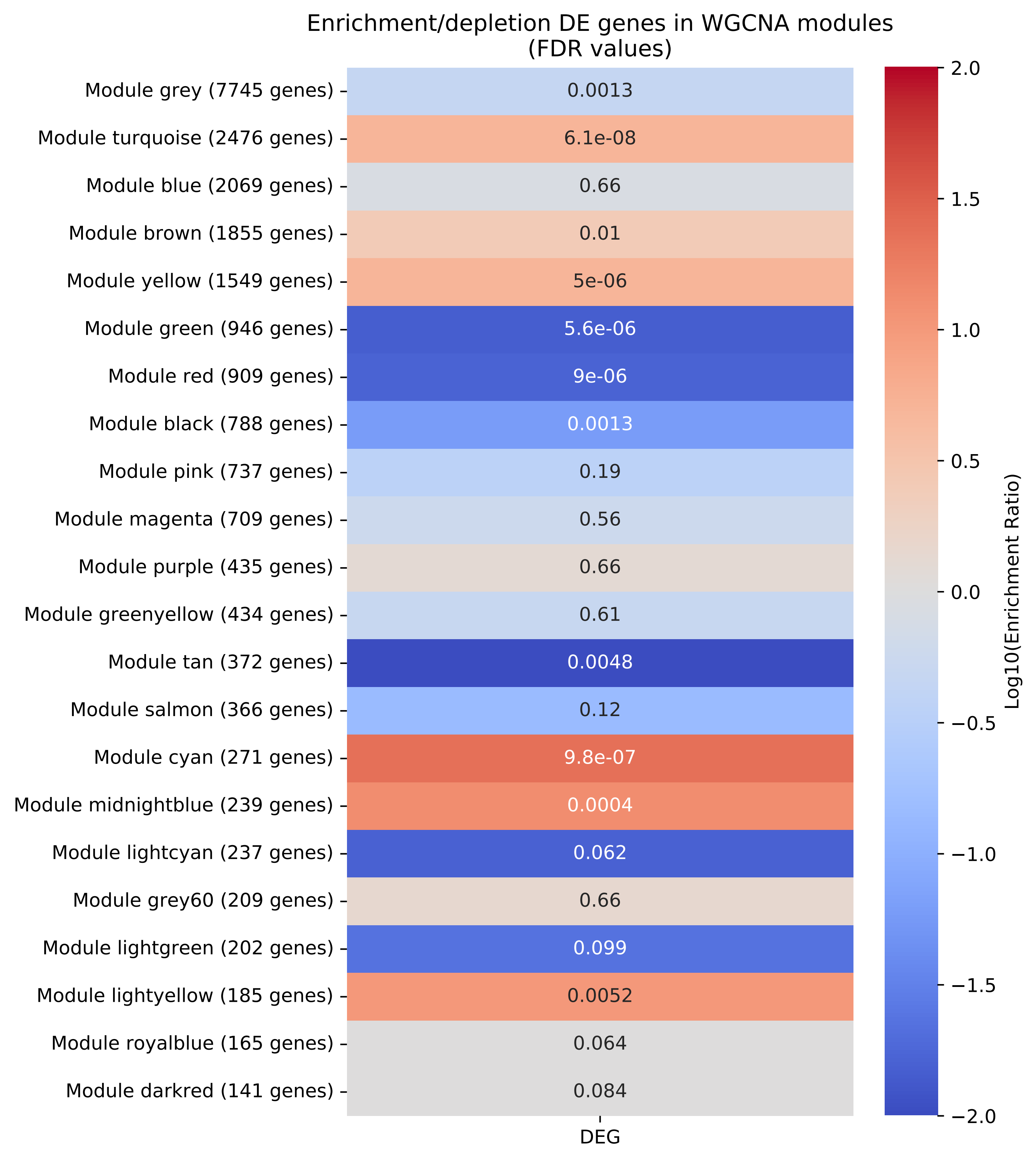

### wgcna_module_enrichment_DEG.pdf

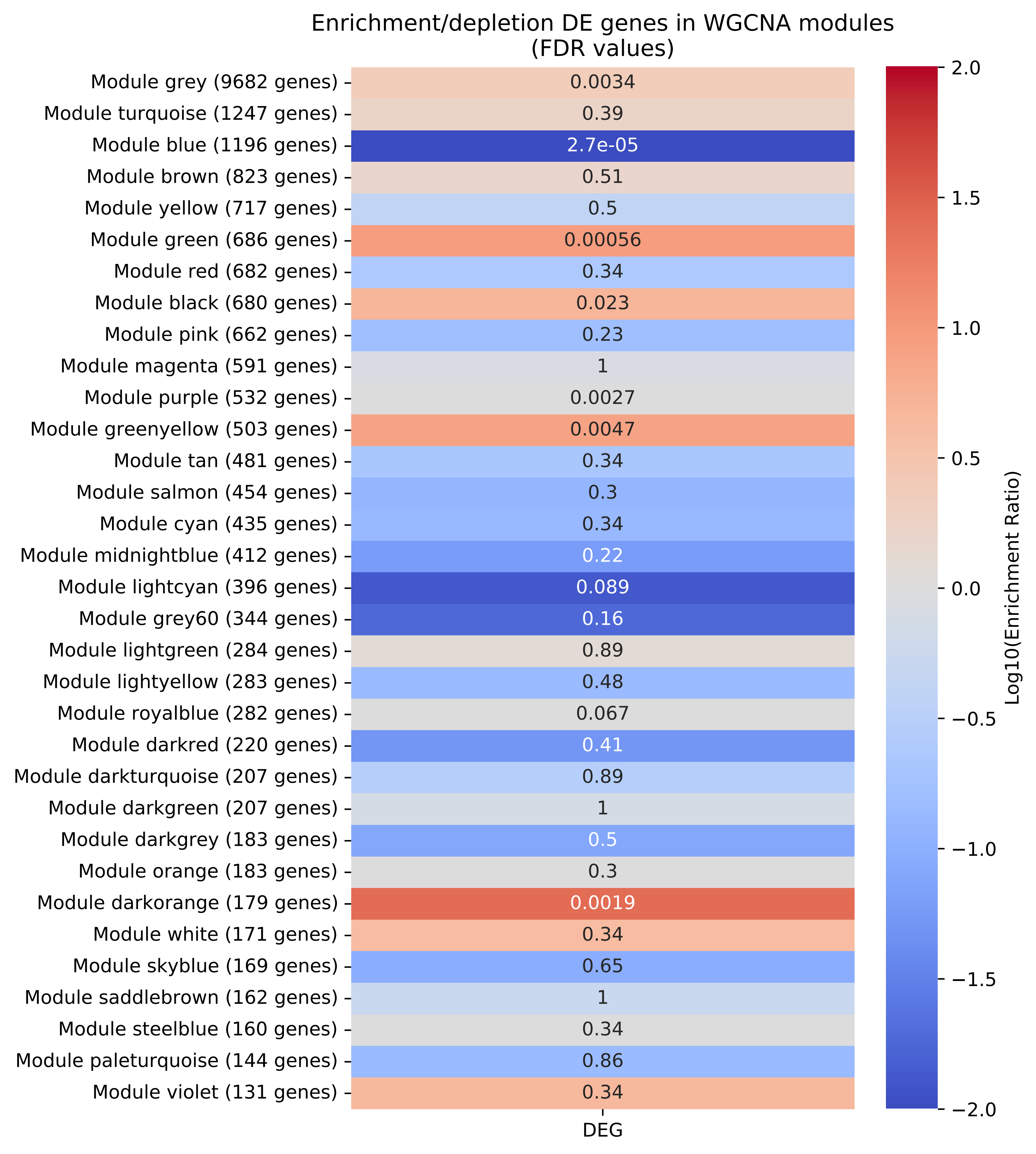

### wgcna_module_enrichment_DEG_noMHC.pdf

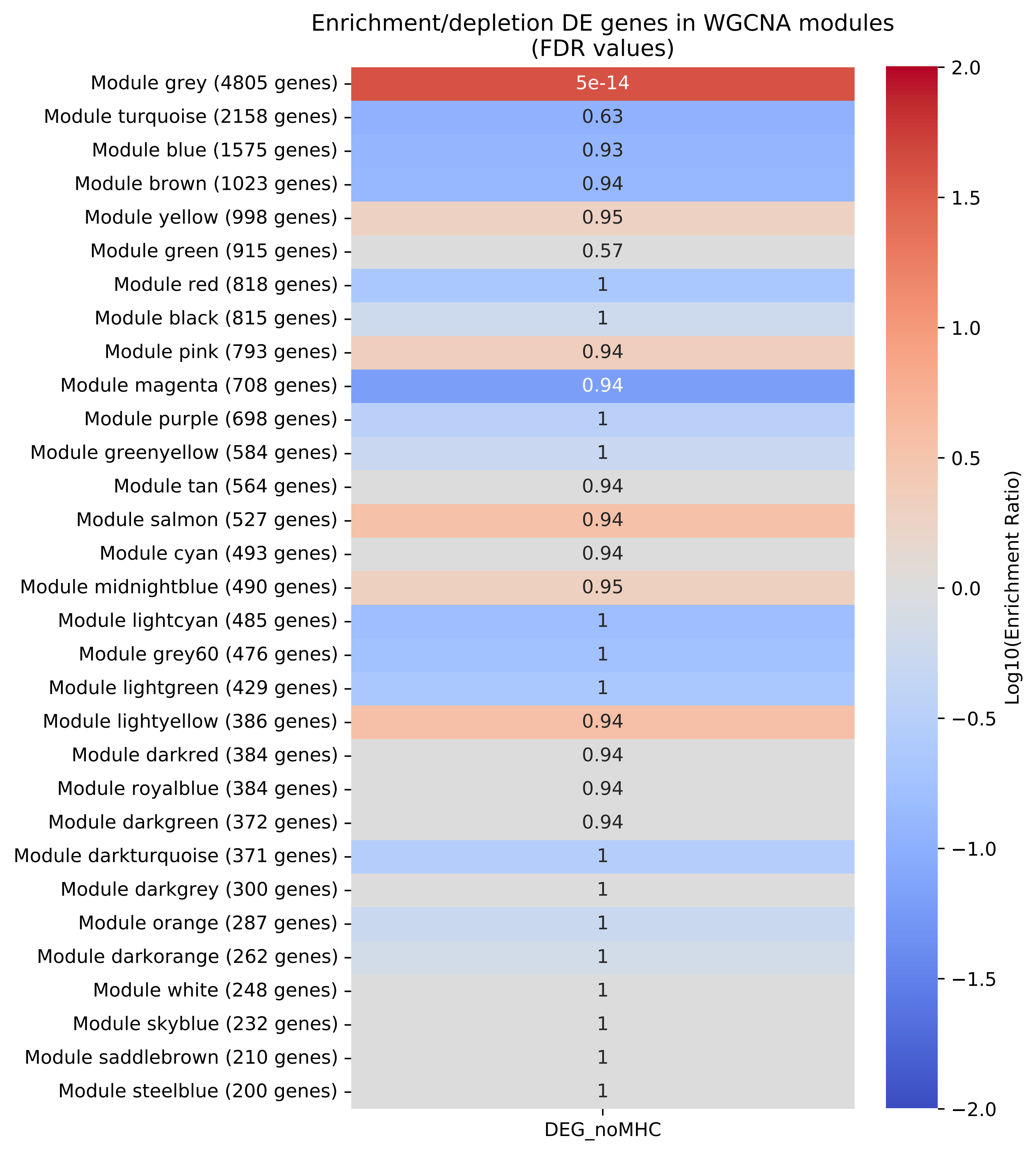

### wgcna_module_enrichment_DEG_noMHC.pdf

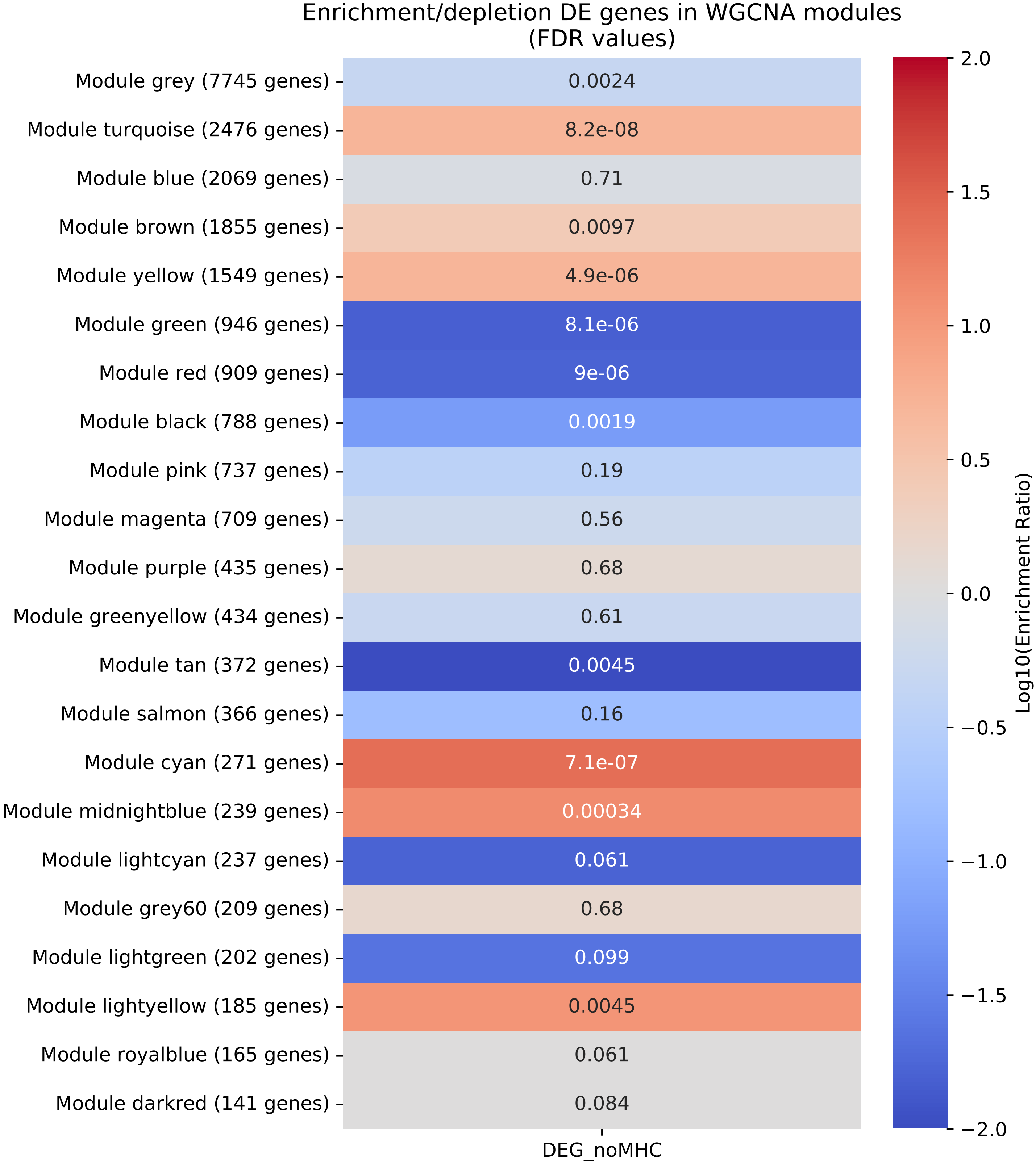

### wgcna_module_enrichment_DEG_noMHC.pdf

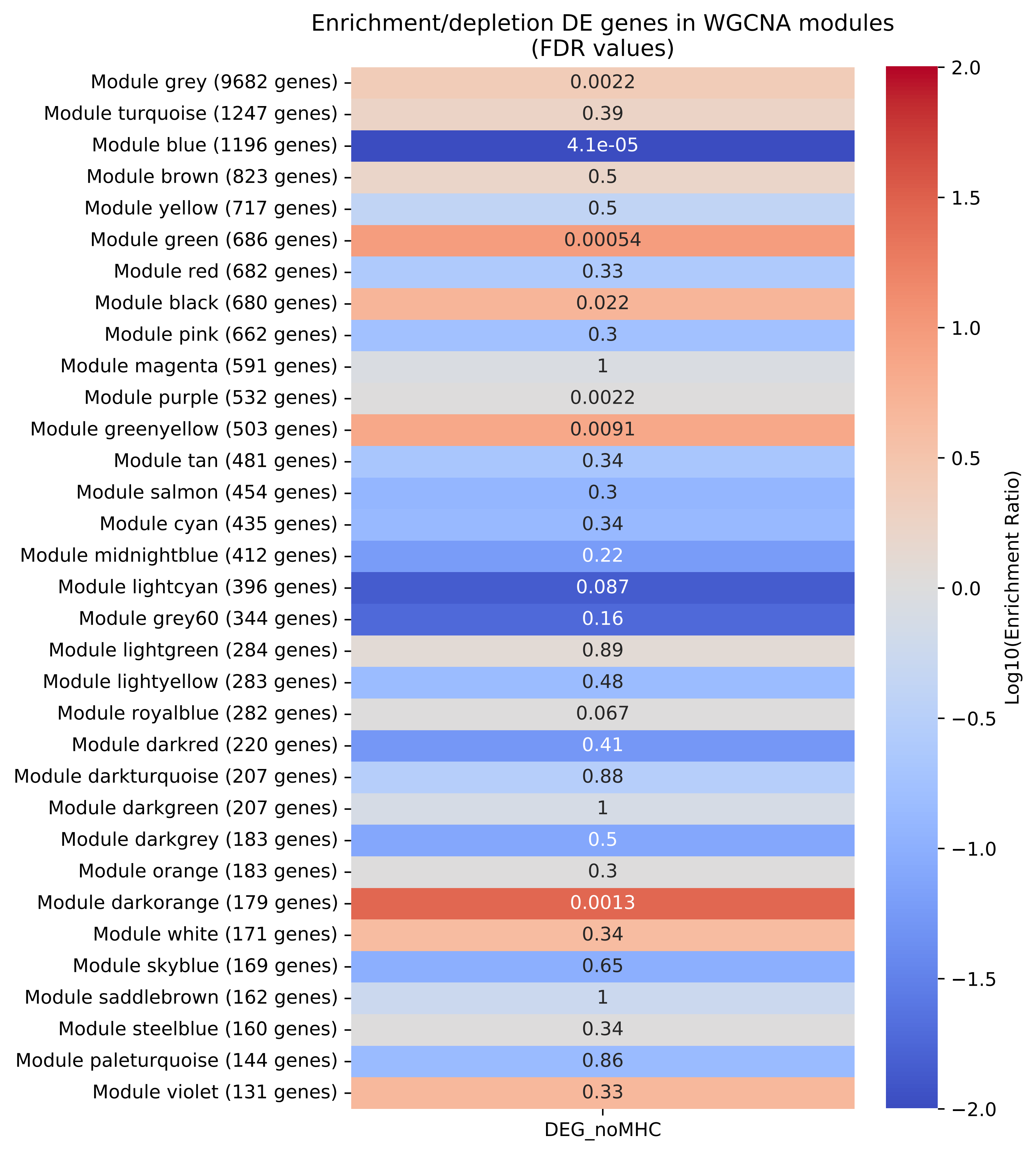
